# Comprehensive genomic profiling of 30,000 consecutive solid tumors

**DOI:** 10.1101/2020.11.19.20233866

**Authors:** Scott A. Tomlins, Daniel H. Hovelson, Jennifer M. Suga, Daniel M. Anderson, Han A. Koh, Elizabeth C. Dees, Brendan McNulty, Mark E. Burkard, Michael Guarino, Jamil Khatri, Malek M. Safa, Marc R. Matrana, Eddy S. Yang, Alex R. Menter, Benjamin M. Parsons, Jennifer N. Slim, Michael A. Thompson, Leon Hwang, William J. Edenfield, Suresh Nair, Adedayo Onitilo, Robert Siegel, Alan Miller, Timothy Wassenaar, William J. Irvin, William Schulz, Arvinda Padmanabhan, Vallathucherry Harish, Anneliese Gonzalez, Abdul Hai Mansoor, Andrew Kellum, Paul Harms, Stephanie Drewery, Jayson Falkner, Andrew Fischer, Jennifer Hipp, Kat Kwiatkowski, Lorena Lazo de la Vega, Khalis Mitchell, Travis Reeder, Javed Siddiqui, Hana Vakil, D. Bryan Johnson, Daniel R. Rhodes

**Author notes:** Corresponding Author: Scott A. Tomlins, M.D., Ph.D., Strata Oncology, 8192 Jackson Road, Suite A, Ann Arbor, MI 48103USA. Conflict of Interest Disclosures: Drs. Tomlins, Hovelson, Falkner, Hipp, Lazo de la Vega, Vakil, Rhodes, Kwiatkowski; and Drewery, Fischer, Mitchell, Reeder, Siddiqui, Vakil, and Johnson are/were equity holders and/or employees of Strata Oncology. Drs. Tomlins and Rhodes, and Johnson are named as co-inventors on a patent issued to Strata Oncology related to MSI status assessment. Drs. Tomlins and Rhodes are named as co-inventors and are included in royalty streams for a patent issued to the University of Michigan regarding ETS fusions in prostate cancer that has been licensed to Hologic/Gen-Probe (sublicensed to Ventana Medical Systems) and LynxDx. They are equity holders in Javelin Oncology. Dr. Tomlins previously served as a consultant to Strata Oncology and has consulted for Astellas/Medivation and Janssen. He has received research (to University of Michigan) funding from Astellas and has received travel support from the Prostate Cancer Foundation. Dr. Burkard reports receiving research funding from Abbvie, Genentech, Puma Biotechnology, Arcus Biosciences, Apollomics, and Loxo Oncology. Dr. Matrana reports receiving fees for serving on the speakers bureau from Pfizer, Janssen, Astellas, AstraZeneca Eisai Pharmaceuticals, Bristol-Myers Squibb, Genentech, SirTex, Merck and as a consultant from AstraZeneca. Dr. Yang reports receiving fees for serving on the advisory board from AstraZeneca, Bayer, Clovis Oncology, and for receiving research support from Eli Lilly and Puma Biotechnology. Dr. Parsons reports receiving fees for serving on the speakers bureau and advisory board from Amgen and Celgene, and for research funding from the Wisconsin Idea Grant, Gundersen Medical Foundation. Dr. Thompson reports receiving consulting fees from Syapse Precision Medicine Council, Elsevier ClinicalPath, Adaptive, UpToDate, GlaxoSmithKline, Takeda, Celgene, Doximity, and institutional research funding from Abbvie, Bristol-Myers Squibb, CRAB CTC, Denovo, Hoosier Research Network, Eli Lilly, LynxBio, Takeda, and TG Therapeutics. Dr. Edenfield reports receiving consulting fees from Chimerix. Drs. Dees, Anderson, Suga, Burkard, Matrana, and Yang receive fees for serving on the Strata Oncology Clinical Advisory Board. The remaining authors have no disclosures.

## Abstract

**Purpose:** Tissue-based comprehensive genomic profiling (CGP) is increasingly utilized for treatment selection in patients with advanced solid tumors, however real-world tissue availability may limit widespread implementation. Here we established real-world CGP tissue availability and assessed CGP performance on consecutively received samples.

**Patients and Methods:** Post-hoc, non-prespecified analysis of 32,048 consecutive tumor tissue samples received for StrataNGS, a multiplex PCR based-CGP (PCR-CGP) test, as part of an ongoing observational trial (NCT03061305). Tumor tissue sample characteristics and PCR-CGP performance were assessed across all tested tumor samples, including exception samples not meeting minimum input requirements (<20% tumor content [TC], <2mm^2^ tumor surface area [TSA], DNA or RNA yield <1ng/ul, or specimen age >5yrs). Tests reporting at least one prioritized alteration or meeting all sequencing QC metrics (and ≥20% TC) were considered successful. For prostate carcinoma and lung adenocarcinoma, tests reporting at least one actionable/informative alteration or those meeting all sequencing QC metrics (and ≥20% TC) were considered actionable.

**Results:** PCR-CGP was attempted in 31,165 of 32,048 (97.2%) consecutively received solid tumor tissue samples. Among the 31,165 tested samples, 10.7% had low (<20%) tumor content (TC) and 58.4% were small (<25mm^2^ TSA), highlighting the challenging nature of samples received for CGP. Of the 31,101 samples evaluable for input requirements, 8,079 (26.0%) were exceptions not meeting requirements. However, 94.2% of the 31,101 tested samples were successfully reported, including 80.6% of exception samples. Importantly, 80.6% of 1,344 tested prostate carcinomas and 87.8% of 1,144 tested lung adenocarcinomas yielded results informing treatment selection.

**Conclusion:** Most real-world tumor tissue samples from patients with advanced cancer desiring CGP are limited, requiring optimized CGP approaches to produce meaningful results. An optimized PCR-CGP test, coupled with an inclusive exception testing policy, delivered reportable results for >94% of samples, potentially expanding the proportion of CGP-testable patients, and thus the impact of biomarker-guided targeted and immunotherapies.

## Introduction

Molecular profiling of patient tumor specimens is increasingly important as more cancer therapies are indicated for use in biomarker-defined patient populations ^1^. Next generation sequencing (NGS) has emerged as the diagnostic method of choice to assess relevant biomarkers simultaneously from a single formalin-fixed paraffin embedded (FFPE) tumor tissue ^2-10^ or circulating cell free DNA (cfDNA) liquid biopsy sample ^11-14^. The U.S. Center for Medicare and Medicaid Services (CMS) has deemed tissue based comprehensive genomic profiling (CGP) by NGS—which includes evaluation of single-/multi-nucleotide variants (SNVs), short frame preserving insertions/deletions (indels), copy number amplifications and deep-deletions, gene fusions, microsatellite instability (MSI) status, and tumor mutation burden (TMB)—medically necessary for patients with advanced solid tumors (National Coverage Decision [NCD] CAG-00450N and MolDX Local Coverage Decision [LCD] L38045).

Successful FFPE tissue CGP requires the isolation of adequate quantity and quality of nucleic acid from tumor cells. Several challenges affect real world CGP applicability, including 1) minute tissue specimens that do not yield enough nucleic acid, 2) tissue samples with low tumor content (TC), such that the majority of cells do not harbor tumor-defining mutations and 3) low quality nucleic acid, which is affected by sample age and fixation ^15^. To ensure successful testing, most available CGP tests have tumor tissue size (generally measured in mm^2^ of tumor surface area [TSA]), TC (the # tumor cells / # nucleated cells in the area used for testing) and nucleic acid yield/input requirements ^3,5,15^. However, CGP test failure rates of 30-50% in recent clinical trial cohorts, where testing is attempted on all received samples (or those received below input requirements are considered failures), suggest that current CGP approaches, which are largely based on hybrid capture library preparation, may only be applicable for a subset of available tissue specimens^16-19^.

Herein, we sought to characterize the attributes of >30,000 consecutive real-world samples submitted for CGP from an observational clinical trial (NCT03061305) and the performance of a multiplex PCR based (PCR-) CGP test, StrataNGS, as applied to consecutively received and tested samples, including those below sample input requirements.

## Materials and Methods

### Patient Cohort

The Strata Trial (NCT03061305) is a 100,000-patient observational study for patients with advanced solid tumors that leverages NGS to: 1) Understand the impact of NGS on treatment selection, including clinical trial enrollment, 2) Test the feasibility of a system-wide NGS screening protocol across a network of health systems, and 3) Understand the relationships between NGS biomarkers and patient outcomes, including treatment response. At enrolling health systems, all adult patients with unresectable or metastatic solid tumors and available FFPE tumor tissue are eligible. No power analysis was performed to determine the sample size for the post-hoc, case series analysis presented herein, which is focused on received sample characteristics and PCR-CGP performance; these do not represent primary objectives of the trial. All sample, sequencing QC metrics, and clinically reported PCR-CGP results from StrataNGS testing were retrieved from StrataPOINT, a deidentified database of Strata Trial Results, and all samples received for testing from February 13, 2017 to June 25, 2020 were included. The Strata Trial has been reviewed and approved by an IRB prior to study start.

### CGP Testing

StrataNGS is a 429 gene PCR-CGP laboratory developed test for FFPE tumor tissue samples performed on co-isolated DNA and RNA, which has been validated on over 1,900 clinical FFPE tumor samples, and is covered for Medicare beneficiaries with advanced solid tumors after undergoing MolDX technical assessment (^20,21^ and Tomlins *et al*., Manuscript submitted). The current version of the StrataNGS test includes all CGP variant classes (mutations, indels, copy number alterations, MSI status and TMB reported from two DNA panels; fusions reported from an RNA panel). An earlier version of StrataNGS ^21^ did not include TMB, and testing has been performed in different pooling configurations during the study period, however as specimen requirements have not changed regardless of test version or configuration, all received samples during the study time period were included.

StrataNGS requires one FFPE block or 10 x 5um-thick unstained slides from the part with the greatest TSA and highest TC. Based on preclinical testing and limits of detection (LOD), minimum sample tissue input requirements are TSA ≥ 2mm^2^, TC ≥ 20% and time from sample acquisition < 5 years. Samples that do not meet these requirements but with identifiable and isolatable tumor (TSA ≥ 0.1mm^2^) are deemed exceptions and testing is still attempted, given our experience that meaningful results can often be obtained in a large proportion of samples. Macrodissection is performed as necessary to enrich TC, and DNA and RNA are co-isolated from tissue specimens. Nucleic acid concentration input requirements are ≥1ng/uL for both DNA and RNA; however, samples not meeting this requirement were also deemed exceptions, and testing proceeded. After PCR-CGP by NGS on semi-conductor (Ion Torrent) sequencers, a series of sequencing quality control (QC) assessments are performed per variant type and a molecularly informed TC is determined as described (^20,21^ and Tomlins et al., Manuscript submitted). For samples that fail one or more sequencing QC assessments or have a final molecularly informed TC < 20%, which is the StrataNGS LOD for most alteration types, positive alterations may still be called via an expert molecular pathology review process that considers the sequencing evidence and clinical context; however other alterations cannot be definitively ruled out, thus yielding a partial test result.

In the reported study period, verification of the molecularly informed TC, variant review, and test sign out for all cases was performed by a single board-certified pathologist with significant anatomic and molecular pathology experience (S.A.T.), using an in house developed review/sign-out portal. Tests with one or more reported prioritized alterations (e.g. oncogenic mutations or amplifications in oncogenes, deleterious mutations or deep deletions [equivalent to two-copy loss] in tumor suppressors, gene fusions involving driver genes, MSI-high [MSI-H] or TMB-high) or with no prioritized alterations but passing all sequencing QC assessments and having TC ≥ 20% were considered successfully reported. Tests with no reported prioritized alterations and failing one or more sequencing QC assessments (or having TC < 20%), were considered failures. To characterize PCR-CGP performance, samples were binned into a single group (in the following order) based on input sample exception criteria, followed by TSA: (1) TC < 20%, (2) TSA < 2mm^2^, (3) sample age > 5 years, (4) DNA or RNA concentration < 1ng/uL, (5) TSA < 25mm^2^ and (6) TSA >= 25mm^2^. Samples without recorded DNA/RNA concentration information in the deidentified database were considered to meet sample input criteria if all other input characteristics were met, as >90% of samples with TSA ≥ 2mm^2^ yielded at least 1ng/ul DNA/RNA (**Supplementary Table 1**).

### External CGP Dataset

For overall biomarker comparisons in this PCR-CGP cohort to an external hybrid capture pan-solid tumor CGP cohort, we obtained biomarker frequencies from the Memorial Sloan Kettering (MSK)-IMPACT cohort of 10,917 clinical tumor specimens profiled by hybrid capture CGP (limited to a single sample per subject)^7^. For the MSK-IMPACT cohort, mutations and copy number alterations of unknown significance were excluded (as in cBioPortal^22^). For all comparisons, MSK-IMPACT primary and secondary tumor type was mapped to the PCR-CGP cohort tumor types; to best facilitate meaningful comparisons between cohort-specific biomarker frequencies, several PCR-CGP tumor types were combined into hybrid tumor type groupings (‘Biliary_Liver’: Biliary and Liver, and ‘Esophagus_Stomach’: Esophagus and Stomach) for frequency calculations and comparison analyses. PCR-CGP biomarker frequency calculations utilized all input requirement evaluable samples that were successfully reported (as defined above) with ≥20% TC, limiting comparison to CGP variant classes reportable by the version of PCR-CGP test used. PCR-CGP and MSK-IMPACT biomarker frequencies / frequency comparisons were restricted to tumor types with ≥50 samples in the PCR-CGP cohort. Samples with multiple reported variants in a single gene + biomarker category (e.g., two *TP53* mutations) were only counted once in corresponding frequency calculations.

Overall and tumor-specific frequencies presented and compared in this paper were restricted to mutation or copy number alteration (DNA) or chimeric gene fusion (RNA) biomarker categories reportable by both PCR-CGP and MSK-IMPACT tests. Frequencies were calculated for each unique gene / biomarker category combination, using the following MSK-IMPACT biomarker categories: MUT, HOMDEL, AMP, and FUSION. PCR-CGP reported SNVs and indels were grouped together for each gene-specific MUT frequency calculation, while clinically reported PCR-CGP copy number amplifications and deletions were mapped to MSK-IMPACT AMP and HOMDEL labels (respectively) to facilitate frequency comparisons. Pan tumor biomarkers including microsatellite instability (MSI) and tumor mutation burden (TMB) were excluded from all frequency comparisons, as were all biomarker classes associated with tumor suppressors reported in the MSK-IMPACT cohort with >20% germline frequency after excluding low VAF alterations as germline mutations are not available from MSK-IMPACT for all of these genes (*ATM, MSH2, MSH6, BRCA1*, and *BRCA2*^*23*^).

Differences in assay design and clinical reporting approaches necessitated special handling for several biomarkers. For instance, given MSK-IMPACT’s assay intronic tiling, only *EGFR-SEPT14* fusions were considered eligible for PCR-CGP *EGFR* fusion frequency calculations. Further, the PCR-CGP test leverages DNA and RNA-based evidence to identify *MET* exon 14 deletion (METex14del) events, whereas MSK-IMPACT considers strictly DNA-based tiling of potential METex14del breakpoints. To simplify cohort comparisons, PCR-CGP and MSK-IMPACT *MET* mutation frequencies reported here (e.g., ‘*MET* MUT’ frequencies) are restricted solely to METex14del events. For PCR-CGP METex14del rates, any case with a DNA-(*MET* exon 14 splice donor or acceptor mutations) and/or RNA-based METex14del variant call was considered to be METex14del (or ‘MET MUT’) positive. *ERG, ETV1, ETV4* and *ETV5* fusion frequencies were only assessed in prostate cancer, given the intronic tiling of the most common 5’ partner (*TMPRSS2*) of ETS gene fusions in prostate cancer and the inclusion of only prostate fusion related 5’ partners for these genes in the PCR-CGP test.

For both PCR-CGP and MSK-IMPACT cohorts, overall and tumor-specific biomarker frequencies were calculated as the number of positive cases divided by the total number of tumor-specific or overall cohort samples. Frequency Pearson correlations were calculated using the Python (v3.6.5) pandas (v1.0.0) package corr() ^24^.

### Prostate Cancer Analysis

To characterize sample characteristics and test performance for treatment selection in prostate cancer, we considered all input characteristic evaluable consecutively tested samples using the current StrataNGS test version and all results were classified as either ‘informative’ or ‘non-informative’. We defined informative results as: 1) testing positive for a therapy selection biomarker from NCCN guidelines (MSI-H or deleterious mutation/deep deletion [homozygous loss if diploid] in *BRCA1, BRCA2, ATM, MSH2* or *MSH6* ^*25*^; or 2) testing definitively negative for all therapy selection biomarkers, which requires passing all sequencing QC assessments for the applicable variant classes (mutations, copy number alterations and MSI status), as well as having ≥20% TC.

### Lung Adenocarcinoma Analysis

To characterize sample characteristics and test performance for treatment selection in non-small cell lung cancer (NSCLC) adenocarcinoma, we considered all input characteristic evaluable consecutively tested ‘Lung – NSCLC’ samples assessed by the current StrataNGS test version during the time period when NSCLC subtypes were prospectively recorded at the time of histopathology review (to ensure only adenocarcinoma were included). All results were classified as either ‘informative’ or ‘non-informative’. We defined informative results as: 1) testing positive for a therapy selection biomarker or a primary oncogenic driver ^26^ thought to be mutually exclusive with therapy selection biomarkers (**Supplementary Table 2)**;or 2) testing definitively negative for all therapy selection biomarkers, which requires passing all sequencing QC assessments for the applicable variant classes (mutations, copy number alterations, MSI status, gene fusions, TMB), as well as having ≥20% TC.

## Results

### Characteristics of samples received for CGP

CGP testing was performed by a single CLIA-certified, CAP-accredited laboratory (Strata Oncology, Ann Arbor, MI) as part of an observational clinical trial evaluating the impact of solid tumor sequencing in the advanced cancer setting (NCT03061305) using a previously validated PCR-CGP test (StrataNGS) that is covered for all Medicare beneficiaries after technical review by MolDX. Across 28 diverse U.S. health systems, 31,165 consecutive unique solid tumor samples (from 30,565 unique patients) were received for CGP testing between February 13, 2017 and June 25, 2020; an additional 883 samples were rejected (**Figure 1A**). Although most rejections were for no identifiable tissue or tumor, samples were also rejected for being decalcified, having incorrect subject or specimen information, or when multiple samples were received from the same procedure. As characteristics for these samples were not available in the deidentified dataset, these samples were excluded from further analysis.

**Figure 1.**
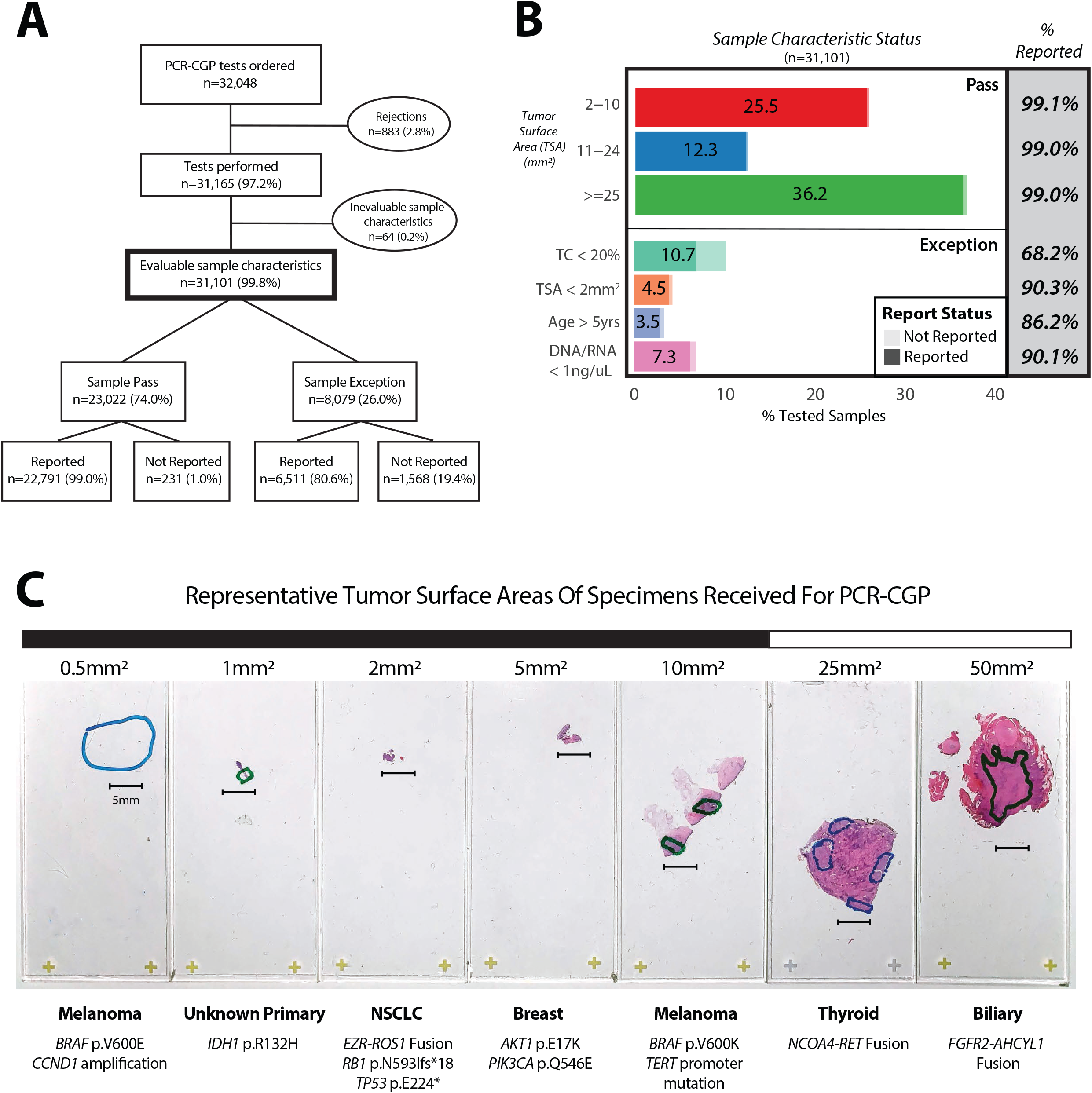
**A)** Breakdown of consecutive PCR-CGP tests ordered from a single commercial clinical sequencing provider between February 13, 2017 and June 25, 2020, including the number of samples rejected prior to testing, the number of tests performed, the number of samples with evaluable input characteristics, and the number of PCR-CGP tests successfully reported. Samples were grouped into those meeting (‘Pass’) or not meeting (‘Exception’) PCR-CGP input requirements. **B)** For all samples with evaluable input characteristics, the distribution of samples per characteristic are shown. For samples passing all input requirements (‘Pass’), samples are stratified by tumor surface area (TSA); exception samples were stratified by indicated sample attribute (‘TC < 20%’: tumor content < 20%; TSA < 2mm^2^; ‘Age > 5yrs’: specimen collected > 5 years prior to PCR-CGP; ‘DNA/RNA < 1ng/uL’: DNA and/or RNA concentration < 1ng/uL). For each sample characteristic category, the proportion of the total number of samples with evaluable input characteristics is shown within the bar; the percentage successfully reported samples is indicated by darker shading in the stacked bar chart and displayed numerically in the gray box at right. **C)** Representative successfully reported samples received for PCR-CGP across a TSA range (small [<25mm^2^] and large [≥ 25mm^2^] samples indicated). Cancer types and selected prioritized alterations are shown.

Sample characteristics from all 31,165 consecutively tested samples are shown in **Table 1**, with cohort demographics and characteristics by tumor type shown in **Supplementary Tables 3 and 4**. Importantly, testing in the Strata Trial during the study period was not restricted by tumor type, was provided at no cost to patients with advanced cancer (even if prior single gene testing had been performed), and enrolled patients from >25 diverse U.S. health care systems. With the minimal tumor tissue specimen recommendations provided to sites (≥20% TC and TSA ≥ 2mm^2^), and the number of specimens received without any tissue/tumor or not meeting these requirements (see above and below), given our general approach of attempting testing even in all input exception samples, this cohort uniquely informs on general tissue availability for patients with advanced cancer desiring CGP.

**Table 1.**
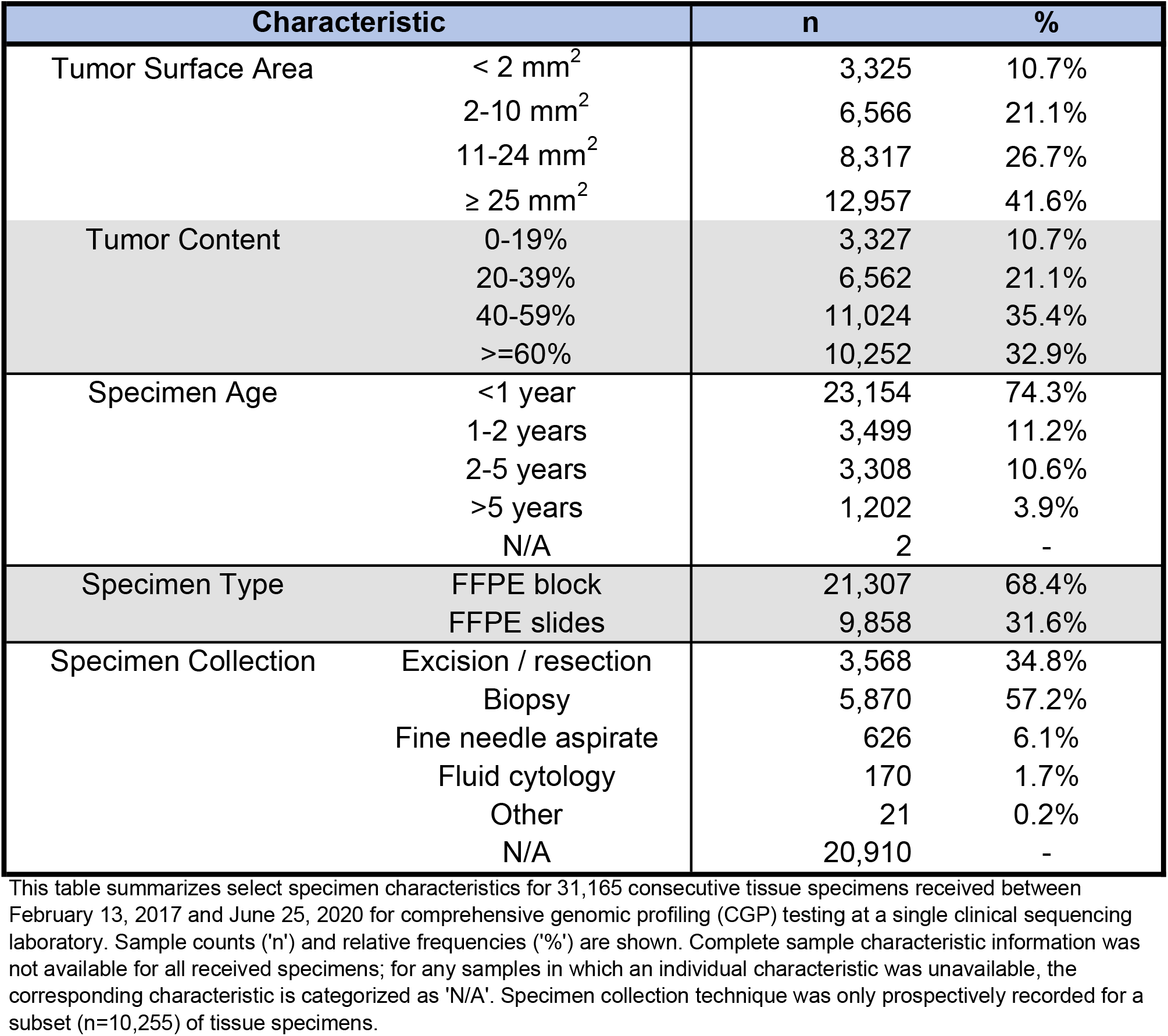
Characteristics of 31,165 specimens received for CGP.

Notably, 10.7% of all tested samples had a final molecularly informed TC less than 20% (**Table 1**). A 20% minimum TC overall requirement is common for CGP tests (including the PCR-CGP test used herein), because corresponding limits of detection (LOD) can preclude exclusion of certain variants/variant types below that TC ^3,5^. Likewise, only 41.6% of samples had TSA ≥ 25mm^2^, with 31.8% of samples having ≤ 10mm^2^ TSA (including 10.7% with < 2mm^2^ TSA) (**Table 1**). Importantly, TSA ≥ 25mm^2^ is the minimum TSA requirement for several leading commercial hybrid capture-based (HC-) CGP tests, including the only FDA approved tissue CGP companion diagnostic device (FoundationOne CDx) ^3,5,27,28^.

As expected, in the consecutive subset of samples where specimen type was prospectively recorded at time of histopathologic review, the majority of samples were from biopsies (57.2%) and cytology cell blocks (fine needle aspirate [6.1%] and fluid cytology [1.7%]) (**Table 1**), which are common in patients with unresectable, advanced and metastatic settings. Although 96.1% of samples received were < 5 years old (**Table 1**), this varied by tumor type, with 14.6% of all tested prostate cancer samples being >5 years old (**Supplementary Table 4**). As expected, nucleic acid yield was associated with TSA. Among samples with ≥2mm^2^ TSA, 90.0% and 96.3% yielded >1 ng/uL DNA and >1 ng/uL RNA, respectively; in contrast, among samples with <2mm^2^ TSA, only 44.5% and 51.7% yielded >1 ng/uL DNA and >1 ng/uL RNA, respectively (**Supplementary Table 1**). In summary, across this consecutive series of real world samples submitted for CGP, we found that TC, TSA, sample age and nucleic acid yield varied widely, with a disproportionate number of minute samples, highlighting the importance of optimizing CGP for limited input material.

### Pan-tumor CGP Experience

Given our previous experience that PCR-CGP could often deliver partial results even in very poor quality samples, CGP was attempted on *all* 31,165 tumor samples using the PCR-CGP test, including “exception” samples not passing input requirements (those with TC < 20%, TSA < 2mm^2^, specimen age > 5 years, or DNA/RNA concentration < 1ng/uL); median turnaround time from sample receipt to report release was 7 business days (interquartile range 6-9 business days). Of these 31,165 samples, 31,101 (99.8%) were evaluable for passing sample input requirements and were further considered for assessing the impact of sample characteristics on PCR-CGP performance and reportability (**Figure 1A**).

As shown in **Figure 1** and **Supplementary Table 5**, in total, 29,302 of 31,101 (94.2%) samples were successfully reported, defined as having at least one reported prioritized alteration or having no prioritized alteration but passing all sequencing QC assessments (as well as the ≥20% TC sample input requirement). Among the 23,022 (74.0%) samples passing all input requirements, 22,791 (99.0%) were successfully reported. As shown in **Figure 1B** and **Supplementary Table 5**, reportability did not vary by sample size, demonstrating that this PCR-CGP test is suitable for minute samples with as little as 2mm^2^ TSA, so long as the other input requirements are met. Notably, among 8,079 (26.0%) exception samples, 6,511 (80.6%) were still successfully reported (**Figure 1A&B**). Samples with TC < 20% made up the largest (10.7%) and poorest performing exception category (68.2% successfully reported), which is expected given that samples with TC < 20% automatically fail QC due to the PCR-CGP test’s overall LOD, and thus all such samples without reported prioritized alterations are deemed test failures (not reported) as the presence of variants cannot be excluded. Notably, however, even in this sub-LOD setting, successful test results were obtained for most samples. Samples failing to meet other input requirements (TSA, sample age, DNA/RNA yield) had decreased reportability rates (86.2% -90.3%) relative to samples passing all requirements, but again, still provided reportable results for most samples (**Figure 1B**).

In the subset of 8,458 consecutively samples tested in the current PCR-CGP configuration during 2020, we assessed the impact of sample characteristics on individual CGP variant class performance. Importantly, this subset was similar to the overall cohort in terms of the proportion of sample exceptions (24.3% vs. 26.0%), overall reportability rate (94.3% vs. 94.2%) and exception sample reportability rate (79.4% vs. 80.6%). In this subset, while sequencing QC pass rates were universally high (95.7%-99.4%) for all CGP variant types in samples meeting input requirements (**Supplementary Table 6**), pass rates were lower in samples not meeting input requirements (68.2%-88.2%%), with gene fusion (74.8% sequencing QC pass) and TMB (68.2% sequencing QC pass) most impacted by sample input characteristics. Importantly, the full sequencing QC pass rate was 92.8% for samples meeting all input requirements. Taken together, across this large, real-world cohort of consecutive tumor samples tested by CGP, PCR-CGP successfully reported results for nearly all samples, including the majority of samples below input requirements. Representative successfully tested samples across the TSA range are depicted in **Figure 1C**.

### Comparison to an External Hybrid Capture CGP cohort

Although previously technically validated as part of the MolDX review process, we further sought to clinically validate the results of our PCR-CGP test through comparison of biomarker frequency distributions in our cohort with those from MSK-IMPACT, an independent, large (*n* > 10,000), tertiary care, academic, single institution experience profiling advanced solid tumors using a hybrid capture CGP test cleared by the FDA ^7^. As shown in **Supplementary Tables 7 and 8**, across 20 major tumor types having at least 80 samples in each cohort, biomarker frequencies were highly correlated (overall Pearson *r* = 0.990; per tumor type r = 0.897-0.999). Importantly, these results also confirm that biomarker frequency rates are similar in the range of real-world tumor samples from our multi-institutional cohort to those from a single academic institution.

### Prostate Carcinoma Experience

Molecular testing to guide therapy in metastatic castration resistant prostate cancer (CRPC) has been limited until recently by a lack of clinical utility, despite the recognition that MSI-H (often somatic) and homologous recombination deficiency (HRD; most commonly by germline/somatic mutation/deep deletion of *BRCA2*) are relatively frequent in CRPC ^29-31^. With the FDA approval of pembrolizumab for all advanced MSI-H solid tumors, and the approval of rucaparib (for *BRCA1/2*) and olaparib (for *BRCA1/2, ATM*, and 11 other potential HRD genes) for CRPC, testing of all men with CRPC for MSI-H and HRD gene alterations are now recommended by the NCCN ^17,25,32,33^.

Hence, we characterized our prostate cancer cohort, limiting to the 1,344 samples tested by the current StrataNGS test version. As shown in **Figure 2A**, although the overall proportion of sample exceptions was similar in prostate cancer to the overall pan-tumor cohort (33.6% vs.%), prostate cancer had the highest frequency of age exception samples (10.8%), consistent with the frequent delay between diagnosis and recurrence after definitive therapy and/or androgen deprivation therapy. Likewise, only 20.4% of samples met input requirements and had TSA ≥ 25mm^2^, suggesting that such tumor requirements are impractical for real world prostate cancer samples where CGP is desired.

**Figure 2.**
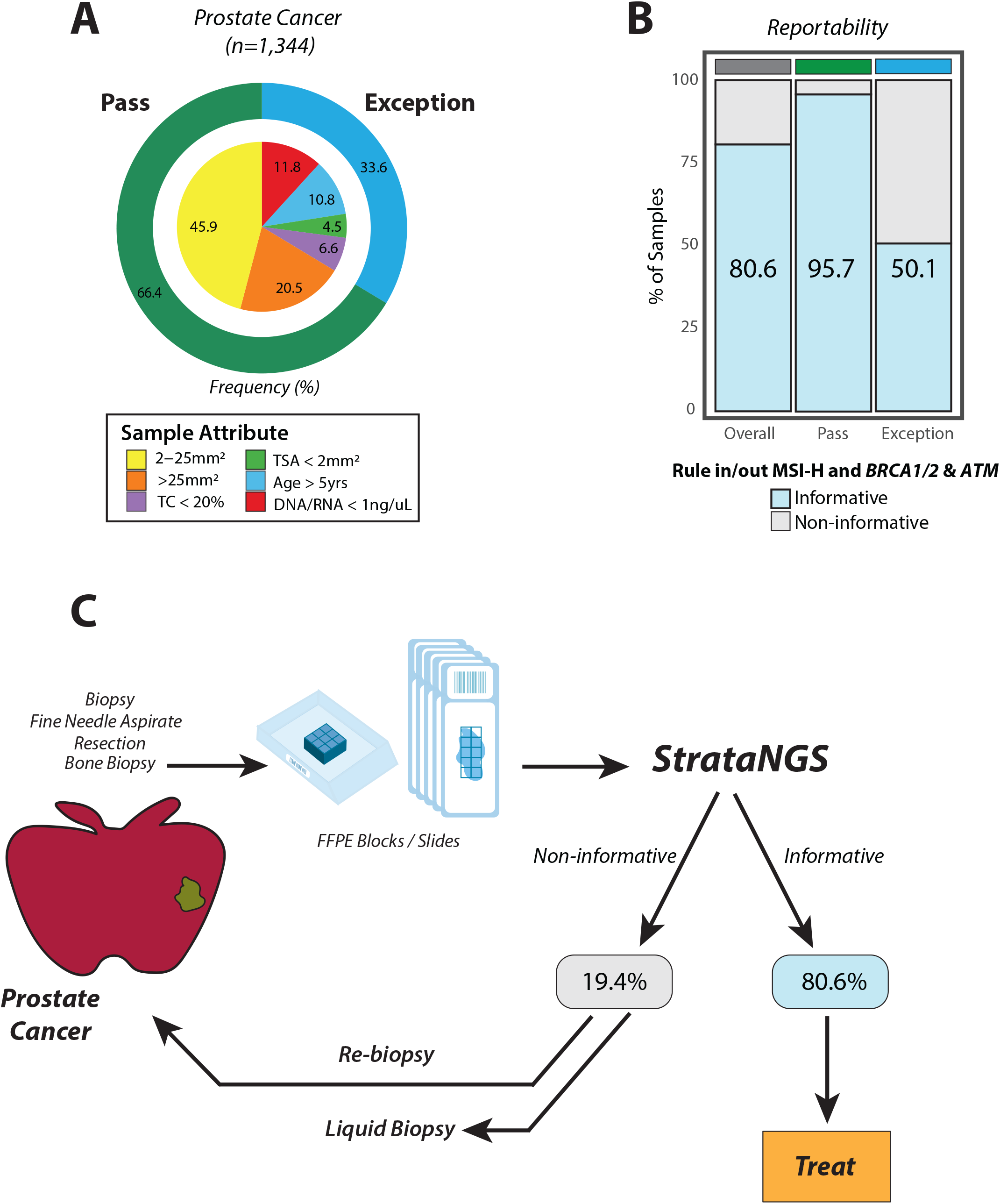
**A)** Donut plot characterizing the composition of consecutively tested, sample input characteristic evaluable prostate cancer samples (n=1,344) from the overall PCR-CGP test cohort. The outer ring indicates the percentage of samples meeting (‘Pass’: green) or not meeting ‘Exception’: cyan) PCR-CGP input requirements. In the inner pie chart, samples passing all input requirements are stratified by tumor surface area (TSA); exception samples are stratified by indicated sample attribute (‘TC < 20%’: tumor content < 20%; TSA < 2mm^2^; ‘Age > 5yrs’: specimen collected > 5 years prior to PCR-CGP; ‘DNA/RNA < 1ng/uL’: DNA and/or RNA concentration < 1ng/uL). **B)** The proportion of tested samples (overall and by sample input requirement category) for which an informative result (able to rule in/our actionable alterations) was reported. To be considered informative, the test must have reported 1) either microsatellite instability high (MSI-H) or a deleterious mutation/copy number deep deletion in *MSH2/6, BRCA1/2*, or *ATM*, or 2) tested definitively negative for these biomarkers by meeting all sequencing QC metrics and having TC ≥20% (the PCR-CGP test’s overall limit of detection). **C)** Real-world prostate cancer testing paradigm based on sample characteristics and PCR-CGP performance characteristics observed in this cohort.

To assess the performance of the PCR-CGP test for therapy selection in prostate cancer, we separated reports into those yielding ‘informative’ therapy selection results (able to rule in/out biomarker guided therapy) for MSI status and HRD gene (*BRCA1, BRCA2* and *ATM*) alterations from those yielding ‘non-informative’ results where additional testing (either by liquid biopsy or obtaining and testing another sample) would be required (see **Methods**). Overall, 80.6% of prostate cancer samples yielded informative results (including 95.7% of samples meeting all input requirements, and 50.7% of exception samples **(Figure 2B & Supplementary Table 9)**. Importantly, the positive MSI-H/HRD biomarker rate was similar between samples meeting all input requirements vs. exception samples (14.8% vs. 12.4%, **Supplementary Table 9**), consistent with most non-informative testing being due to the inability to definitely assert negative results in samples not meeting TC requirements or sequencing QC metrics given the overall low rate of actionable biomarkers in prostate cancer. For example, although 6.8% of <20% TC exception samples were positive for MSI-H/HRD biomarkers (vs. 14.8% of non-exception samples), the remaining 93.2% of <20% TC exception samples were considered non-informative as they cannot exclude the presence of those biomarkers. Nevertheless, our results suggest that ∼80% of all patients with advanced prostate cancer desiring CGP have sufficient tissue samples for informative PCR-CGP, minimizing the need to obtain and test a new sample or pursue liquid biopsy testing (**Figure 2C**).

### NSCLC Adenocarcinoma Experience

In contrast to prostate cancer, biomarker testing is prevalent in NSCLC, and CGP is especially relevant given the large number of recommended biomarkers required to determine appropriate therapy (**Supplementary Table 2**). Therefore, we characterized our NSCLC adenocarcinoma cohort, limiting to the 1,144 NSCLC samples tested by the current StrataNGS test version when NSCLC subtype (adenocarcinoma) was prospectively determined at the time of histopathologic review (**Figure 3**). The proportion of the NSCLC adenocarcinoma cohort not meeting sample input requirements was greater than the overall pan-tumor cohort (40.7% vs. 26.0%), with 21.8% of having TC < 20% and 12.1% having TSA < 2mm^2^ (**Figure 3A & Supplementary Table 10**). Similar to prostate cancer, only 17% of NSCLC adenocarcinoma samples met input requirements and had TSA ≥ 25mm^2^, again demonstrating that this tissue requirement is impractical for real-world samples. To assess the performance of the PCR-CGP test for therapy selection in NSCLC adenocarcinoma, we separated reported results yielding ‘informative’ therapy selection results (able to rule in/out biomarker guided therapy) from those yielding ‘non-informative’ results (see **Methods**). Overall, 87.8% of NSCLC adenocarcinoma samples yielded informative results (**Figure 3B & Supplementary Table 10**), including 98.2% of samples meeting input requirements and 72.5% of exception samples. Importantly, overall informative biomarker frequencies in this NSCLC adenocarcinoma cohort were highly similar to those observed in NSCLC adenocarcinoma from the MSK-IMPACT cohort (Pearson correlation coefficient *r*=0.95, *p* < 0.001; **Figure 3C**).

**Figure 3.**
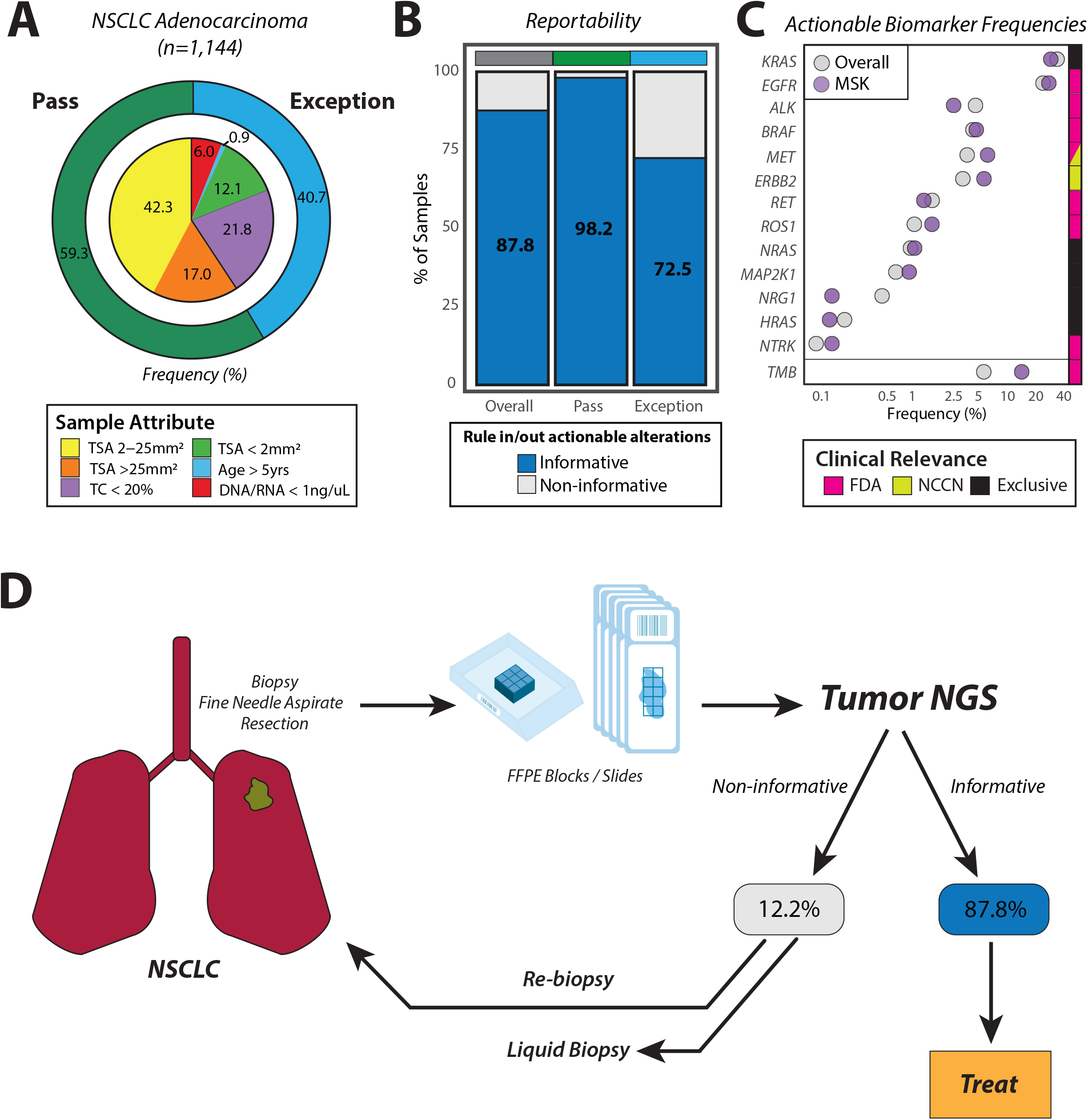
**A)** Donut plot characterizing the composition of consecutively tested, sample input characteristic evaluable non-small cell lung cancer (NSCLC) adenocarcinoma samples (n=1,144) from the overall PCR-CGP test cohort. The outer ring indicates the percentage of samples meeting (‘Pass’: green) or not meeting ‘Exception’: cyan) PCR-CGP input requirements. In the inner pie chart, samples passing all input requirements are stratified by tumor surface area (TSA); exception samples are stratified by indicated sample attribute (‘TC < 20%’: tumor content < 20%; TSA < 2mm^2^; ‘Age > 5yrs’: specimen collected > 5 years prior to PCR-CGP; ‘DNA/RNA < 1ng/uL’: DNA and/or RNA concentration < 1ng/uL). **B)** The proportion of tested samples (overall and by sample input requirement category) for which an informative result (able to rule in/our actionable alterations) was reported. To be considered informative, the test must have either reported a therapy selection/mutually exclusive biomarker (as in **C** and **Supplementary Table 2)** or tested definitively negative for all such biomarkers by meeting all sequencing QC metrics and having TC ≥20% (the PCR-CGP test’s overall limit of detection). **C)** Reported biomarker frequencies from this PCR-CGP NSCLC Adenocarcinoma cohort (overall) are shown along with those from an external single institution cohort (MSK-IMPACT; MSK: purple). The color bar at right indicates whether testing positive for each corresponding biomarker is associated with an FDA-approved (pink) or NCCN-recommended (lime green) targeted therapy or thought to be mutual exclusive with known LUAD therapy selection biomarkers. Tumor mutation burden (TMB) frequencies are presented separately from gene-specific biomarkers given the expected overlap between TMB-High and some therapy selection/actionable biomarkers. **D)** Real-world NSCLC Adenocarcinoma testing paradigm based on sample characteristics and PCR-CGP performance characteristics observed in this cohort.

Like prostate cancer, NSCLC adenocarcinoma samples with TC < 20% had the lowest informative result rate, as negative results cannot be definitively asserted in this sub-LOD setting (**Supplementary Table 10**). However, in contrast to prostate cancer, where the overall actionable biomarker frequency is low, actionable/informative biomarkers are frequent in NSCLC adenocarcinoma. Hence, the positive informative biomarker detection rate in NSCLC adenocarcinoma TC exception samples of 57.8% is only marginally lower than the positive detection rate in samples meeting input requirements (79.1%), and all other sample exception groups had positive detection rates of 79.0-81.2% (**Supplementary Table 10**). Importantly, these results suggest that the majority of positive biomarkers can be identified even in the most challenging NSCLC adenocarcinoma samples, and ∼88% of patients with advanced NSCLC adenocarcinoma desiring CGP have sufficient tissue samples for informative PCR-CGP (**Figure 3C**).

## Discussion

Herein we present the tissue characteristics and PCR-CGP test performance from over 30,000 consecutively tested solid tumor samples from patients with advanced cancer collected from 28 diverse health systems across the U.S. through a multi-institutional observational clinical trial (NCT03061305). Importantly, sites were provided with minimal requirements to guide sample submission (1 block or 10 x 5um slides with ≥ 2mm^2^ TSA and ≥20% TC), and PCR-CGP testing was attempted for essentially all samples with identifiable tumor, providing a unique view on real-world tumor tissue availability and CGP test performance.

Unexpectedly, we found that a large proportion of available samples were limited, with 10.7% having TC < 20% and 44.7% having TSA ≤10mm^2^ (including 7.0% with < 2mm^2^ TSA). Despite these challenges, PCR-CGP reported results for 94.2% of all tested tumor samples, including 99.0% of samples meeting all PCR-CGP sample input requirements (≥ 2mm^2^ TSA, ≥20% TC, sample age < 5 yrs and DNA/RNA yield ≥1ng/ul) and 80.6% of exception samples not meeting those criteria. Specifically, among NSCLC adenocarcinoma, where CGP is especially important for treatment selection, we found that limited tissue was even more pronounced with 21.8% of samples having < 20% TC and 12.1% having < 2 mm^2^ TSA. Despite these limitations, PCR-CGP testing successfully reported results for 93.6% of all samples, including results informative for treatment selection in 87.8% of samples, obviating the need to obtain additional tissue or pursue liquid biopsy. Similar results were observed in prostate carcinoma, where despite a much lower rate of positive informative biomarkers, PCR-CGP testing yielded results informative for treatment selection in 80.6% of all tested samples.

We attribute the high reportability rates of PCR-CGP to three main factors. First, the CGP test utilized a multiplex PCR-based library preparation method, in contrast to other large cohort studies, which have all utilized hybrid capture-based library preparation (HC-) approaches requiring significantly higher nucleic acid input and thus, tumor size, input requirements. For example, while the PCR-CGP test evaluated herein requires minimum tumor surface area ≥2mm^2^, other leading commercially available HC-CGP tests require ≥25mm^2 3,5,27,28^. Notably, only 40.8% of samples in our total cohort met this requirement and among lung NSCLC samples the proportion was even smaller (23.6%, **Supplementary Table 4**). We suspect that this HC-CGP input requirement may be significantly limiting the uptake of tissue-based CGP, with may samples never being submitted for CGP or being returned/reported as quantity not sufficient. Consistent with these findings, in clinical trials testing available FFPE tissue samples from patients with advanced NSCLC or prostate cancer, HC-CGP failure rates of ∼30-40% have been reported^16-19^. Thus, the PCR-CGP test, with its lower input requirements, has the potential to expand the proportion of testable tumor samples.

Second, given the ability of PCR based CGP to generate some data on nearly all samples and our belief that even a single biomarker may be highly actionable (regardless of the ability to assess other CGP variant classes), we employed a liberal exception testing policy, where we attempted testing on nearly all samples that had any identifiable tumor, even if the sample did not meet one or more of our input requirements. This approach necessitated PCR-CGP sequencing QC metrics optimized for minute, low quality samples, and included variant level review. Hence, although not all CGP variant classes may be assessable in all samples, this approach seeks to maximize actionable insights from the available tissue. While only 74% of samples met all input requirements, reported results were delivered for 94.2% of all tested samples. As expected, reportability rates were higher in samples meeting all input requirements (99.0%) relative to samples not meeting one or more requirements (80.6%). Low tumor content samples were the most challenging (68.2% reportability), given the inability to exclude the presence of alterations in such samples based on test LOD; hence, future CGP improvements should focus on improving the limit of detection down to 5-10% TC for all variant classes.

To further improve the reportability rate, CGP can be performed ahead of potentially redundant single marker testing, which can rapidly exhaust tissue. Additionally, when possible, additional tissue specifically collected for molecular analysis could be considered for all advanced tumors, similar to NCCN recommendations for NSCLC ^34^. Lastly, liquid biopsy represents an alternative CGP methodology when no tissue is available or is difficult to procure. While liquid biopsy can produce highly specific results for treatment selection^16^, as evidence by the recent FDA approval of both the FoundationOne Liquid CDx and Guardant360 CDx cfDNA CGP tests, sensitivity can be challenged by the lack of circulating tumor DNA in some patients and the difficulty of differentiating between an informative negative test and a lack of detectable cfDNA (e.g. does a cfDNA test in a patient with NSCLC identifying a TP53 mutation at 0.5% variant allele frequency [VAF] exclude the possibility of an EGFR exon 19 deletion in the sample) given the prevalence of clonal hematopoiesis and the poor positive predictive value of many de novo alterations at VAFs <1% ^35-38^. For example, the Guardant360 CDx test showed only 67.4-77.7% positive predictive agreement compared to tissue based testing for EGFR exon 19 deletions and p.L858R and p.T790M mutations^37^, while the Guardant360 laboratory developed test similarly had only 61% sensitivity compared to tissue based targeted NGS for detecting *ALK* resistance mutations in patients progressing on ALK inhibitors ^39^.

Taken together, with the growing compendium of biomarker-guided targeted and immunotherapies, the importance of CGP for treatment selection in patients with advanced cancer is clear. Our study demonstrates that the majority of patients desiring CGP have challenging tissue specimens, optimized approaches including PCR-CGP and a broad sample exception testing approach can maximize the actionable information from each received tissue sample. Future developments should seek to further improve CGP testing technology and the routine collection of sufficient tissue specifically for CGP.

## Data Availability

: De-identified subject and molecular data is made available to all Strata Trial partnered institutions.

## Author Contributions

Drs. Tomlins and Hovelson had full access to all the data in the study and take responsibility for the integrity of the data and the accuracy of the data analysis.

Concept and design: Drs. Rhodes and Tomlins; Johnson and Kwiatkowski.

Acquisition, analysis, or interpretation of data: All Authors.

Drafting of the manuscript: Drs. Tomlins, Hovelson, and Rhodes.

Critical revision of the manuscript for important intellectual content: All Authors.

Statistical analysis: Hovelson, Tomlins.

Administrative, technical, or material support: Drewery, Dr. Falkner, Fischer, Dr. Hipp, Kwiatkowski, Dr. Lazo de la Vega, Mitchell, Reeder, Siddiqui, and Dr. Vakil.

Supervision: Drs. Rhodes and Tomlins.

## Meeting Presentation

Partial results from this study were presented at the American Society of Clinical Oncology 2020 Annual Meeting; Chicago, IL.

## Data Sharing Statement

De-identified subject and molecular data is made available to all Strata Trial partnered institutions.

## References

1. Cheng ML, Berger MF, Hyman DM, et al: Clinical tumour sequencing for precision oncology: time for a universal strategy. Nat Rev Cancer 18:527–528, 2018

2. Beaubier N, Bontrager M, Huether R, et al: Integrated genomic profiling expands clinical options for patients with cancer. Nat Biotechnol 37:1351–1360, 2019

3. Frampton GM, Fichtenholtz A, Otto GA, et al: Development and validation of a clinical cancer genomic profiling test based on massively parallel DNA sequencing. Nat Biotechnol 31:1023–31, 2013

4. Robinson DR, Wu YM, Lonigro RJ, et al: Integrative clinical genomics of metastatic cancer. Nature 548:297–303, 2017

5. Vanderwalde A, Spetzler D, Xiao N, et al: Microsatellite instability status determined by next-generation sequencing and compared with PD-L1 and tumor mutational burden in 11,348 patients. Cancer Med 7:746–756, 2018

6. Wood DE, White JR, Georgiadis A, et al: A machine learning approach for somatic mutation discovery. Sci Transl Med 10, 2018

7. Zehir A, Benayed R, Shah RH, et al: Mutational landscape of metastatic cancer revealed from prospective clinical sequencing of 10,000 patients. Nat Med 23:703–713, 2017

8. Dy GK, Nesline MK, Papanicolau-Sengos A, et al: Treatment recommendations to cancer patients in the context of FDA guidance for next generation sequencing. BMC Med Inform Decis Mak 19:14, 2019

9. Morris SM, Subramanian J, Gel ES, et al: Performance of next-generation sequencing on small tumor specimens and/or low tumor content samples using a commercially available platform. PLoS One 13:e0196556, 2018

10. Sholl LM, Do K, Shivdasani P, et al: Institutional implementation of clinical tumor profiling on an unselected cancer population. JCI Insight 1:e87062, 2016

11. Woodhouse R, Li M, Hughes J, et al: Clinical and analytical validation of FoundationOne Liquid CDx, a novel 324-Gene cfDNA-based comprehensive genomic profiling assay for cancers of solid tumor origin. PLoS One 15:e0237802, 2020

12. Gandara DR, Paul SM, Kowanetz M, et al: Blood-based tumor mutational burden as a predictor of clinical benefit in non-small-cell lung cancer patients treated with atezolizumab. Nat Med 24:1441–1448, 2018

13. Odegaard JI, Vincent JJ, Mortimer S, et al: Validation of a Plasma-Based Comprehensive Cancer Genotyping Assay Utilizing Orthogonal Tissue- and Plasma-Based Methodologies. Clin Cancer Res 24:3539–3549, 2018

14. Georgiadis A, Durham JN, Keefer LA, et al: Noninvasive Detection of Microsatellite Instability and High Tumor Mutation Burden in Cancer Patients Treated with PD-1 Blockade. Clin Cancer Res 25:7024–7034, 2019

15. Brown NA, Elenitoba-Johnson KSJ: Enabling Precision Oncology Through Precision Diagnostics. Annu Rev Pathol 15:97–121, 2020

16. Aggarwal C, Thompson JC, Black TA, et al: Clinical Implications of Plasma-Based Genotyping With the Delivery of Personalized Therapy in Metastatic Non-Small Cell Lung Cancer. JAMA Oncol 5:173–180, 2019

17. de Bono J, Mateo J, Fizazi K, et al: Olaparib for Metastatic Castration-Resistant Prostate Cancer. N Engl J Med 382:2091–2102, 2020

18. Gilson C, Ingleby F, Gilbert DC, et al: Targeted next-generation sequencing (tNGS) of metastatic castrate-sensitive prostate cancer (M1 CSPC): A pilot molecular analysis in the STAMPEDE multi-center clinical trial. Journal of Clinical Oncology 37:5019–5019, 2019

19. Hellmann MD, Ciuleanu TE, Pluzanski A, et al: Nivolumab plus Ipilimumab in Lung Cancer with a High Tumor Mutational Burden. N Engl J Med 378:2093–2104, 2018

20. File DM, Morgan KP, Khagi S: Durable Near-Complete Response to Olaparib Plus Temozolomide and Radiation in a Patient With ATM-Mutated Glioblastoma and MSH6-Deficient Lynch Syndrome. JCO Precision Oncology:841–847, 2020

21. Miller TI, Zoumberos NA, Johnson B, et al: A genomic survey of sarcomas on sun-exposed skin reveals distinctive candidate drivers and potentially targetable mutations. Hum Pathol 102:60–69, 2020

22. Cerami E, Gao J, Dogrusoz U, et al: The cBio cancer genomics portal: an open platform for exploring multidimensional cancer genomics data. Cancer Discov 2:401–4, 2012

23. Mandelker D, Donoghue M, Talukdar S, et al: Germline-focussed analysis of tumour-only sequencing: recommendations from the ESMO Precision Medicine Working Group. Ann Oncol 30:1221–1231, 2019

24. McKinney W: Data structures for statistical computing in python, Proceedings of the 9th Python in Science Conference. Austin, TX, 2010, pp 51–56

25. Network NCC: Prostate Cancer (Version 2.2020), 2020

26. Jordan EJ, Kim HR, Arcila ME, et al: Prospective Comprehensive Molecular Characterization of Lung Adenocarcinomas for Efficient Patient Matching to Approved and Emerging Therapies. Cancer Discov 7:596–609, 2017

27. Medicine F: FoundationOne CDx Specimen Instructions, 2020

28. Caris: Caris MI Molecular Intelligence Specimen Preparation Instructions, 2020

29. Grasso CS, Wu YM, Robinson DR, et al: The mutational landscape of lethal castration-resistant prostate cancer. Nature 487:239–43, 2012

30. Kumar A, White TA, MacKenzie AP, et al: Exome sequencing identifies a spectrum of mutation frequencies in advanced and lethal prostate cancers. Proc Natl Acad Sci U S A 108:17087–92, 2011

31. Robinson D, Van Allen EM, Wu YM, et al: Integrative Clinical Genomics of Advanced Prostate Cancer. Cell 162:454, 2015

32. Abida W, Patnaik A, Campbell D, et al: Rucaparib in Men With Metastatic Castration-Resistant Prostate Cancer Harboring a BRCA1 or BRCA2 Gene Alteration. J Clin Oncol:JCO2001035, 2020

33. Le DT, Durham JN, Smith KN, et al: Mismatch repair deficiency predicts response of solid tumors to PD-1 blockade. Science 357:409–413, 2017

34. Network NCC: Non-Small Cell Lung Cancer (version 8.2020), 2020

35. Stetson D, Ahmed A, Xu X, et al: Orthogonal Comparison of Four Plasma NGS Tests With Tumor Suggests Technical Factors are a Major Source of Assay Discordance. JCO Precision Oncology:1–9, 2019

36. Razavi P, Li BT, Brown DN, et al: High-intensity sequencing reveals the sources of plasma circulating cell-free DNA variants. Nat Med 25:1928–1937, 2019

37. Administration FaD: PMA P200010: FDA Summary of Safety and Effectiveness Data, 2020

38. Adminsitration FaD: PMA P190032: FDA Summary of Safety and Effectiveness Data, 2020

39. Shaw AT, Solomon BJ, Besse B, et al: ALK Resistance Mutations and Efficacy of Lorlatinib in Advanced Anaplastic Lymphoma Kinase-Positive Non-Small-Cell Lung Cancer. J Clin Oncol 37:1370–1379, 2019

