## Supplementary Tables 1-10 for "Comprehensive genomic profiling of 30,000 consecutive solid tumors"

### SUPPLEMENTARY INFORMATION

#### Supplementary Tables 1-10

### Supplementary Table 1 - Nucleic acid concentrations (by specimen sample size)

| | | TSA < 2 $mm^2$ | | TSA >= 2 $mm^2$ | | Total | |
|---|---|---|---|---|---|---|---|
| **Nucleic Acid** | **Bin** | **n** | **%** | **n** | **%** | **n** | **%** |
| **RNA** | < 1 ng/uL | 1,029 | 48.3% | 1,025 | 3.7% | 2,054 | 6.9% |
| | >= 1 ng/uL | 1,103 | 51.7% | 26,636 | 96.3% | 27,739 | 93.1% |
| **DNA** | < 1 ng/uL | 1,183 | 55.5% | 2,772 | 10.0% | 3,955 | 13.3% |
| | >=1 ng/uL | 949 | 44.5% | 24,889 | 90.0% | 25,838 | 86.7% |
| | ***Total*** | ***2,132*** | ***100.0%*** | ***27,661*** | ***100.0%*** | ***29,793*** | ***100.0%*** |

This table presents sample counts/frequencies from the overall PCR-CGP cohort broken down by nucleic acid concentration and specimen tumor surface area (TSA). Results presented here were restricted to analysis of samples with complete DNA/RNA concentration and specimen TSA info (n=29,793; 95.6% of total n=31,165). Sample counts and frequencies were calculated separately using DNA- and RNA-based nucleic acid concentrations, and both sets of summaries are presented here for comparison.

### Supplementary Table 2 - Informative/Actionable NSCLC adenocarcinoma biomarkers

| **Gene / Biomarker** | **Variant Types** | **Relevance** |
|---|---|---|
| *ALK* | Fusion | FDA |
| *BRAF* | Fusion, INDEL, SNV | FDA |
| *EGFR* | INDEL, SNV | FDA |
| *ERBB2* | CNV, INDEL, SNV | NCCN |
| *HRAS* | SNV | Exclusive |
| *KRAS* | SNV | Exclusive |
| *MAP2K1* | INDEL, SNV | Exclusive |
| *MET* | CNV, Fusion, INDEL, SNV | FDA, NCCN |
| *NRAS* | SNV | Exclusive |
| *NRG1* | Fusion | Exclusive |
| *NTRK (NTRK3)* | Fusion | FDA |
| *RET* | Fusion | FDA |
| *ROS1* | Fusion | FDA |
| TMB | TMB | FDA, NCCN |
| **Total** | | |

Clinically relevant biomarkers included here are those where, for patients with lung NSCLC adenocarcinoma, a positive test result is (a) associated with an FDA-approved and/or NCCN-recommended therapy (e.g., a therapy selection biomarker); or (b) thought to be mutually exclusive with known therapy selection biomarkers. Gene/biomarker labels associated with each indicated biomarker are listed in the first column, with pertinent PCR-CGP variant type(s) associated with each biomarker listed in the *'Variant Types'* column. The *'Relevance'* column indicates whether a given biomarker is associated with any FDA-approved therapy ('FDA'), NCCN therapy recommendation ('NCCN'), thought to be mutually exclusive with FDA-approved/NCCN-recommended therapy selection biomarkers ('Exclusive'), or a combination of these categories.

**Supplementary Table 3 - Patient Characteristics**

| Characteristic | Attribute | n | % |
|---|---|---|---|
| **Gender** | Female | 16,493 | 54.0% |
| | Male | 14,072 | 46.0% |
| **Age** | 18-29 | 275 | 0.9% |
| | 30-49 | 3,193 | 10.4% |
| | 50-69 | 14,792 | 48.4% |
| | 70-89 | 12,302 | 40.3% |
| | N/A | 3 | - |
| **Race** | White | 15,146 | 73.4% |
| | Black or African American | 2,501 | 12.1% |
| | Asian | 1,406 | 6.8% |
| | Native Hawaiian or Pacific Islander | 65 | 0.3% |
| | American Indian or Alaska Native | 68 | 0.3% |
| | Other | 1,452 | 7.0% |
| | N/A | 9,927 | - |
| ***Total*** | | ***30,565*** | ***100.0%*** |

This table summarizes select patient characteristics for the 30,565 unique patients associated with 31,165 consecutive tissue specimens received for PCR-CGP testing. Any patients for which corresponding characteristic info was unavailable are categorized as 'N/A' for the indicated characteristic. Percentages displayed ('%') reflect the frequency of patients assigned to each sub-group (e.g., attribute) within the corresponding patient characteristic. Submission of race information was not required during the study period.

**Supplementary Table 4. Characteristics of 31,165 specimens received for CGP by Cancer Type**

| Cancer Type | | | Tumor Surface Area | | | | Tumor Content | | | | Sample Age (yrs) | | | |
|---|---|---|---|---|---|---|---|---|---|---|---|---|---|---|
| **Cancer Type** | **Total *n*** | **%** | **<2 mm$^2$** | **2-9 mm$^2$** | **10-24 mm$^2$** | **≥25 mm$^2$** | **0-19%** | **20-39%** | **40-59%** | **≥60%** | **<1** | **1-2** | **2-5** | **>5** |
| **Lung - NSCLC** | **4457** | **14.3%** | 13.9% | 50.9% | 11.6% | 23.6% | 18.4% | 31.1% | 28.5% | 22.0% | 86.2% | 7.2% | 5.5% | 1.2% |
| **Breast** | **3713** | **11.9%** | 5.7% | 36.0% | 20.5% | 37.8% | 7.0% | 16.3% | 27.7% | 49.0% | 64.2% | 12.3% | 16.5% | 7.0% |
| **Colon and Rectum** | **3700** | **11.9%** | 5.2% | 38.4% | 12.3% | 44.0% | 7.2% | 23.0% | 35.6% | 34.2% | 71.8% | 13.9% | 12.2% | 2.2% |
| **Prostate** | **2045** | **6.6%** | 4.5% | 42.4% | 23.9% | 29.1% | 6.1% | 19.5% | 32.6% | 41.8% | 53.2% | 12.8% | 19.5% | 14.6% |
| **Pancreas** | **1955** | **6.3%** | 12.4% | 46.1% | 14.2% | 27.2% | 29.5% | 34.0% | 22.4% | 14.1% | 82.8% | 10.4% | 5.6% | 1.3% |
| **Ovary** | **1857** | **6.0%** | 2.4% | 20.0% | 10.3% | 67.2% | 8.7% | 16.2% | 24.3% | 50.7% | 58.6% | 16.1% | 19.3% | 6.0% |
| **Unknown Primary** | **1513** | **4.9%** | 13.8% | 51.7% | 14.9% | 19.6% | 14.3% | 22.2% | 27.5% | 36.0% | 92.7% | 4.2% | 2.6% | 0.5% |
| **Other** | **1500** | **4.8%** | 3.1% | 31.2% | 14.8% | 50.9% | 8.3% | 16.0% | 22.9% | 52.8% | 71.2% | 12.1% | 12.3% | 4.4% |
| **Brain** | **1421** | **4.6%** | 1.4% | 11.1% | 7.7% | 79.7% | 3.0% | 8.2% | 14.7% | 74.1% | 71.8% | 11.0% | 10.3% | 6.8% |
| **Endometrium** | **1099** | **3.5%** | 4.0% | 22.4% | 10.1% | 63.5% | 5.7% | 17.0% | 25.7% | 51.6% | 66.7% | 16.3% | 14.1% | 2.9% |
| **Sarcoma** | **912** | **2.9%** | 3.0% | 22.5% | 13.7% | 60.9% | 3.0% | 8.1% | 13.6% | 75.3% | 77.5% | 10.0% | 9.4% | 3.1% |
| **Head and Neck** | **902** | **2.9%** | 4.8% | 31.0% | 16.8% | 47.5% | 5.1% | 17.4% | 30.7% | 46.8% | 78.8% | 12.1% | 7.1% | 2.0% |
| **Melanoma** | **843** | **2.7%** | 6.2% | 32.0% | 16.1% | 45.7% | 6.9% | 11.7% | 18.7% | 62.6% | 84.0% | 7.9% | 7.0% | 1.1% |
| **Esophagus** | **789** | **2.5%** | 6.6% | 67.5% | 13.3% | 12.6% | 8.1% | 23.8% | 34.9% | 33.2% | 85.8% | 9.3% | 4.8% | 0.1% |
| **Bladder** | **734** | **2.4%** | 6.4% | 27.1% | 11.2% | 55.3% | 5.9% | 16.5% | 30.0% | 47.7% | 81.1% | 11.9% | 5.9% | 1.2% |
| **Kidney** | **678** | **2.2%** | 6.2% | 29.6% | 9.2% | 55.0% | 6.0% | 24.3% | 25.4% | 44.2% | 68.1% | 12.1% | 14.9% | 4.9% |
| **Biliary** | **633** | **2.0%** | 6.6% | 43.0% | 20.5% | 29.9% | 19.3% | 34.0% | 27.0% | 19.7% | 82.9% | 11.5% | 4.9% | 0.6% |
| **Stomach** | **574** | **1.8%** | 8.2% | 55.2% | 12.5% | 24.0% | 23.9% | 37.5% | 21.4% | 17.2% | 83.8% | 9.6% | 5.7% | 0.9% |
| **Lung - SCLC** | **533** | **1.7%** | 6.2% | 48.9% | 17.9% | 27.1% | 6.4% | 9.9% | 11.8% | 71.9% | 86.3% | 9.9% | 3.6% | 0.2% |
| **Thyroid** | **330** | **1.1%** | 3.3% | 19.7% | 11.8% | 65.2% | 6.1% | 9.7% | 20.3% | 63.9% | 61.2% | 12.1% | 14.8% | 11.8% |
| **Lymphoma** | **255** | **0.8%** | 6.4% | 32.0% | 21.2% | 40.4% | 14.1% | 16.5% | 17.6% | 51.8% | 72.5% | 15.3% | 8.6% | 3.5% |
| **Cervical** | **175** | **0.6%** | 3.4% | 39.4% | 14.9% | 42.3% | 6.3% | 13.7% | 35.4% | 44.6% | 73.7% | 15.4% | 8.6% | 2.3% |
| **Liver** | **154** | **0.5%** | 6.6% | 44.1% | 18.4% | 30.9% | 1.9% | 18.2% | 24.0% | 55.8% | 77.9% | 12.3% | 7.8% | 1.9% |
| **Non-Melanoma Skin** | **151** | **0.5%** | 2.0% | 31.1% | 15.2% | 51.7% | 2.6% | 15.9% | 28.5% | 53.0% | 84.1% | 11.3% | 3.3% | 1.3% |
| **Small Intestine** | **104** | **0.3%** | 3.8% | 36.5% | 9.6% | 50.0% | 13.5% | 27.9% | 33.7% | 25.0% | 77.9% | 11.5% | 10.6% | 0.0% |
| **Thymus** | **52** | **0.2%** | 7.7% | 32.7% | 17.3% | 42.3% | 13.5% | 11.5% | 32.7% | 42.3% | 75.0% | 9.6% | 11.5% | 3.8% |
| **Meninges** | **46** | **0.1%** | 0.0% | 6.5% | 6.5% | 87.0% | 0.0% | 2.2% | 4.3% | 93.5% | 58.7% | 8.7% | 19.6% | 13.0% |
| **Adrenal** | **29** | **0.1%** | 0.0% | 31.0% | 17.2% | 51.7% | 3.4% | 0.0% | 10.3% | 86.2% | 89.7% | 0.0% | 10.3% | 0.0% |
| **Lung** | **10** | **0.0%** | 33.3% | 50.0% | 0.0% | 16.7% | 10.0% | 30.0% | 30.0% | 30.0% | 70.0% | 10.0% | 10.0% | 10.0% |
| **NSCLC** | **1** | **0.0%** | 100.0% | 0.0% | 0.0% | 0.0% | 0.0% | 0.0% | 0.0% | 100.0% | 100.0% | 0.0% | 0.0% | 0.0% |
| ***Total*** | ***31,165*** | ***100.0%*** | ***7.0%*** | ***37.7%*** | ***14.5%*** | ***40.8%*** | ***10.7%*** | ***21.1%*** | ***26.7%*** | ***41.6%*** | ***74.3%*** | ***11.2%*** | ***10.6%*** | ***3.9%*** |

This table summarizes count and frequency of tumor specimens in the overall PCR-CGP test cohort (n=31,165) by cancer type. For all samples from each cancer type, the relative frequency of the indicated sample characteristic groups are shown.

**Supplementary Table 5: PCR-CGP Reportability by Sample Input Requirement Characteristics**

| Specimen | | Overall | | Reported | | Test Failure | |
|---|---|---|---|---|---|---|---|
| **Requirement Status** | **Attribute** | ***n*** | **%** | ***n*** | **%** | ***n*** | **%** |
| **Pass** | TSA 2-10 $mm^2$ | 7,944 | 25.5% | 7,869 | 99.1% | 75 | 0.9% |
| | TSA 11-24 $mm^2$ | 3,814 | 12.3% | 3,774 | 99.0% | 40 | 1.0% |
| | TSA >= 25 $mm^2$ | 11,264 | 36.2% | 11,148 | 99.0% | 116 | 1.0% |
| | **Subtotal** | **23,022** | **74.0%** | **22,791** | **99.0%** | **231** | **1.0%** |
| **Exception** | TC < 20% | 3,327 | 10.7% | 2,270 | 68.2% | 1,057 | 31.8% |
| | TSA < 2 $mm^2$ | 1,396 | 4.5% | 1,260 | 90.3% | 136 | 9.7% |
| | Age > 5 yrs | 1,090 | 3.5% | 940 | 86.2% | 150 | 13.8% |
| | DNA or RNA < 1 ng/uL | 2,266 | 7.3% | 2,041 | 90.1% | 225 | 9.9% |
| | **Subtotal** | **8,079** | **26.0%** | **6,511** | **80.6%** | **1,568** | **19.4%** |
| | ***Total*** | ***31,101*** | ***100.0%*** | ***29,302*** | ***94.2%*** | ***1,799*** | ***5.8%*** |

Sample counts and PCR-CGP test reportability in 31,101 consecutively tested tissue specimens evaluable for meeting sample input requirements are shown. Overall samples counts (n) and frequencies (%) are stratified by sample requirement status (Pass/Exception) and indicated specimen attribute groupings. Sample requirement status indicates whether a given sample met all minimum PCR-CGP test requirements ('Pass': tumor surface area (TSA) >= 2$mm^2$, tumor content (TC) >= 20%, sample age <= 5 years, and DNA & RNA concentration >= 1ng/uL), or failed to meet at least one of these requirements ('Exception'). For 'Pass' samples, reportability results are stratified by tumor surface area (TSA); for 'Exception' samples, individual samples are grouped by the indicated attribute rules in the following order: (1) TC < 20%; (2) TSA < 2$mm^2$; (3) sample age > 5 years; then, (4) DNA or RNA concentration < 1ng/uL. The overall cohort was then stratified by successfully reported tests or test failures. Tests were considered successfully reported if at least one prioritized alteration was reported *OR* the sample passed all sequencing quality control (QC) metrics and had TC >= 20%.

**Supplementary Table 6 - Sequencing QC (by sample requirement status and specimen attributes)**

| Specimen | | Overall | | | | CGP Variant Class QC Pass | | | | |
|---|---|---|---|---|---|---|---|---|---|---|
| QC Status | Attribute | *n* | % | Reported (*n* = 7,975) | Full QC Pass (*n* = 6,381) | Mutation (*n* = 8,130) | CNA (*n* = 8,180) | Fusion (*n* = 7,712) | MSI (*n* = 8,118) | TMB (*n* = 7,531) |
| **Pass** | TSA 2-10 $mm^2$ | 2,622 | 31.0% | **99.5%** | **91.9%** | 99.3% | 99.5% | 96.6% | 99.1% | 94.7% |
| | TSA 11-24 $mm^2$ | 995 | 11.8% | **99.3%** | **92.7%** | 99.2% | 99.3% | 96.5% | 99.1% | 95.3% |
| | TSA >= 25 $mm^2$ | 2,782 | 32.9% | **98.7%** | **93.6%** | 99.1% | 99.5% | 96.3% | 99.1% | 96.9% |
| | **Subtotal** | **6,399** | **75.7%** | **99.1%** | **92.8%** | **99.2%** | **99.4%** | **96.4%** | **99.1%** | **95.7%** |
| **Exception** | TC < 20%* | 947 | 11.2% | **69.8%** | **0.0%** | 90.1% | 90.2% | 78.0% | 89.8% | 75.1% |
| | TSA < 2 mm2 | 495 | 5.9% | **90.1%** | **44.4%** | 86.1% | 86.3% | 72.1% | 86.5% | 62.6% |
| | Age > 5 yrs | 263 | 3.1% | **82.5%** | **50.6%** | 82.5% | 88.2% | 67.7% | 82.1% | 70.3% |
| | DNA or RNA < 1 ng/uL | 354 | 4.2% | **87.6%** | **26.0%** | 80.5% | 85.9% | 75.4% | 79.9% | 55.9% |
| | **Subtotal** | **2,059** | **24.3%** | **79.4%** | **21.6%** | **86.5%** | **88.2%** | **74.8%** | **86.3%** | **68.2%** |
| | **Total** | **8,458** | **100.0%** | **94.3%** | **75.4%** | **96.1%** | **96.7%** | **91.2%** | **96.0%** | **89.0%** |

This table summarizes sequencing quality control (QC) performance across a subset of samples from the overall PCR-CGP sequencing cohort profiled between January 1, 2020 and June 25, 2020 (n=8,458). Sequencing QC performance is summarized by sample requirement status (Pass/Exception) and indicated specimen attribute groupings, with frequency of successfully reported samples presented separately for each requirement/attribute group. Here, sample requirement status indicates whether a given sample met all minimum PCR-CGP test requirements ('Pass'; minimum PCR-CGP test requirements include: tumor surface area (TSA) >= 2mm2, tumor content (TC) >= 20%, sample age <= 5 years, and DNA & RNA concentration >= 1ng/uL), or failed to meet at least one of these requirements ('Exception'). For 'Pass' samples, sequencing QC (as well as reportability) results are broken down by TSA; for 'Exception' samples, individual samples are grouped by the indicated attribute rules in the following order: (1) TC < 20%; (2) TSA < 2mm2; (3) sample age > 5 years; then, (4) DNA or RNA concentration < 1ng/uL. For each indicated requirement/attribute combination, sample counts and relative contribution ('%') to the sample set is presented in the first 2 columns of this table's *'Overall'* section. Within the *'Overall'* section, the *'Reported'* column indicates the percentage of samples in the corresponding requirement/attribute bucket with a successfully reported test. Tests were considered successfully reported if at least one prioritized alteration was reported OR the sample passed all sequencing quality control (QC) metrics and had TC >= 20%. The `*Full QC Pass*` column indicates the percentage of samples in the requirement/attribute bucket that passed all PCR-CGP variant class-specific sequencing QC assessment checks (includes QC checks specific to: point mutations or short insertions/deletions (Mutation); copy-number alterations (CNA); gene fusions (Fusion); microsatellite instability (MSI); and tumor mutation burden (TMB)). Within the *'CGP Variant Class'* section of the table, variant class-specific sequencing QC 'pass' rates are presented across indicated requirement/attribute buckets. Each column in the *'CGP Variant Class'* section reflects a separate variant class QC check routinely performed on each sample tested via PCR-CGP, with column headers reflecting the pertinent variant class (i.e., 'MSI') and number of samples passing that particular QC check across the full sample subset (i.e., '(n=8,118)'). *TC ≥ 20% is a QC metric for all variant classes and thus is equivalent to a QC fail for all variants (reflected in the 0% Full QC Pass rate); in the variant class QC pass metrics it was not applied to isolate the impact of TC on other QC metrics.

Supplementary Table 7 - PCR-CGP vs MSK-IMPACT Biomarker Frequencies

| Cancer Type | Gene | Variant Type | PCR-CGP | | | | MSK-IMPACT | | | |
|---|---|---|---|---|---|---|---|---|---|---|
| | | | Variant *n* | Sample *n* | Cancer Type Freq | Overall Freq | Variant *n* | Sample *n* | Cancer Type Freq | Overall Freq |
| Biliary_Liver | AKT1 | MUT | 0 | 558 | 0.00000 | 0.01026 | 1 | 343 | 0.00292 | 0.01045 |
| Biliary_Liver | ALK | AMP | 0 | 558 | 0.00000 | 0.00018 | 0 | 343 | 0.00000 | 0.00039 |
| Biliary_Liver | ALK | FUSION | 0 | 558 | 0.00000 | 0.00570 | 0 | 343 | 0.00000 | 0.00474 |
| Biliary_Liver | ALK | MUT | 0 | 558 | 0.00000 | 0.00061 | 0 | 343 | 0.00000 | 0.00184 |
| Biliary_Liver | AR | AMP | 0 | 558 | 0.00000 | 0.00601 | 0 | 343 | 0.00000 | 0.00842 |
| Biliary_Liver | AR | MUT | 0 | 558 | 0.00000 | 0.00136 | 0 | 343 | 0.00000 | 0.00213 |
| Biliary_Liver | ARAF | MUT | 1 | 558 | 0.00179 | 0.00039 | 1 | 343 | 0.00292 | 0.00068 |
| Biliary_Liver | BRAF | AMP | 1 | 558 | 0.00179 | 0.00250 | 0 | 343 | 0.00000 | 0.00164 |
| Biliary_Liver | BRAF | FUSION | 0 | 558 | 0.00000 | 0.00118 | 1 | 343 | 0.00292 | 0.00368 |
| Biliary_Liver | BRAF | MUT | 12 | 558 | 0.02151 | 0.04106 | 7 | 343 | 0.02041 | 0.04247 |
| Biliary_Liver | CCND1 | AMP | 15 | 558 | 0.02688 | 0.04299 | 11 | 343 | 0.03207 | 0.04189 |
| Biliary_Liver | CDK4 | AMP | 6 | 558 | 0.01075 | 0.01921 | 3 | 343 | 0.00875 | 0.02632 |
| Biliary_Liver | CDK4 | MUT | 1 | 558 | 0.00179 | 0.00061 | 0 | 343 | 0.00000 | 0.00116 |
| Biliary_Liver | CDK6 | AMP | 4 | 558 | 0.00717 | 0.00697 | 3 | 343 | 0.00875 | 0.00590 |
| Biliary_Liver | CDKN2A | HOMDEL | 95 | 558 | 0.17025 | 0.12234 | 23 | 343 | 0.06706 | 0.07498 |
| Biliary_Liver | CDKN2A | MUT | 31 | 558 | 0.05556 | 0.04917 | 6 | 343 | 0.01749 | 0.03599 |
| Biliary_Liver | CTNNB1 | MUT | 39 | 558 | 0.06989 | 0.02601 | 36 | 343 | 0.10496 | 0.02254 |
| Biliary_Liver | EGFR | AMP | 4 | 558 | 0.00717 | 0.02996 | 4 | 343 | 0.01166 | 0.03154 |
| Biliary_Liver | EGFR | MUT | 0 | 558 | 0.00000 | 0.02649 | 1 | 343 | 0.00292 | 0.04112 |
| Biliary_Liver | ERBB2 | AMP | 26 | 558 | 0.04659 | 0.03202 | 10 | 343 | 0.02915 | 0.03938 |
| Biliary_Liver | ERBB2 | MUT | 14 | 558 | 0.02509 | 0.01689 | 3 | 343 | 0.00875 | 0.01945 |
| Biliary_Liver | ERBB3 | MUT | 1 | 558 | 0.00179 | 0.00561 | 5 | 343 | 0.01458 | 0.00987 |
| Biliary_Liver | ERBB4 | MUT | 0 | 558 | 0.00000 | 0.00022 | 0 | 343 | 0.00000 | 0.00155 |
| Biliary_Liver | ESR1 | MUT | 0 | 558 | 0.00000 | 0.00965 | 0 | 343 | 0.00000 | 0.01006 |
| Biliary_Liver | EZH2 | MUT | 0 | 558 | 0.00000 | 0.00140 | 1 | 343 | 0.00292 | 0.00281 |
| Biliary_Liver | FGFR1 | AMP | 4 | 558 | 0.00717 | 0.02816 | 1 | 343 | 0.00292 | 0.02554 |
| Biliary_Liver | FGFR1 | FUSION | 0 | 558 | 0.00000 | 0.00013 | 0 | 343 | 0.00000 | 0.00039 |
| Biliary_Liver | FGFR2 | AMP | 1 | 558 | 0.00179 | 0.00347 | 1 | 343 | 0.00292 | 0.00358 |
| Biliary_Liver | FGFR2 | FUSION | 10 | 558 | 0.01792 | 0.00083 | 22 | 343 | 0.06414 | 0.00300 |
| Biliary_Liver | FGFR2 | MUT | 6 | 558 | 0.01075 | 0.00531 | 2 | 343 | 0.00583 | 0.00435 |
| Biliary_Liver | FGFR3 | AMP | 6 | 558 | 0.01075 | 0.00263 | 7 | 343 | 0.02041 | 0.00387 |
| Biliary_Liver | FGFR3 | FUSION | 1 | 558 | 0.00179 | 0.00294 | 1 | 343 | 0.00292 | 0.00271 |
| Biliary_Liver | FGFR3 | MUT | 1 | 558 | 0.00179 | 0.00588 | 0 | 343 | 0.00000 | 0.01200 |
| Biliary_Liver | FGFR4 | AMP | 2 | 558 | 0.00358 | 0.00075 | 1 | 343 | 0.00292 | 0.00252 |
| Biliary_Liver | GNA11 | MUT | 0 | 558 | 0.00000 | 0.00105 | 0 | 343 | 0.00000 | 0.00184 |
| Biliary_Liver | GNAQ | MUT | 0 | 558 | 0.00000 | 0.00096 | 0 | 343 | 0.00000 | 0.00281 |
| Biliary_Liver | HRAS | MUT | 0 | 558 | 0.00000 | 0.00504 | 0 | 343 | 0.00000 | 0.00658 |
| Biliary_Liver | IDH1 | MUT | 56 | 558 | 0.10036 | 0.01776 | 46 | 343 | 0.13411 | 0.02351 |
| Biliary_Liver | IDH2 | MUT | 18 | 558 | 0.03226 | 0.00281 | 5 | 343 | 0.01458 | 0.00290 |
| Biliary_Liver | JAK1 | MUT | 2 | 558 | 0.00358 | 0.00026 | 8 | 343 | 0.02332 | 0.00755 |
| Biliary_Liver | JAK2 | MUT | 1 | 558 | 0.00179 | 0.00066 | 0 | 343 | 0.00000 | 0.00019 |
| Biliary_Liver | KIT | AMP | 1 | 558 | 0.00179 | 0.00675 | 0 | 343 | 0.00000 | 0.00832 |
| Biliary_Liver | KIT | MUT | 0 | 558 | 0.00000 | 0.00675 | 1 | 343 | 0.00292 | 0.01335 |
| Biliary_Liver | KRAS | AMP | 14 | 558 | 0.02509 | 0.01614 | 2 | 343 | 0.00583 | 0.01809 |
| Biliary_Liver | KRAS | MUT | 71 | 558 | 0.12724 | 0.17580 | 29 | 343 | 0.08455 | 0.15257 |
| Biliary_Liver | MAP2K1 | MUT | 3 | 558 | 0.00538 | 0.00412 | 3 | 343 | 0.00875 | 0.00629 |
| Biliary_Liver | MAP2K2 | MUT | 0 | 558 | 0.00000 | 0.00022 | 0 | 343 | 0.00000 | 0.00058 |
| Biliary_Liver | MAP2K4 | MUT | 0 | 558 | 0.00000 | 0.00092 | 1 | 343 | 0.00292 | 0.00687 |
| Biliary_Liver | MAPK1 | MUT | 0 | 558 | 0.00000 | 0.00079 | 0 | 343 | 0.00000 | 0.00116 |
| Biliary_Liver | MDM2 | AMP | 22 | 558 | 0.03943 | 0.02772 | 15 | 343 | 0.04373 | 0.03696 |
| Biliary_Liver | MET | AMP | 8 | 558 | 0.01434 | 0.00693 | 2 | 343 | 0.00583 | 0.00948 |
| Biliary_Liver | MET | MUT | 1 | 558 | 0.00179 | 0.00355 | 0 | 343 | 0.00000 | 0.00300 |
| Biliary_Liver | MTOR | MUT | 0 | 558 | 0.00000 | 0.00390 | 0 | 343 | 0.00000 | 0.00619 |
| Biliary_Liver | MYC | AMP | 19 | 558 | 0.03405 | 0.04066 | 9 | 343 | 0.02624 | 0.03957 |
| Biliary_Liver | MYCN | AMP | 1 | 558 | 0.00179 | 0.00325 | 1 | 343 | 0.00292 | 0.00484 |
| Biliary_Liver | MYD88 | MUT | 1 | 558 | 0.00179 | 0.00118 | 0 | 343 | 0.00000 | 0.00145 |
| Biliary_Liver | NOTCH1 | FUSION | 0 | 558 | 0.00000 | 0.00018 | 0 | 343 | 0.00000 | 0.00048 |
| Biliary_Liver | NRAS | MUT | 16 | 558 | 0.02867 | 0.02048 | 7 | 343 | 0.02041 | 0.02070 |
| Biliary_Liver | NRG1 | FUSION | 0 | 558 | 0.00000 | 0.00039 | 0 | 343 | 0.00000 | 0.00039 |
| Biliary_Liver | NTRK1 | FUSION | 0 | 558 | 0.00000 | 0.00026 | 1 | 343 | 0.00292 | 0.00097 |
| Biliary_Liver | NTRK3 | FUSION | 0 | 558 | 0.00000 | 0.00066 | 0 | 343 | 0.00000 | 0.00097 |
| Biliary_Liver | PDGFRA | AMP | 1 | 558 | 0.00179 | 0.00627 | 0 | 343 | 0.00000 | 0.00851 |
| Biliary_Liver | PDGFRA | MUT | 0 | 558 | 0.00000 | 0.00048 | 0 | 343 | 0.00000 | 0.00223 |
| Biliary_Liver | PIK3CA | AMP | 1 | 558 | 0.00179 | 0.02298 | 3 | 343 | 0.00875 | 0.00697 |
| Biliary_Liver | PIK3CA | MUT | 30 | 558 | 0.05376 | 0.12387 | 13 | 343 | 0.03790 | 0.11474 |
| Biliary_Liver | POLE | MUT | 0 | 558 | 0.00000 | 0.00079 | 0 | 343 | 0.00000 | 0.00252 |
| Biliary_Liver | PRKACA | FUSION | 7 | 558 | 0.01254 | 0.00039 | 9 | 343 | 0.02624 | 0.00087 |
| Biliary_Liver | PTEN | HOMDEL | 5 | 558 | 0.00896 | 0.03228 | 5 | 343 | 0.01458 | 0.02390 |
| Biliary_Liver | PTEN | MUT | 12 | 558 | 0.02151 | 0.06536 | 5 | 343 | 0.01458 | 0.05563 |

| | | | | | | | | | | |
|---|---|---|---|---|---|---|---|---|---|---|
| Biliary_Liver | RAF1 | MUT | 4 | 558 | 0.00717 | 0.00154 | 2 | 343 | 0.00583 | 0.00261 |
| Biliary_Liver | RB1 | HOMDEL | 6 | 558 | 0.01075 | 0.02298 | 4 | 343 | 0.01166 | 0.01896 |
| Biliary_Liver | RB1 | MUT | 8 | 558 | 0.01434 | 0.04053 | 6 | 343 | 0.01749 | 0.04092 |
| Biliary_Liver | RET | FUSION | 0 | 558 | 0.00000 | 0.00219 | 0 | 343 | 0.00000 | 0.00281 |
| Biliary_Liver | RET | MUT | 0 | 558 | 0.00000 | 0.00175 | 0 | 343 | 0.00000 | 0.00213 |
| Biliary_Liver | RIT1 | MUT | 0 | 558 | 0.00000 | 0.00105 | 2 | 343 | 0.00583 | 0.00126 |
| Biliary_Liver | ROS1 | FUSION | 0 | 558 | 0.00000 | 0.00149 | 0 | 343 | 0.00000 | 0.00358 |
| Biliary_Liver | ROS1 | MUT | 0 | 558 | 0.00000 | 0.00009 | 0 | 343 | 0.00000 | 0.00019 |
| Biliary_Liver | SF3B1 | MUT | 13 | 558 | 0.02330 | 0.00807 | 4 | 343 | 0.01166 | 0.00803 |
| Biliary_Liver | SMO | MUT | 0 | 558 | 0.00000 | 0.00013 | 0 | 343 | 0.00000 | 0.00039 |
| Biliary_Liver | SPOP | MUT | 0 | 558 | 0.00000 | 0.00355 | 0 | 343 | 0.00000 | 0.00793 |
| Biliary_Liver | TERT | FUSION | 0 | 558 | 0.00000 | 0.00009 | 0 | 343 | 0.00000 | 0.00000 |
| Biliary_Liver | TERT | MUT | 78 | 558 | 0.13978 | 0.09795 | 44 | 343 | 0.12828 | 0.12307 |
| Biliary_Liver | TP53 | HOMDEL | 4 | 558 | 0.00717 | 0.01276 | 2 | 343 | 0.00583 | 0.00900 |
| Biliary_Liver | TP53 | MUT | 198 | 558 | 0.35484 | 0.52404 | 90 | 343 | 0.26239 | 0.41428 |
| Bladder | AKT1 | MUT | 5 | 614 | 0.00814 | 0.01026 | 3 | 402 | 0.00746 | 0.01045 |
| Bladder | ALK | AMP | 1 | 614 | 0.00163 | 0.00018 | 0 | 402 | 0.00000 | 0.00039 |
| Bladder | ALK | FUSION | 0 | 614 | 0.00000 | 0.00570 | 0 | 402 | 0.00000 | 0.00474 |
| Bladder | ALK | MUT | 0 | 614 | 0.00000 | 0.00061 | 0 | 402 | 0.00000 | 0.00184 |
| Bladder | AR | AMP | 0 | 614 | 0.00000 | 0.00601 | 0 | 402 | 0.00000 | 0.00842 |
| Bladder | AR | MUT | 1 | 614 | 0.00163 | 0.00136 | 0 | 402 | 0.00000 | 0.00213 |
| Bladder | ARAF | MUT | 0 | 614 | 0.00000 | 0.00039 | 0 | 402 | 0.00000 | 0.00068 |
| Bladder | BRAF | AMP | 1 | 614 | 0.00163 | 0.00250 | 1 | 402 | 0.00249 | 0.00164 |
| Bladder | BRAF | FUSION | 1 | 614 | 0.00163 | 0.00118 | 0 | 402 | 0.00000 | 0.00368 |
| Bladder | BRAF | MUT | 6 | 614 | 0.00977 | 0.04106 | 10 | 402 | 0.02488 | 0.04247 |
| Bladder | CCND1 | AMP | 55 | 614 | 0.08958 | 0.04299 | 38 | 402 | 0.09453 | 0.04189 |
| Bladder | CDK4 | AMP | 11 | 614 | 0.01792 | 0.01921 | 6 | 402 | 0.01493 | 0.02632 |
| Bladder | CDK4 | MUT | 1 | 614 | 0.00163 | 0.00061 | 1 | 402 | 0.00249 | 0.00116 |
| Bladder | CDK6 | AMP | 2 | 614 | 0.00326 | 0.00697 | 1 | 402 | 0.00249 | 0.00590 |
| Bladder | CDKN2A | HOMDEL | 150 | 614 | 0.24430 | 0.12234 | 65 | 402 | 0.16169 | 0.07498 |
| Bladder | CDKN2A | MUT | 35 | 614 | 0.05700 | 0.04917 | 21 | 402 | 0.05224 | 0.03599 |
| Bladder | CTNNB1 | MUT | 13 | 614 | 0.02117 | 0.02601 | 10 | 402 | 0.02488 | 0.02254 |
| Bladder | EGFR | AMP | 14 | 614 | 0.02280 | 0.02996 | 7 | 402 | 0.01741 | 0.03154 |
| Bladder | EGFR | MUT | 4 | 614 | 0.00651 | 0.02649 | 7 | 402 | 0.01741 | 0.04112 |
| Bladder | ERBB2 | AMP | 45 | 614 | 0.07329 | 0.03202 | 23 | 402 | 0.05721 | 0.03938 |
| Bladder | ERBB2 | MUT | 43 | 614 | 0.07003 | 0.01689 | 39 | 402 | 0.09701 | 0.01945 |
| Bladder | ERBB3 | MUT | 18 | 614 | 0.02932 | 0.00561 | 20 | 402 | 0.04975 | 0.00987 |
| Bladder | ERBB4 | MUT | 0 | 614 | 0.00000 | 0.00022 | 1 | 402 | 0.00249 | 0.00155 |
| Bladder | ESR1 | MUT | 0 | 614 | 0.00000 | 0.00965 | 0 | 402 | 0.00000 | 0.01006 |
| Bladder | EZH2 | MUT | 0 | 614 | 0.00000 | 0.00140 | 2 | 402 | 0.00498 | 0.00281 |
| Bladder | FGFR1 | AMP | 10 | 614 | 0.01629 | 0.02816 | 7 | 402 | 0.01741 | 0.02554 |
| Bladder | FGFR1 | FUSION | 0 | 614 | 0.00000 | 0.00013 | 0 | 402 | 0.00000 | 0.00039 |
| Bladder | FGFR2 | AMP | 2 | 614 | 0.00326 | 0.00347 | 1 | 402 | 0.00249 | 0.00358 |
| Bladder | FGFR2 | FUSION | 0 | 614 | 0.00000 | 0.00083 | 0 | 402 | 0.00000 | 0.00300 |
| Bladder | FGFR2 | MUT | 2 | 614 | 0.00326 | 0.00531 | 6 | 402 | 0.01493 | 0.00435 |
| Bladder | FGFR3 | AMP | 5 | 614 | 0.00814 | 0.00263 | 4 | 402 | 0.00995 | 0.00387 |
| Bladder | FGFR3 | FUSION | 18 | 614 | 0.02932 | 0.00294 | 5 | 402 | 0.01244 | 0.00271 |
| Bladder | FGFR3 | MUT | 88 | 614 | 0.14332 | 0.00588 | 106 | 402 | 0.26368 | 0.01200 |
| Bladder | FGFR4 | AMP | 0 | 614 | 0.00000 | 0.00075 | 0 | 402 | 0.00000 | 0.00252 |
| Bladder | GNA11 | MUT | 0 | 614 | 0.00000 | 0.00105 | 0 | 402 | 0.00000 | 0.00184 |
| Bladder | GNAQ | MUT | 0 | 614 | 0.00000 | 0.00096 | 0 | 402 | 0.00000 | 0.00281 |
| Bladder | HRAS | MUT | 17 | 614 | 0.02769 | 0.00504 | 15 | 402 | 0.03731 | 0.00658 |
| Bladder | IDH1 | MUT | 0 | 614 | 0.00000 | 0.01776 | 0 | 402 | 0.00000 | 0.02351 |
| Bladder | IDH2 | MUT | 1 | 614 | 0.00163 | 0.00281 | 0 | 402 | 0.00000 | 0.00290 |
| Bladder | JAK1 | MUT | 0 | 614 | 0.00000 | 0.00026 | 3 | 402 | 0.00746 | 0.00755 |
| Bladder | JAK2 | MUT | 0 | 614 | 0.00000 | 0.00066 | 1 | 402 | 0.00249 | 0.00019 |
| Bladder | KIT | AMP | 4 | 614 | 0.00651 | 0.00675 | 0 | 402 | 0.00000 | 0.00832 |
| Bladder | KIT | MUT | 0 | 614 | 0.00000 | 0.00675 | 0 | 402 | 0.00000 | 0.01335 |
| Bladder | KRAS | AMP | 1 | 614 | 0.00163 | 0.01614 | 4 | 402 | 0.00995 | 0.01809 |
| Bladder | KRAS | MUT | 26 | 614 | 0.04235 | 0.17580 | 26 | 402 | 0.06468 | 0.15257 |
| Bladder | MAP2K1 | MUT | 1 | 614 | 0.00163 | 0.00412 | 2 | 402 | 0.00498 | 0.00629 |
| Bladder | MAP2K2 | MUT | 0 | 614 | 0.00000 | 0.00022 | 0 | 402 | 0.00000 | 0.00058 |
| Bladder | MAP2K4 | MUT | 0 | 614 | 0.00000 | 0.00092 | 1 | 402 | 0.00249 | 0.00687 |
| Bladder | MAPK1 | MUT | 1 | 614 | 0.00163 | 0.00079 | 4 | 402 | 0.00995 | 0.00116 |
| Bladder | MDM2 | AMP | 46 | 614 | 0.07492 | 0.02772 | 32 | 402 | 0.07960 | 0.03696 |
| Bladder | MET | AMP | 0 | 614 | 0.00000 | 0.00693 | 2 | 402 | 0.00498 | 0.00948 |
| Bladder | MET | MUT | 0 | 614 | 0.00000 | 0.00355 | 1 | 402 | 0.00249 | 0.00300 |
| Bladder | MTOR | MUT | 3 | 614 | 0.00489 | 0.00390 | 4 | 402 | 0.00995 | 0.00619 |
| Bladder | MYC | AMP | 19 | 614 | 0.03094 | 0.04066 | 7 | 402 | 0.01741 | 0.03957 |
| Bladder | MYCN | AMP | 6 | 614 | 0.00977 | 0.00325 | 4 | 402 | 0.00995 | 0.00484 |
| Bladder | MYD88 | MUT | 0 | 614 | 0.00000 | 0.00118 | 0 | 402 | 0.00000 | 0.00145 |
| Bladder | NOTCH1 | FUSION | 0 | 614 | 0.00000 | 0.00018 | 0 | 402 | 0.00000 | 0.00048 |
| Bladder | NRAS | MUT | 4 | 614 | 0.00651 | 0.02048 | 5 | 402 | 0.01244 | 0.02070 |
| Bladder | NRG1 | FUSION | 0 | 614 | 0.00000 | 0.00039 | 0 | 402 | 0.00000 | 0.00039 |
| Bladder | NTRK1 | FUSION | 0 | 614 | 0.00000 | 0.00026 | 0 | 402 | 0.00000 | 0.00097 |
| Bladder | NTRK3 | FUSION | 0 | 614 | 0.00000 | 0.00066 | 0 | 402 | 0.00000 | 0.00097 |

| | | | | | | | | | | |
|---|---|---|---|---|---|---|---|---|---|---|
| Bladder | PDGFRA | AMP | 2 | 614 | 0.00326 | 0.00627 | 0 | 402 | 0.00000 | 0.00851 |
| Bladder | PDGFRA | MUT | 0 | 614 | 0.00000 | 0.00048 | 0 | 402 | 0.00000 | 0.00223 |
| Bladder | PIK3CA | AMP | 10 | 614 | 0.01629 | 0.02298 | 1 | 402 | 0.00249 | 0.00697 |
| Bladder | PIK3CA | MUT | 113 | 614 | 0.18404 | 0.12387 | 84 | 402 | 0.20896 | 0.11474 |
| Bladder | POLE | MUT | 0 | 614 | 0.00000 | 0.00079 | 3 | 402 | 0.00746 | 0.00252 |
| Bladder | PRKACA | FUSION | 0 | 614 | 0.00000 | 0.00039 | 0 | 402 | 0.00000 | 0.00087 |
| Bladder | PTEN | HOMDEL | 7 | 614 | 0.01140 | 0.03228 | 1 | 402 | 0.00249 | 0.02390 |
| Bladder | PTEN | MUT | 20 | 614 | 0.03257 | 0.06536 | 8 | 402 | 0.01990 | 0.05563 |
| Bladder | RAF1 | MUT | 0 | 614 | 0.00000 | 0.00154 | 3 | 402 | 0.00746 | 0.00261 |
| Bladder | RB1 | HOMDEL | 20 | 614 | 0.03257 | 0.02298 | 7 | 402 | 0.01741 | 0.01896 |
| Bladder | RB1 | MUT | 77 | 614 | 0.12541 | 0.04053 | 48 | 402 | 0.11940 | 0.04092 |
| Bladder | RET | FUSION | 0 | 614 | 0.00000 | 0.00219 | 0 | 402 | 0.00000 | 0.00281 |
| Bladder | RET | MUT | 0 | 614 | 0.00000 | 0.00175 | 0 | 402 | 0.00000 | 0.00213 |
| Bladder | RIT1 | MUT | 3 | 614 | 0.00489 | 0.00105 | 3 | 402 | 0.00746 | 0.00126 |
| Bladder | ROS1 | FUSION | 0 | 614 | 0.00000 | 0.00149 | 1 | 402 | 0.00249 | 0.00358 |
| Bladder | ROS1 | MUT | 0 | 614 | 0.00000 | 0.00009 | 0 | 402 | 0.00000 | 0.00019 |
| Bladder | SF3B1 | MUT | 3 | 614 | 0.00489 | 0.00807 | 9 | 402 | 0.02239 | 0.00803 |
| Bladder | SMO | MUT | 0 | 614 | 0.00000 | 0.00013 | 1 | 402 | 0.00249 | 0.00039 |
| Bladder | SPOP | MUT | 1 | 614 | 0.00163 | 0.00355 | 1 | 402 | 0.00249 | 0.00793 |
| Bladder | TERT | FUSION | 0 | 614 | 0.00000 | 0.00009 | 0 | 402 | 0.00000 | 0.00000 |
| Bladder | TERT | MUT | 375 | 614 | 0.61075 | 0.09795 | 263 | 402 | 0.65423 | 0.12307 |
| Bladder | TP53 | HOMDEL | 9 | 614 | 0.01466 | 0.01276 | 1 | 402 | 0.00249 | 0.00900 |
| Bladder | TP53 | MUT | 354 | 614 | 0.57655 | 0.52404 | 160 | 402 | 0.39801 | 0.41428 |
| Brain | AKT1 | MUT | 2 | 1203 | 0.00166 | 0.01026 | 1 | 513 | 0.00195 | 0.01045 |
| Brain | ALK | AMP | 0 | 1203 | 0.00000 | 0.00018 | 0 | 513 | 0.00000 | 0.00039 |
| Brain | ALK | FUSION | 1 | 1203 | 0.00083 | 0.00570 | 0 | 513 | 0.00000 | 0.00474 |
| Brain | ALK | MUT | 0 | 1203 | 0.00000 | 0.00061 | 0 | 513 | 0.00000 | 0.00184 |
| Brain | AR | AMP | 0 | 1203 | 0.00000 | 0.00601 | 0 | 513 | 0.00000 | 0.00842 |
| Brain | AR | MUT | 0 | 1203 | 0.00000 | 0.00136 | 0 | 513 | 0.00000 | 0.00213 |
| Brain | ARAF | MUT | 0 | 1203 | 0.00000 | 0.00039 | 0 | 513 | 0.00000 | 0.00068 |
| Brain | BRAF | AMP | 1 | 1203 | 0.00083 | 0.00250 | 2 | 513 | 0.00390 | 0.00164 |
| Brain | BRAF | FUSION | 4 | 1203 | 0.00333 | 0.00118 | 4 | 513 | 0.00780 | 0.00368 |
| Brain | BRAF | MUT | 39 | 1203 | 0.03242 | 0.04106 | 10 | 513 | 0.01949 | 0.04247 |
| Brain | CCND1 | AMP | 4 | 1203 | 0.00333 | 0.04299 | 2 | 513 | 0.00390 | 0.04189 |
| Brain | CDK4 | AMP | 123 | 1203 | 0.10224 | 0.01921 | 57 | 513 | 0.11111 | 0.02632 |
| Brain | CDK4 | MUT | 0 | 1203 | 0.00000 | 0.00061 | 0 | 513 | 0.00000 | 0.00116 |
| Brain | CDK6 | AMP | 11 | 1203 | 0.00914 | 0.00697 | 8 | 513 | 0.01559 | 0.00590 |
| Brain | CDKN2A | HOMDEL | 425 | 1203 | 0.35328 | 0.12234 | 169 | 513 | 0.32943 | 0.07498 |
| Brain | CDKN2A | MUT | 23 | 1203 | 0.01912 | 0.04917 | 9 | 513 | 0.01754 | 0.03599 |
| Brain | CTNNB1 | MUT | 1 | 1203 | 0.00083 | 0.02601 | 0 | 513 | 0.00000 | 0.02254 |
| Brain | EGFR | AMP | 323 | 1203 | 0.26850 | 0.02996 | 121 | 513 | 0.23587 | 0.03154 |
| Brain | EGFR | FUSION | 23 | 1203 | 0.01912 | 0.00110 | 2 | 513 | 0.00390 | 0.00019 |
| Brain | EGFR | MUT | 109 | 1203 | 0.09061 | 0.02649 | 57 | 513 | 0.11111 | 0.04112 |
| Brain | ERBB2 | AMP | 0 | 1203 | 0.00000 | 0.03202 | 0 | 513 | 0.00000 | 0.03938 |
| Brain | ERBB2 | MUT | 0 | 1203 | 0.00000 | 0.01689 | 0 | 513 | 0.00000 | 0.01945 |
| Brain | ERBB3 | MUT | 1 | 1203 | 0.00083 | 0.00561 | 0 | 513 | 0.00000 | 0.00987 |
| Brain | ERBB4 | MUT | 0 | 1203 | 0.00000 | 0.00022 | 0 | 513 | 0.00000 | 0.00155 |
| Brain | ESR1 | MUT | 0 | 1203 | 0.00000 | 0.00965 | 0 | 513 | 0.00000 | 0.01006 |
| Brain | EZH2 | MUT | 0 | 1203 | 0.00000 | 0.00140 | 1 | 513 | 0.00195 | 0.00281 |
| Brain | FGFR1 | AMP | 0 | 1203 | 0.00000 | 0.02816 | 0 | 513 | 0.00000 | 0.02554 |
| Brain | FGFR1 | FUSION | 0 | 1203 | 0.00000 | 0.00013 | 0 | 513 | 0.00000 | 0.00039 |
| Brain | FGFR2 | AMP | 0 | 1203 | 0.00000 | 0.00347 | 0 | 513 | 0.00000 | 0.00358 |
| Brain | FGFR2 | FUSION | 0 | 1203 | 0.00000 | 0.00083 | 0 | 513 | 0.00000 | 0.00300 |
| Brain | FGFR2 | MUT | 0 | 1203 | 0.00000 | 0.00531 | 0 | 513 | 0.00000 | 0.00435 |
| Brain | FGFR3 | AMP | 5 | 1203 | 0.00416 | 0.00263 | 3 | 513 | 0.00585 | 0.00387 |
| Brain | FGFR3 | FUSION | 20 | 1203 | 0.01663 | 0.00294 | 12 | 513 | 0.02339 | 0.00271 |
| Brain | FGFR3 | MUT | 3 | 1203 | 0.00249 | 0.00588 | 1 | 513 | 0.00195 | 0.01200 |
| Brain | FGFR4 | AMP | 0 | 1203 | 0.00000 | 0.00075 | 1 | 513 | 0.00195 | 0.00252 |
| Brain | GNA11 | MUT | 0 | 1203 | 0.00000 | 0.00105 | 0 | 513 | 0.00000 | 0.00184 |
| Brain | GNAQ | MUT | 0 | 1203 | 0.00000 | 0.00096 | 0 | 513 | 0.00000 | 0.00281 |
| Brain | HRAS | MUT | 1 | 1203 | 0.00083 | 0.00504 | 1 | 513 | 0.00195 | 0.00658 |
| Brain | IDH1 | MUT | 256 | 1203 | 0.21280 | 0.01776 | 165 | 513 | 0.32164 | 0.02351 |
| Brain | IDH2 | MUT | 15 | 1203 | 0.01247 | 0.00281 | 13 | 513 | 0.02534 | 0.00290 |
| Brain | JAK1 | MUT | 0 | 1203 | 0.00000 | 0.00026 | 1 | 513 | 0.00195 | 0.00755 |
| Brain | JAK2 | MUT | 0 | 1203 | 0.00000 | 0.00066 | 0 | 513 | 0.00000 | 0.00019 |
| Brain | KIT | AMP | 48 | 1203 | 0.03990 | 0.00675 | 33 | 513 | 0.06433 | 0.00832 |
| Brain | KIT | MUT | 1 | 1203 | 0.00083 | 0.00675 | 0 | 513 | 0.00000 | 0.01335 |
| Brain | KRAS | AMP | 10 | 1203 | 0.00831 | 0.01614 | 8 | 513 | 0.01559 | 0.01809 |
| Brain | KRAS | MUT | 12 | 1203 | 0.00998 | 0.17580 | 4 | 513 | 0.00780 | 0.15257 |
| Brain | MAP2K1 | MUT | 1 | 1203 | 0.00083 | 0.00412 | 2 | 513 | 0.00390 | 0.00629 |
| Brain | MAP2K2 | MUT | 0 | 1203 | 0.00000 | 0.00022 | 0 | 513 | 0.00000 | 0.00058 |
| Brain | MAP2K4 | MUT | 0 | 1203 | 0.00000 | 0.00092 | 0 | 513 | 0.00000 | 0.00687 |
| Brain | MAPK1 | MUT | 0 | 1203 | 0.00000 | 0.00079 | 0 | 513 | 0.00000 | 0.00116 |
| Brain | MDM2 | AMP | 70 | 1203 | 0.05819 | 0.02772 | 33 | 513 | 0.06433 | 0.03696 |
| Brain | MET | AMP | 15 | 1203 | 0.01247 | 0.00693 | 9 | 513 | 0.01754 | 0.00948 |
| Brain | MET | MUT | 1 | 1203 | 0.00083 | 0.00355 | 0 | 513 | 0.00000 | 0.00300 |

| | | | | | | | | | | |
|---|---|---|---|---|---|---|---|---|---|---|
| Brain | MTOR | MUT | 5 | 1203 | 0.00416 | 0.00390 | 2 | 513 | 0.00390 | 0.00619 |
| Brain | MYC | AMP | 3 | 1203 | 0.00249 | 0.04066 | 5 | 513 | 0.00975 | 0.03957 |
| Brain | MYCN | AMP | 12 | 1203 | 0.00998 | 0.00325 | 10 | 513 | 0.01949 | 0.00484 |
| Brain | MYD88 | MUT | 0 | 1203 | 0.00000 | 0.00118 | 0 | 513 | 0.00000 | 0.00145 |
| Brain | NOTCH1 | FUSION | 0 | 1203 | 0.00000 | 0.00018 | 1 | 513 | 0.00195 | 0.00048 |
| Brain | NRAS | MUT | 5 | 1203 | 0.00416 | 0.02048 | 2 | 513 | 0.00390 | 0.02070 |
| Brain | NRG1 | FUSION | 0 | 1203 | 0.00000 | 0.00039 | 0 | 513 | 0.00000 | 0.00039 |
| Brain | NTRK1 | FUSION | 1 | 1203 | 0.00083 | 0.00026 | 0 | 513 | 0.00000 | 0.00097 |
| Brain | NTRK3 | FUSION | 0 | 1203 | 0.00000 | 0.00066 | 1 | 513 | 0.00195 | 0.00097 |
| Brain | PDGFRA | AMP | 64 | 1203 | 0.05320 | 0.00627 | 40 | 513 | 0.07797 | 0.00851 |
| Brain | PDGFRA | MUT | 8 | 1203 | 0.00665 | 0.00048 | 7 | 513 | 0.01365 | 0.00223 |
| Brain | PIK3CA | AMP | 5 | 1203 | 0.00416 | 0.02298 | 1 | 513 | 0.00195 | 0.00697 |
| Brain | PIK3CA | MUT | 113 | 1203 | 0.09393 | 0.12387 | 57 | 513 | 0.11111 | 0.11474 |
| Brain | POLE | MUT | 0 | 1203 | 0.00000 | 0.00079 | 0 | 513 | 0.00000 | 0.00252 |
| Brain | PRKACA | FUSION | 0 | 1203 | 0.00000 | 0.00039 | 0 | 513 | 0.00000 | 0.00087 |
| Brain | PTEN | HOMDEL | 49 | 1203 | 0.04073 | 0.03228 | 44 | 513 | 0.08577 | 0.02390 |
| Brain | PTEN | MUT | 273 | 1203 | 0.22693 | 0.06536 | 116 | 513 | 0.22612 | 0.05563 |
| Brain | RAF1 | MUT | 2 | 1203 | 0.00166 | 0.00154 | 0 | 513 | 0.00000 | 0.00261 |
| Brain | RB1 | HOMDEL | 11 | 1203 | 0.00914 | 0.02298 | 10 | 513 | 0.01949 | 0.01896 |
| Brain | RB1 | MUT | 53 | 1203 | 0.04406 | 0.04053 | 40 | 513 | 0.07797 | 0.04092 |
| Brain | RET | FUSION | 1 | 1203 | 0.00083 | 0.00219 | 0 | 513 | 0.00000 | 0.00281 |
| Brain | RET | MUT | 1 | 1203 | 0.00083 | 0.00175 | 0 | 513 | 0.00000 | 0.00213 |
| Brain | RIT1 | MUT | 1 | 1203 | 0.00083 | 0.00105 | 0 | 513 | 0.00000 | 0.00126 |
| Brain | ROS1 | FUSION | 1 | 1203 | 0.00083 | 0.00149 | 1 | 513 | 0.00195 | 0.00358 |
| Brain | ROS1 | MUT | 0 | 1203 | 0.00000 | 0.00009 | 0 | 513 | 0.00000 | 0.00019 |
| Brain | SF3B1 | MUT | 1 | 1203 | 0.00083 | 0.00807 | 0 | 513 | 0.00000 | 0.00803 |
| Brain | SMO | MUT | 2 | 1203 | 0.00166 | 0.00013 | 0 | 513 | 0.00000 | 0.00039 |
| Brain | SPOP | MUT | 0 | 1203 | 0.00000 | 0.00355 | 0 | 513 | 0.00000 | 0.00793 |
| Brain | TERT | FUSION | 0 | 1203 | 0.00000 | 0.00009 | 0 | 513 | 0.00000 | 0.00000 |
| Brain | TERT | MUT | 675 | 1203 | 0.56110 | 0.09795 | 337 | 513 | 0.65692 | 0.12307 |
| Brain | TP53 | HOMDEL | 3 | 1203 | 0.00249 | 0.01276 | 6 | 513 | 0.01170 | 0.00900 |
| Brain | TP53 | MUT | 428 | 1203 | 0.35578 | 0.52404 | 199 | 513 | 0.38791 | 0.41428 |
| Breast | AKT1 | MUT | 96 | 2659 | 0.03610 | 0.01026 | 54 | 1228 | 0.04397 | 0.01045 |
| Breast | ALK | AMP | 0 | 2659 | 0.00000 | 0.00018 | 0 | 1228 | 0.00000 | 0.00039 |
| Breast | ALK | FUSION | 0 | 2659 | 0.00000 | 0.00570 | 1 | 1228 | 0.00081 | 0.00474 |
| Breast | ALK | MUT | 0 | 2659 | 0.00000 | 0.00061 | 0 | 1228 | 0.00000 | 0.00184 |
| Breast | AR | AMP | 10 | 2659 | 0.00376 | 0.00601 | 4 | 1228 | 0.00326 | 0.00842 |
| Breast | AR | MUT | 1 | 2659 | 0.00038 | 0.00136 | 0 | 1228 | 0.00000 | 0.00213 |
| Breast | ARAF | MUT | 1 | 2659 | 0.00038 | 0.00039 | 0 | 1228 | 0.00000 | 0.00068 |
| Breast | BRAF | AMP | 7 | 2659 | 0.00263 | 0.00250 | 3 | 1228 | 0.00244 | 0.00164 |
| Breast | BRAF | FUSION | 0 | 2659 | 0.00000 | 0.00118 | 1 | 1228 | 0.00081 | 0.00368 |
| Breast | BRAF | MUT | 15 | 2659 | 0.00564 | 0.04106 | 3 | 1228 | 0.00244 | 0.04247 |
| Breast | CCND1 | AMP | 405 | 2659 | 0.15231 | 0.04299 | 228 | 1228 | 0.18567 | 0.04189 |
| Breast | CDK4 | AMP | 41 | 2659 | 0.01542 | 0.01921 | 19 | 1228 | 0.01547 | 0.02632 |
| Breast | CDK4 | MUT | 0 | 2659 | 0.00000 | 0.00061 | 0 | 1228 | 0.00000 | 0.00116 |
| Breast | CDK6 | AMP | 4 | 2659 | 0.00150 | 0.00697 | 4 | 1228 | 0.00326 | 0.00590 |
| Breast | CDKN2A | HOMDEL | 131 | 2659 | 0.04927 | 0.12234 | 27 | 1228 | 0.02199 | 0.07498 |
| Breast | CDKN2A | MUT | 24 | 2659 | 0.00903 | 0.04917 | 12 | 1228 | 0.00977 | 0.03599 |
| Breast | CTNNB1 | MUT | 4 | 2659 | 0.00150 | 0.02601 | 2 | 1228 | 0.00163 | 0.02254 |
| Breast | EGFR | AMP | 28 | 2659 | 0.01053 | 0.02996 | 15 | 1228 | 0.01221 | 0.03154 |
| Breast | EGFR | MUT | 2 | 2659 | 0.00075 | 0.02649 | 6 | 1228 | 0.00489 | 0.04112 |
| Breast | ERBB2 | AMP | 263 | 2659 | 0.09891 | 0.03202 | 179 | 1228 | 0.14577 | 0.03938 |
| Breast | ERBB2 | MUT | 79 | 2659 | 0.02971 | 0.01689 | 42 | 1228 | 0.03420 | 0.01945 |
| Breast | ERBB3 | MUT | 7 | 2659 | 0.00263 | 0.00561 | 14 | 1228 | 0.01140 | 0.00987 |
| Breast | ERBB4 | MUT | 0 | 2659 | 0.00000 | 0.00022 | 1 | 1228 | 0.00081 | 0.00155 |
| Breast | ESR1 | MUT | 195 | 2659 | 0.07334 | 0.00965 | 94 | 1228 | 0.07655 | 0.01006 |
| Breast | EZH2 | MUT | 0 | 2659 | 0.00000 | 0.00140 | 0 | 1228 | 0.00000 | 0.00281 |
| Breast | FGFR1 | AMP | 335 | 2659 | 0.12599 | 0.02816 | 152 | 1228 | 0.12378 | 0.02554 |
| Breast | FGFR1 | FUSION | 1 | 2659 | 0.00038 | 0.00013 | 0 | 1228 | 0.00000 | 0.00039 |
| Breast | FGFR2 | AMP | 26 | 2659 | 0.00978 | 0.00347 | 20 | 1228 | 0.01629 | 0.00358 |
| Breast | FGFR2 | FUSION | 0 | 2659 | 0.00000 | 0.00083 | 1 | 1228 | 0.00081 | 0.00300 |
| Breast | FGFR2 | MUT | 19 | 2659 | 0.00715 | 0.00531 | 7 | 1228 | 0.00570 | 0.00435 |
| Breast | FGFR3 | AMP | 9 | 2659 | 0.00338 | 0.00263 | 10 | 1228 | 0.00814 | 0.00387 |
| Breast | FGFR3 | FUSION | 2 | 2659 | 0.00075 | 0.00294 | 1 | 1228 | 0.00081 | 0.00271 |
| Breast | FGFR3 | MUT | 1 | 2659 | 0.00038 | 0.00588 | 2 | 1228 | 0.00163 | 0.01200 |
| Breast | FGFR4 | AMP | 6 | 2659 | 0.00226 | 0.00075 | 6 | 1228 | 0.00489 | 0.00252 |
| Breast | GNA11 | MUT | 0 | 2659 | 0.00000 | 0.00105 | 0 | 1228 | 0.00000 | 0.00184 |
| Breast | GNAQ | MUT | 0 | 2659 | 0.00000 | 0.00096 | 0 | 1228 | 0.00000 | 0.00281 |
| Breast | HRAS | MUT | 5 | 2659 | 0.00188 | 0.00504 | 2 | 1228 | 0.00163 | 0.00658 |
| Breast | IDH1 | MUT | 4 | 2659 | 0.00150 | 0.01776 | 1 | 1228 | 0.00081 | 0.02351 |
| Breast | IDH2 | MUT | 3 | 2659 | 0.00113 | 0.00281 | 0 | 1228 | 0.00000 | 0.00290 |
| Breast | JAK1 | MUT | 0 | 2659 | 0.00000 | 0.00026 | 8 | 1228 | 0.00651 | 0.00755 |
| Breast | JAK2 | MUT | 1 | 2659 | 0.00038 | 0.00066 | 0 | 1228 | 0.00000 | 0.00019 |
| Breast | KIT | AMP | 14 | 2659 | 0.00527 | 0.00675 | 5 | 1228 | 0.00407 | 0.00832 |
| Breast | KIT | MUT | 1 | 2659 | 0.00038 | 0.00675 | 2 | 1228 | 0.00163 | 0.01335 |
| Breast | KRAS | AMP | 30 | 2659 | 0.01128 | 0.01614 | 11 | 1228 | 0.00896 | 0.01809 |

| | | | | | | | | | | |
|---|---|---|---|---|---|---|---|---|---|---|
| Breast | KRAS | MUT | 42 | 2659 | 0.01580 | 0.17580 | 10 | 1228 | 0.00814 | 0.15257 |
| Breast | MAP2K1 | MUT | 0 | 2659 | 0.00000 | 0.00412 | 1 | 1228 | 0.00081 | 0.00629 |
| Breast | MAP2K2 | MUT | 1 | 2659 | 0.00038 | 0.00022 | 1 | 1228 | 0.00081 | 0.00058 |
| Breast | MAP2K4 | MUT | 13 | 2659 | 0.00489 | 0.00092 | 36 | 1228 | 0.02932 | 0.00687 |
| Breast | MAPK1 | MUT | 1 | 2659 | 0.00038 | 0.00079 | 0 | 1228 | 0.00000 | 0.00116 |
| Breast | MDM2 | AMP | 109 | 2659 | 0.04099 | 0.02772 | 50 | 1228 | 0.04072 | 0.03696 |
| Breast | MET | AMP | 1 | 2659 | 0.00038 | 0.00693 | 2 | 1228 | 0.00163 | 0.00948 |
| Breast | MET | MUT | 0 | 2659 | 0.00000 | 0.00355 | 0 | 1228 | 0.00000 | 0.00300 |
| Breast | MTOR | MUT | 6 | 2659 | 0.00226 | 0.00390 | 2 | 1228 | 0.00163 | 0.00619 |
| Breast | MYC | AMP | 214 | 2659 | 0.08048 | 0.04066 | 116 | 1228 | 0.09446 | 0.03957 |
| Breast | MYCN | AMP | 8 | 2659 | 0.00301 | 0.00325 | 2 | 1228 | 0.00163 | 0.00484 |
| Breast | MYD88 | MUT | 0 | 2659 | 0.00000 | 0.00118 | 1 | 1228 | 0.00081 | 0.00145 |
| Breast | NOTCH1 | FUSION | 3 | 2659 | 0.00113 | 0.00018 | 1 | 1228 | 0.00081 | 0.00048 |
| Breast | NRAS | MUT | 1 | 2659 | 0.00038 | 0.02048 | 0 | 1228 | 0.00000 | 0.02070 |
| Breast | NRG1 | FUSION | 0 | 2659 | 0.00000 | 0.00039 | 1 | 1228 | 0.00081 | 0.00039 |
| Breast | NTRK1 | FUSION | 1 | 2659 | 0.00038 | 0.00026 | 0 | 1228 | 0.00000 | 0.00097 |
| Breast | NTRK3 | FUSION | 2 | 2659 | 0.00075 | 0.00066 | 0 | 1228 | 0.00000 | 0.00097 |
| Breast | PDGFRA | AMP | 6 | 2659 | 0.00226 | 0.00627 | 5 | 1228 | 0.00407 | 0.00851 |
| Breast | PDGFRA | MUT | 0 | 2659 | 0.00000 | 0.00048 | 0 | 1228 | 0.00000 | 0.00223 |
| Breast | PIK3CA | AMP | 74 | 2659 | 0.02783 | 0.02298 | 16 | 1228 | 0.01303 | 0.00697 |
| Breast | PIK3CA | MUT | 909 | 2659 | 0.34186 | 0.12387 | 423 | 1228 | 0.34446 | 0.11474 |
| Breast | POLE | MUT | 0 | 2659 | 0.00000 | 0.00079 | 4 | 1228 | 0.00326 | 0.00252 |
| Breast | PRKACA | FUSION | 0 | 2659 | 0.00000 | 0.00039 | 0 | 1228 | 0.00000 | 0.00087 |
| Breast | PTEN | HOMDEL | 91 | 2659 | 0.03422 | 0.03228 | 29 | 1228 | 0.02362 | 0.02390 |
| Breast | PTEN | MUT | 188 | 2659 | 0.07070 | 0.06536 | 80 | 1228 | 0.06515 | 0.05563 |
| Breast | RAF1 | MUT | 1 | 2659 | 0.00038 | 0.00154 | 1 | 1228 | 0.00081 | 0.00261 |
| Breast | RB1 | HOMDEL | 56 | 2659 | 0.02106 | 0.02298 | 16 | 1228 | 0.01303 | 0.01896 |
| Breast | RB1 | MUT | 70 | 2659 | 0.02633 | 0.04053 | 31 | 1228 | 0.02524 | 0.04092 |
| Breast | RET | FUSION | 0 | 2659 | 0.00000 | 0.00219 | 0 | 1228 | 0.00000 | 0.00281 |
| Breast | RET | MUT | 2 | 2659 | 0.00075 | 0.00175 | 1 | 1228 | 0.00081 | 0.00213 |
| Breast | RIT1 | MUT | 0 | 2659 | 0.00000 | 0.00105 | 1 | 1228 | 0.00081 | 0.00126 |
| Breast | ROS1 | FUSION | 2 | 2659 | 0.00075 | 0.00149 | 0 | 1228 | 0.00000 | 0.00358 |
| Breast | ROS1 | MUT | 1 | 2659 | 0.00038 | 0.00009 | 0 | 1228 | 0.00000 | 0.00019 |
| Breast | SF3B1 | MUT | 57 | 2659 | 0.02144 | 0.00807 | 17 | 1228 | 0.01384 | 0.00803 |
| Breast | SMO | MUT | 0 | 2659 | 0.00000 | 0.00013 | 0 | 1228 | 0.00000 | 0.00039 |
| Breast | SPOP | MUT | 1 | 2659 | 0.00038 | 0.00355 | 2 | 1228 | 0.00163 | 0.00793 |
| Breast | TERT | FUSION | 0 | 2659 | 0.00000 | 0.00009 | 0 | 1228 | 0.00000 | 0.00000 |
| Breast | TERT | MUT | 15 | 2659 | 0.00564 | 0.09795 | 7 | 1228 | 0.00570 | 0.12307 |
| Breast | TP53 | HOMDEL | 39 | 2659 | 0.01467 | 0.01276 | 6 | 1228 | 0.00489 | 0.00900 |
| Breast | TP53 | MUT | 1185 | 2659 | 0.44566 | 0.52404 | 510 | 1228 | 0.41531 | 0.41428 |
| Colon and Rectum | AKT1 | MUT | 31 | 3020 | 0.01026 | 0.01026 | 12 | 1053 | 0.01140 | 0.01045 |
| Colon and Rectum | ALK | AMP | 0 | 3020 | 0.00000 | 0.00018 | 0 | 1053 | 0.00000 | 0.00039 |
| Colon and Rectum | ALK | FUSION | 2 | 3020 | 0.00066 | 0.00570 | 0 | 1053 | 0.00000 | 0.00474 |
| Colon and Rectum | ALK | MUT | 0 | 3020 | 0.00000 | 0.00061 | 3 | 1053 | 0.00285 | 0.00184 |
| Colon and Rectum | AR | AMP | 1 | 3020 | 0.00033 | 0.00601 | 0 | 1053 | 0.00000 | 0.00842 |
| Colon and Rectum | AR | MUT | 3 | 3020 | 0.00099 | 0.00136 | 0 | 1053 | 0.00000 | 0.00213 |
| Colon and Rectum | ARAF | MUT | 0 | 3020 | 0.00000 | 0.00039 | 1 | 1053 | 0.00095 | 0.00068 |
| Colon and Rectum | BRAF | AMP | 2 | 3020 | 0.00066 | 0.00250 | 1 | 1053 | 0.00095 | 0.00164 |
| Colon and Rectum | BRAF | FUSION | 2 | 3020 | 0.00066 | 0.00118 | 2 | 1053 | 0.00190 | 0.00368 |
| Colon and Rectum | BRAF | MUT | 258 | 3020 | 0.08543 | 0.04106 | 91 | 1053 | 0.08642 | 0.04247 |
| Colon and Rectum | CCND1 | AMP | 25 | 3020 | 0.00828 | 0.04299 | 10 | 1053 | 0.00950 | 0.04189 |
| Colon and Rectum | CDK4 | AMP | 0 | 3020 | 0.00000 | 0.01921 | 1 | 1053 | 0.00095 | 0.02632 |
| Colon and Rectum | CDK4 | MUT | 0 | 3020 | 0.00000 | 0.00061 | 0 | 1053 | 0.00000 | 0.00116 |
| Colon and Rectum | CDK6 | AMP | 9 | 3020 | 0.00298 | 0.00697 | 4 | 1053 | 0.00380 | 0.00590 |
| Colon and Rectum | CDKN2A | HOMDEL | 39 | 3020 | 0.01291 | 0.12234 | 10 | 1053 | 0.00950 | 0.07498 |
| Colon and Rectum | CDKN2A | MUT | 24 | 3020 | 0.00795 | 0.04917 | 12 | 1053 | 0.01140 | 0.03599 |
| Colon and Rectum | CTNNB1 | MUT | 45 | 3020 | 0.01490 | 0.02601 | 23 | 1053 | 0.02184 | 0.02254 |
| Colon and Rectum | EGFR | AMP | 47 | 3020 | 0.01556 | 0.02996 | 14 | 1053 | 0.01330 | 0.03154 |
| Colon and Rectum | EGFR | MUT | 1 | 3020 | 0.00033 | 0.02649 | 8 | 1053 | 0.00760 | 0.04112 |
| Colon and Rectum | ERBB2 | AMP | 79 | 3020 | 0.02616 | 0.03202 | 29 | 1053 | 0.02754 | 0.03938 |
| Colon and Rectum | ERBB2 | MUT | 80 | 3020 | 0.02649 | 0.01689 | 26 | 1053 | 0.02469 | 0.01945 |
| Colon and Rectum | ERBB3 | MUT | 46 | 3020 | 0.01523 | 0.00561 | 21 | 1053 | 0.01994 | 0.00987 |
| Colon and Rectum | ERBB4 | MUT | 0 | 3020 | 0.00000 | 0.00022 | 3 | 1053 | 0.00285 | 0.00155 |
| Colon and Rectum | ESR1 | MUT | 0 | 3020 | 0.00000 | 0.00965 | 0 | 1053 | 0.00000 | 0.01006 |
| Colon and Rectum | EZH2 | MUT | 0 | 3020 | 0.00000 | 0.00140 | 2 | 1053 | 0.00190 | 0.00281 |
| Colon and Rectum | FGFR1 | AMP | 56 | 3020 | 0.01854 | 0.02816 | 17 | 1053 | 0.01614 | 0.02554 |
| Colon and Rectum | FGFR1 | FUSION | 0 | 3020 | 0.00000 | 0.00013 | 2 | 1053 | 0.00190 | 0.00039 |
| Colon and Rectum | FGFR2 | AMP | 1 | 3020 | 0.00033 | 0.00347 | 0 | 1053 | 0.00000 | 0.00358 |
| Colon and Rectum | FGFR2 | FUSION | 0 | 3020 | 0.00000 | 0.00083 | 0 | 1053 | 0.00000 | 0.00300 |
| Colon and Rectum | FGFR2 | MUT | 1 | 3020 | 0.00033 | 0.00531 | 5 | 1053 | 0.00475 | 0.00435 |
| Colon and Rectum | FGFR3 | AMP | 2 | 3020 | 0.00066 | 0.00263 | 0 | 1053 | 0.00000 | 0.00387 |
| Colon and Rectum | FGFR3 | FUSION | 0 | 3020 | 0.00000 | 0.00294 | 0 | 1053 | 0.00000 | 0.00271 |
| Colon and Rectum | FGFR3 | MUT | 1 | 3020 | 0.00033 | 0.00588 | 0 | 1053 | 0.00000 | 0.01200 |
| Colon and Rectum | FGFR4 | AMP | 0 | 3020 | 0.00000 | 0.00075 | 1 | 1053 | 0.00095 | 0.00252 |
| Colon and Rectum | GNA11 | MUT | 0 | 3020 | 0.00000 | 0.00105 | 0 | 1053 | 0.00000 | 0.00184 |
| Colon and Rectum | GNAQ | MUT | 0 | 3020 | 0.00000 | 0.00096 | 0 | 1053 | 0.00000 | 0.00281 |

| | | | | | | | | | | |
|---|---|---|---|---|---|---|---|---|---|---|
| Colon and Rectum | HRAS | MUT | 1 | 3020 | 0.00033 | 0.00504 | 2 | 1053 | 0.00190 | 0.00658 |
| Colon and Rectum | IDH1 | MUT | 12 | 3020 | 0.00397 | 0.01776 | 1 | 1053 | 0.00095 | 0.02351 |
| Colon and Rectum | IDH2 | MUT | 2 | 3020 | 0.00066 | 0.00281 | 0 | 1053 | 0.00000 | 0.00290 |
| Colon and Rectum | JAK1 | MUT | 1 | 3020 | 0.00033 | 0.00026 | 9 | 1053 | 0.00855 | 0.00755 |
| Colon and Rectum | JAK2 | MUT | 1 | 3020 | 0.00033 | 0.00066 | 0 | 1053 | 0.00000 | 0.00019 |
| Colon and Rectum | KIT | AMP | 1 | 3020 | 0.00033 | 0.00675 | 1 | 1053 | 0.00095 | 0.00832 |
| Colon and Rectum | KIT | MUT | 0 | 3020 | 0.00000 | 0.00675 | 2 | 1053 | 0.00190 | 0.01335 |
| Colon and Rectum | KRAS | AMP | 25 | 3020 | 0.00828 | 0.01614 | 12 | 1053 | 0.01140 | 0.01809 |
| Colon and Rectum | KRAS | MUT | 1417 | 3020 | 0.46921 | 0.17580 | 477 | 1053 | 0.45299 | 0.15257 |
| Colon and Rectum | MAP2K1 | MUT | 23 | 3020 | 0.00762 | 0.00412 | 12 | 1053 | 0.01140 | 0.00629 |
| Colon and Rectum | MAP2K2 | MUT | 0 | 3020 | 0.00000 | 0.00022 | 0 | 1053 | 0.00000 | 0.00058 |
| Colon and Rectum | MAP2K4 | MUT | 4 | 3020 | 0.00132 | 0.00092 | 13 | 1053 | 0.01235 | 0.00687 |
| Colon and Rectum | MAPK1 | MUT | 0 | 3020 | 0.00000 | 0.00079 | 0 | 1053 | 0.00000 | 0.00116 |
| Colon and Rectum | MDM2 | AMP | 5 | 3020 | 0.00166 | 0.02772 | 3 | 1053 | 0.00285 | 0.03696 |
| Colon and Rectum | MET | AMP | 11 | 3020 | 0.00364 | 0.00693 | 11 | 1053 | 0.01045 | 0.00948 |
| Colon and Rectum | MET | MUT | 0 | 3020 | 0.00000 | 0.00355 | 0 | 1053 | 0.00000 | 0.00300 |
| Colon and Rectum | MTOR | MUT | 33 | 3020 | 0.01093 | 0.00390 | 13 | 1053 | 0.01235 | 0.00619 |
| Colon and Rectum | MYC | AMP | 116 | 3020 | 0.03841 | 0.04066 | 43 | 1053 | 0.04084 | 0.03957 |
| Colon and Rectum | MYCN | AMP | 2 | 3020 | 0.00066 | 0.00325 | 1 | 1053 | 0.00095 | 0.00484 |
| Colon and Rectum | MYD88 | MUT | 0 | 3020 | 0.00000 | 0.00118 | 1 | 1053 | 0.00095 | 0.00145 |
| Colon and Rectum | NOTCH1 | FUSION | 0 | 3020 | 0.00000 | 0.00018 | 0 | 1053 | 0.00000 | 0.00048 |
| Colon and Rectum | NRAS | MUT | 125 | 3020 | 0.04139 | 0.02048 | 37 | 1053 | 0.03514 | 0.02070 |
| Colon and Rectum | NRG1 | FUSION | 1 | 3020 | 0.00033 | 0.00039 | 0 | 1053 | 0.00000 | 0.00039 |
| Colon and Rectum | NTRK1 | FUSION | 3 | 3020 | 0.00099 | 0.00026 | 3 | 1053 | 0.00285 | 0.00097 |
| Colon and Rectum | NTRK3 | FUSION | 2 | 3020 | 0.00066 | 0.00066 | 1 | 1053 | 0.00095 | 0.00097 |
| Colon and Rectum | PDGFRA | AMP | 1 | 3020 | 0.00033 | 0.00627 | 1 | 1053 | 0.00095 | 0.00851 |
| Colon and Rectum | PDGFRA | MUT | 0 | 3020 | 0.00000 | 0.00048 | 0 | 1053 | 0.00000 | 0.00223 |
| Colon and Rectum | PIK3CA | AMP | 5 | 3020 | 0.00166 | 0.02298 | 1 | 1053 | 0.00095 | 0.00697 |
| Colon and Rectum | PIK3CA | MUT | 482 | 3020 | 0.15960 | 0.12387 | 182 | 1053 | 0.17284 | 0.11474 |
| Colon and Rectum | POLE | MUT | 7 | 3020 | 0.00232 | 0.00079 | 7 | 1053 | 0.00665 | 0.00252 |
| Colon and Rectum | PRKACA | FUSION | 0 | 3020 | 0.00000 | 0.00039 | 0 | 1053 | 0.00000 | 0.00087 |
| Colon and Rectum | PTEN | HOMDEL | 85 | 3020 | 0.02815 | 0.03228 | 15 | 1053 | 0.01425 | 0.02390 |
| Colon and Rectum | PTEN | MUT | 107 | 3020 | 0.03543 | 0.06536 | 59 | 1053 | 0.05603 | 0.05563 |
| Colon and Rectum | RAF1 | MUT | 13 | 3020 | 0.00430 | 0.00154 | 5 | 1053 | 0.00475 | 0.00261 |
| Colon and Rectum | RB1 | HOMDEL | 7 | 3020 | 0.00232 | 0.02298 | 1 | 1053 | 0.00095 | 0.01896 |
| Colon and Rectum | RB1 | MUT | 21 | 3020 | 0.00695 | 0.04053 | 11 | 1053 | 0.01045 | 0.04092 |
| Colon and Rectum | RET | FUSION | 1 | 3020 | 0.00033 | 0.00219 | 1 | 1053 | 0.00095 | 0.00281 |
| Colon and Rectum | RET | MUT | 4 | 3020 | 0.00132 | 0.00175 | 2 | 1053 | 0.00190 | 0.00213 |
| Colon and Rectum | RIT1 | MUT | 0 | 3020 | 0.00000 | 0.00105 | 1 | 1053 | 0.00095 | 0.00126 |
| Colon and Rectum | ROS1 | FUSION | 0 | 3020 | 0.00000 | 0.00149 | 0 | 1053 | 0.00000 | 0.00358 |
| Colon and Rectum | ROS1 | MUT | 0 | 3020 | 0.00000 | 0.00009 | 0 | 1053 | 0.00000 | 0.00019 |
| Colon and Rectum | SF3B1 | MUT | 6 | 3020 | 0.00199 | 0.00807 | 0 | 1053 | 0.00000 | 0.00803 |
| Colon and Rectum | SMO | MUT | 0 | 3020 | 0.00000 | 0.00013 | 0 | 1053 | 0.00000 | 0.00039 |
| Colon and Rectum | SPOP | MUT | 0 | 3020 | 0.00000 | 0.00355 | 0 | 1053 | 0.00000 | 0.00793 |
| Colon and Rectum | TERT | FUSION | 0 | 3020 | 0.00000 | 0.00009 | 0 | 1053 | 0.00000 | 0.00000 |
| Colon and Rectum | TERT | MUT | 28 | 3020 | 0.00927 | 0.09795 | 16 | 1053 | 0.01519 | 0.12307 |
| Colon and Rectum | TP53 | HOMDEL | 44 | 3020 | 0.01457 | 0.01276 | 5 | 1053 | 0.00475 | 0.00900 |
| Colon and Rectum | TP53 | MUT | 2200 | 3020 | 0.72848 | 0.52404 | 731 | 1053 | 0.69421 | 0.41428 |
| Endometrium | AKT1 | MUT | 30 | 919 | 0.03264 | 0.01026 | 8 | 210 | 0.03810 | 0.01045 |
| Endometrium | ALK | AMP | 0 | 919 | 0.00000 | 0.00018 | 0 | 210 | 0.00000 | 0.00039 |
| Endometrium | ALK | FUSION | 0 | 919 | 0.00000 | 0.00570 | 0 | 210 | 0.00000 | 0.00474 |
| Endometrium | ALK | MUT | 0 | 919 | 0.00000 | 0.00061 | 0 | 210 | 0.00000 | 0.00184 |
| Endometrium | AR | AMP | 0 | 919 | 0.00000 | 0.00601 | 0 | 210 | 0.00000 | 0.00842 |
| Endometrium | AR | MUT | 0 | 919 | 0.00000 | 0.00136 | 0 | 210 | 0.00000 | 0.00213 |
| Endometrium | ARAF | MUT | 0 | 919 | 0.00000 | 0.00039 | 0 | 210 | 0.00000 | 0.00068 |
| Endometrium | BRAF | AMP | 0 | 919 | 0.00000 | 0.00250 | 0 | 210 | 0.00000 | 0.00164 |
| Endometrium | BRAF | FUSION | 0 | 919 | 0.00000 | 0.00118 | 0 | 210 | 0.00000 | 0.00368 |
| Endometrium | BRAF | MUT | 7 | 919 | 0.00762 | 0.04106 | 2 | 210 | 0.00952 | 0.04247 |
| Endometrium | CCND1 | AMP | 4 | 919 | 0.00435 | 0.04299 | 4 | 210 | 0.01905 | 0.04189 |
| Endometrium | CDK4 | AMP | 3 | 919 | 0.00326 | 0.01921 | 1 | 210 | 0.00476 | 0.02632 |
| Endometrium | CDK4 | MUT | 0 | 919 | 0.00000 | 0.00061 | 0 | 210 | 0.00000 | 0.00116 |
| Endometrium | CDK6 | AMP | 5 | 919 | 0.00544 | 0.00697 | 0 | 210 | 0.00000 | 0.00590 |
| Endometrium | CDKN2A | HOMDEL | 17 | 919 | 0.01850 | 0.12234 | 3 | 210 | 0.01429 | 0.07498 |
| Endometrium | CDKN2A | MUT | 13 | 919 | 0.01415 | 0.04917 | 4 | 210 | 0.01905 | 0.03599 |
| Endometrium | CTNNB1 | MUT | 162 | 919 | 0.17628 | 0.02601 | 27 | 210 | 0.12857 | 0.02254 |
| Endometrium | EGFR | AMP | 2 | 919 | 0.00218 | 0.02996 | 0 | 210 | 0.00000 | 0.03154 |
| Endometrium | EGFR | MUT | 4 | 919 | 0.00435 | 0.02649 | 1 | 210 | 0.00476 | 0.04112 |
| Endometrium | ERBB2 | AMP | 54 | 919 | 0.05876 | 0.03202 | 17 | 210 | 0.08095 | 0.03938 |
| Endometrium | ERBB2 | MUT | 16 | 919 | 0.01741 | 0.01689 | 8 | 210 | 0.03810 | 0.01945 |
| Endometrium | ERBB3 | MUT | 15 | 919 | 0.01632 | 0.00561 | 8 | 210 | 0.03810 | 0.00987 |
| Endometrium | ERBB4 | MUT | 0 | 919 | 0.00000 | 0.00022 | 0 | 210 | 0.00000 | 0.00155 |
| Endometrium | ESR1 | MUT | 20 | 919 | 0.02176 | 0.00965 | 7 | 210 | 0.03333 | 0.01006 |
| Endometrium | EZH2 | MUT | 0 | 919 | 0.00000 | 0.00140 | 2 | 210 | 0.00952 | 0.00281 |
| Endometrium | FGFR1 | AMP | 19 | 919 | 0.02067 | 0.02816 | 9 | 210 | 0.04286 | 0.02554 |
| Endometrium | FGFR1 | FUSION | 0 | 919 | 0.00000 | 0.00013 | 0 | 210 | 0.00000 | 0.00039 |
| Endometrium | FGFR2 | AMP | 0 | 919 | 0.00000 | 0.00347 | 1 | 210 | 0.00476 | 0.00358 |

| | | | | | | | | | | |
|---|---|---|---|---|---|---|---|---|---|---|
| Endometrium | FGFR2 | FUSION | 0 | 919 | 0.00000 | 0.00083 | 0 | 210 | 0.00000 | 0.00300 |
| Endometrium | FGFR2 | MUT | 64 | 919 | 0.06964 | 0.00531 | 11 | 210 | 0.05238 | 0.00435 |
| Endometrium | FGFR3 | AMP | 3 | 919 | 0.00326 | 0.00263 | 2 | 210 | 0.00952 | 0.00387 |
| Endometrium | FGFR3 | FUSION | 1 | 919 | 0.00109 | 0.00294 | 1 | 210 | 0.00476 | 0.00271 |
| Endometrium | FGFR3 | MUT | 2 | 919 | 0.00218 | 0.00588 | 0 | 210 | 0.00000 | 0.01200 |
| Endometrium | FGFR4 | AMP | 0 | 919 | 0.00000 | 0.00075 | 0 | 210 | 0.00000 | 0.00252 |
| Endometrium | GNA11 | MUT | 0 | 919 | 0.00000 | 0.00105 | 0 | 210 | 0.00000 | 0.00184 |
| Endometrium | GNAQ | MUT | 0 | 919 | 0.00000 | 0.00096 | 0 | 210 | 0.00000 | 0.00281 |
| Endometrium | HRAS | MUT | 0 | 919 | 0.00000 | 0.00504 | 0 | 210 | 0.00000 | 0.00658 |
| Endometrium | IDH1 | MUT | 1 | 919 | 0.00109 | 0.01776 | 0 | 210 | 0.00000 | 0.02351 |
| Endometrium | IDH2 | MUT | 0 | 919 | 0.00000 | 0.00281 | 0 | 210 | 0.00000 | 0.00290 |
| Endometrium | JAK1 | MUT | 0 | 919 | 0.00000 | 0.00026 | 19 | 210 | 0.09048 | 0.00755 |
| Endometrium | JAK2 | MUT | 3 | 919 | 0.00326 | 0.00066 | 0 | 210 | 0.00000 | 0.00019 |
| Endometrium | KIT | AMP | 5 | 919 | 0.00544 | 0.00675 | 0 | 210 | 0.00000 | 0.00832 |
| Endometrium | KIT | MUT | 0 | 919 | 0.00000 | 0.00675 | 0 | 210 | 0.00000 | 0.01335 |
| Endometrium | KRAS | AMP | 6 | 919 | 0.00653 | 0.01614 | 3 | 210 | 0.01429 | 0.01809 |
| Endometrium | KRAS | MUT | 158 | 919 | 0.17193 | 0.17580 | 39 | 210 | 0.18571 | 0.15257 |
| Endometrium | MAP2K1 | MUT | 2 | 919 | 0.00218 | 0.00412 | 1 | 210 | 0.00476 | 0.00629 |
| Endometrium | MAP2K2 | MUT | 0 | 919 | 0.00000 | 0.00022 | 0 | 210 | 0.00000 | 0.00058 |
| Endometrium | MAP2K4 | MUT | 0 | 919 | 0.00000 | 0.00092 | 2 | 210 | 0.00952 | 0.00687 |
| Endometrium | MAPK1 | MUT | 0 | 919 | 0.00000 | 0.00079 | 0 | 210 | 0.00000 | 0.00116 |
| Endometrium | MDM2 | AMP | 5 | 919 | 0.00544 | 0.02772 | 3 | 210 | 0.01429 | 0.03696 |
| Endometrium | MET | AMP | 2 | 919 | 0.00218 | 0.00693 | 0 | 210 | 0.00000 | 0.00948 |
| Endometrium | MET | MUT | 0 | 919 | 0.00000 | 0.00355 | 0 | 210 | 0.00000 | 0.00300 |
| Endometrium | MTOR | MUT | 7 | 919 | 0.00762 | 0.00390 | 5 | 210 | 0.02381 | 0.00619 |
| Endometrium | MYC | AMP | 34 | 919 | 0.03700 | 0.04066 | 12 | 210 | 0.05714 | 0.03957 |
| Endometrium | MYCN | AMP | 5 | 919 | 0.00544 | 0.00325 | 1 | 210 | 0.00476 | 0.00484 |
| Endometrium | MYD88 | MUT | 0 | 919 | 0.00000 | 0.00118 | 0 | 210 | 0.00000 | 0.00145 |
| Endometrium | NOTCH1 | FUSION | 0 | 919 | 0.00000 | 0.00018 | 0 | 210 | 0.00000 | 0.00048 |
| Endometrium | NRAS | MUT | 13 | 919 | 0.01415 | 0.02048 | 3 | 210 | 0.01429 | 0.02070 |
| Endometrium | NRG1 | FUSION | 0 | 919 | 0.00000 | 0.00039 | 0 | 210 | 0.00000 | 0.00039 |
| Endometrium | NTRK1 | FUSION | 0 | 919 | 0.00000 | 0.00026 | 0 | 210 | 0.00000 | 0.00097 |
| Endometrium | NTRK3 | FUSION | 0 | 919 | 0.00000 | 0.00066 | 0 | 210 | 0.00000 | 0.00097 |
| Endometrium | PDGFRA | AMP | 3 | 919 | 0.00326 | 0.00627 | 0 | 210 | 0.00000 | 0.00851 |
| Endometrium | PDGFRA | MUT | 0 | 919 | 0.00000 | 0.00048 | 1 | 210 | 0.00476 | 0.00223 |
| Endometrium | PIK3CA | AMP | 21 | 919 | 0.02285 | 0.02298 | 3 | 210 | 0.01429 | 0.00697 |
| Endometrium | PIK3CA | MUT | 338 | 919 | 0.36779 | 0.12387 | 85 | 210 | 0.40476 | 0.11474 |
| Endometrium | POLE | MUT | 6 | 919 | 0.00653 | 0.00079 | 6 | 210 | 0.02857 | 0.00252 |
| Endometrium | PRKACA | FUSION | 0 | 919 | 0.00000 | 0.00039 | 0 | 210 | 0.00000 | 0.00087 |
| Endometrium | PTEN | HOMDEL | 21 | 919 | 0.02285 | 0.03228 | 0 | 210 | 0.00000 | 0.02390 |
| Endometrium | PTEN | MUT | 338 | 919 | 0.36779 | 0.06536 | 66 | 210 | 0.31429 | 0.05563 |
| Endometrium | RAF1 | MUT | 2 | 919 | 0.00218 | 0.00154 | 0 | 210 | 0.00000 | 0.00261 |
| Endometrium | RB1 | HOMDEL | 15 | 919 | 0.01632 | 0.02298 | 3 | 210 | 0.01429 | 0.01896 |
| Endometrium | RB1 | MUT | 29 | 919 | 0.03156 | 0.04053 | 8 | 210 | 0.03810 | 0.04092 |
| Endometrium | RET | FUSION | 0 | 919 | 0.00000 | 0.00219 | 0 | 210 | 0.00000 | 0.00281 |
| Endometrium | RET | MUT | 0 | 919 | 0.00000 | 0.00175 | 0 | 210 | 0.00000 | 0.00213 |
| Endometrium | RIT1 | MUT | 2 | 919 | 0.00218 | 0.00105 | 1 | 210 | 0.00476 | 0.00126 |
| Endometrium | ROS1 | FUSION | 0 | 919 | 0.00000 | 0.00149 | 0 | 210 | 0.00000 | 0.00358 |
| Endometrium | ROS1 | MUT | 0 | 919 | 0.00000 | 0.00009 | 0 | 210 | 0.00000 | 0.00019 |
| Endometrium | SF3B1 | MUT | 2 | 919 | 0.00218 | 0.00807 | 1 | 210 | 0.00476 | 0.00803 |
| Endometrium | SMO | MUT | 0 | 919 | 0.00000 | 0.00013 | 0 | 210 | 0.00000 | 0.00039 |
| Endometrium | SPOP | MUT | 0 | 919 | 0.00000 | 0.00355 | 7 | 210 | 0.03333 | 0.00793 |
| Endometrium | TERT | FUSION | 0 | 919 | 0.00000 | 0.00009 | 0 | 210 | 0.00000 | 0.00000 |
| Endometrium | TERT | MUT | 12 | 919 | 0.01306 | 0.09795 | 1 | 210 | 0.00476 | 0.12307 |
| Endometrium | TP53 | HOMDEL | 3 | 919 | 0.00326 | 0.01276 | 0 | 210 | 0.00000 | 0.00900 |
| Endometrium | TP53 | MUT | 495 | 919 | 0.53863 | 0.52404 | 112 | 210 | 0.53333 | 0.41428 |
| Esophagus_Stomach | AKT1 | MUT | 2 | 994 | 0.00201 | 0.01026 | 1 | 315 | 0.00317 | 0.01045 |
| Esophagus_Stomach | ALK | AMP | 0 | 994 | 0.00000 | 0.00018 | 0 | 315 | 0.00000 | 0.00039 |
| Esophagus_Stomach | ALK | FUSION | 0 | 994 | 0.00000 | 0.00570 | 0 | 315 | 0.00000 | 0.00474 |
| Esophagus_Stomach | ALK | MUT | 0 | 994 | 0.00000 | 0.00061 | 1 | 315 | 0.00317 | 0.00184 |
| Esophagus_Stomach | AR | AMP | 0 | 994 | 0.00000 | 0.00601 | 0 | 315 | 0.00000 | 0.00842 |
| Esophagus_Stomach | AR | MUT | 0 | 994 | 0.00000 | 0.00136 | 1 | 315 | 0.00317 | 0.00213 |
| Esophagus_Stomach | ARAF | MUT | 0 | 994 | 0.00000 | 0.00039 | 0 | 315 | 0.00000 | 0.00068 |
| Esophagus_Stomach | BRAF | AMP | 1 | 994 | 0.00101 | 0.00250 | 0 | 315 | 0.00000 | 0.00164 |
| Esophagus_Stomach | BRAF | FUSION | 0 | 994 | 0.00000 | 0.00118 | 0 | 315 | 0.00000 | 0.00368 |
| Esophagus_Stomach | BRAF | MUT | 13 | 994 | 0.01308 | 0.04106 | 3 | 315 | 0.00952 | 0.04247 |
| Esophagus_Stomach | CCND1 | AMP | 112 | 994 | 0.11268 | 0.04299 | 21 | 315 | 0.06667 | 0.04189 |
| Esophagus_Stomach | CDK4 | AMP | 11 | 994 | 0.01107 | 0.01921 | 4 | 315 | 0.01270 | 0.02632 |
| Esophagus_Stomach | CDK4 | MUT | 0 | 994 | 0.00000 | 0.00061 | 0 | 315 | 0.00000 | 0.00116 |
| Esophagus_Stomach | CDK6 | AMP | 59 | 994 | 0.05936 | 0.00697 | 12 | 315 | 0.03810 | 0.00590 |
| Esophagus_Stomach | CDKN2A | HOMDEL | 171 | 994 | 0.17203 | 0.12234 | 17 | 315 | 0.05397 | 0.07498 |
| Esophagus_Stomach | CDKN2A | MUT | 94 | 994 | 0.09457 | 0.04917 | 26 | 315 | 0.08254 | 0.03599 |
| Esophagus_Stomach | CTNNB1 | MUT | 14 | 994 | 0.01408 | 0.02601 | 4 | 315 | 0.01270 | 0.02254 |
| Esophagus_Stomach | EGFR | AMP | 60 | 994 | 0.06036 | 0.02996 | 19 | 315 | 0.06032 | 0.03154 |
| Esophagus_Stomach | EGFR | MUT | 0 | 994 | 0.00000 | 0.02649 | 1 | 315 | 0.00317 | 0.04112 |
| Esophagus_Stomach | ERBB2 | AMP | 106 | 994 | 0.10664 | 0.03202 | 65 | 315 | 0.20635 | 0.03938 |

| | | | | | | | | | | |
|---|---|---|---|---|---|---|---|---|---|---|
| Esophagus_Stomach | ERBB2 | MUT | 29 | 994 | 0.02918 | 0.01689 | 11 | 315 | 0.03492 | 0.01945 |
| Esophagus_Stomach | ERBB3 | MUT | 8 | 994 | 0.00805 | 0.00561 | 16 | 315 | 0.05079 | 0.00987 |
| Esophagus_Stomach | ERBB4 | MUT | 0 | 994 | 0.00000 | 0.00022 | 2 | 315 | 0.00635 | 0.00155 |
| Esophagus_Stomach | ESR1 | MUT | 0 | 994 | 0.00000 | 0.00965 | 0 | 315 | 0.00000 | 0.01006 |
| Esophagus_Stomach | EZH2 | MUT | 0 | 994 | 0.00000 | 0.00140 | 0 | 315 | 0.00000 | 0.00281 |
| Esophagus_Stomach | FGFR1 | AMP | 16 | 994 | 0.01610 | 0.02816 | 5 | 315 | 0.01587 | 0.02554 |
| Esophagus_Stomach | FGFR1 | FUSION | 0 | 994 | 0.00000 | 0.00013 | 0 | 315 | 0.00000 | 0.00039 |
| Esophagus_Stomach | FGFR2 | AMP | 27 | 994 | 0.02716 | 0.00347 | 9 | 315 | 0.02857 | 0.00358 |
| Esophagus_Stomach | FGFR2 | FUSION | 0 | 994 | 0.00000 | 0.00083 | 0 | 315 | 0.00000 | 0.00300 |
| Esophagus_Stomach | FGFR2 | MUT | 3 | 994 | 0.00302 | 0.00531 | 2 | 315 | 0.00635 | 0.00435 |
| Esophagus_Stomach | FGFR3 | AMP | 3 | 994 | 0.00302 | 0.00263 | 1 | 315 | 0.00317 | 0.00387 |
| Esophagus_Stomach | FGFR3 | FUSION | 3 | 994 | 0.00302 | 0.00294 | 0 | 315 | 0.00000 | 0.00271 |
| Esophagus_Stomach | FGFR3 | MUT | 0 | 994 | 0.00000 | 0.00588 | 0 | 315 | 0.00000 | 0.01200 |
| Esophagus_Stomach | FGFR4 | AMP | 0 | 994 | 0.00000 | 0.00075 | 0 | 315 | 0.00000 | 0.00252 |
| Esophagus_Stomach | GNA11 | MUT | 0 | 994 | 0.00000 | 0.00105 | 0 | 315 | 0.00000 | 0.00184 |
| Esophagus_Stomach | GNAQ | MUT | 0 | 994 | 0.00000 | 0.00096 | 0 | 315 | 0.00000 | 0.00281 |
| Esophagus_Stomach | HRAS | MUT | 3 | 994 | 0.00302 | 0.00504 | 1 | 315 | 0.00317 | 0.00658 |
| Esophagus_Stomach | IDH1 | MUT | 1 | 994 | 0.00101 | 0.01776 | 0 | 315 | 0.00000 | 0.02351 |
| Esophagus_Stomach | IDH2 | MUT | 0 | 994 | 0.00000 | 0.00281 | 0 | 315 | 0.00000 | 0.00290 |
| Esophagus_Stomach | JAK1 | MUT | 0 | 994 | 0.00000 | 0.00026 | 5 | 315 | 0.01587 | 0.00755 |
| Esophagus_Stomach | JAK2 | MUT | 1 | 994 | 0.00101 | 0.00066 | 0 | 315 | 0.00000 | 0.00019 |
| Esophagus_Stomach | KIT | AMP | 1 | 994 | 0.00101 | 0.00675 | 0 | 315 | 0.00000 | 0.00832 |
| Esophagus_Stomach | KIT | MUT | 2 | 994 | 0.00201 | 0.00675 | 0 | 315 | 0.00000 | 0.01335 |
| Esophagus_Stomach | KRAS | AMP | 102 | 994 | 0.10262 | 0.01614 | 26 | 315 | 0.08254 | 0.01809 |
| Esophagus_Stomach | KRAS | MUT | 64 | 994 | 0.06439 | 0.17580 | 24 | 315 | 0.07619 | 0.15257 |
| Esophagus_Stomach | MAP2K1 | MUT | 5 | 994 | 0.00503 | 0.00412 | 1 | 315 | 0.00317 | 0.00629 |
| Esophagus_Stomach | MAP2K2 | MUT | 0 | 994 | 0.00000 | 0.00022 | 0 | 315 | 0.00000 | 0.00058 |
| Esophagus_Stomach | MAP2K4 | MUT | 0 | 994 | 0.00000 | 0.00092 | 1 | 315 | 0.00317 | 0.00687 |
| Esophagus_Stomach | MAPK1 | MUT | 1 | 994 | 0.00101 | 0.00079 | 0 | 315 | 0.00000 | 0.00116 |
| Esophagus_Stomach | MDM2 | AMP | 33 | 994 | 0.03320 | 0.02772 | 14 | 315 | 0.04444 | 0.03696 |
| Esophagus_Stomach | MET | AMP | 32 | 994 | 0.03219 | 0.00693 | 8 | 315 | 0.02540 | 0.00948 |
| Esophagus_Stomach | MET | MUT | 0 | 994 | 0.00000 | 0.00355 | 0 | 315 | 0.00000 | 0.00300 |
| Esophagus_Stomach | MTOR | MUT | 0 | 994 | 0.00000 | 0.00390 | 1 | 315 | 0.00317 | 0.00619 |
| Esophagus_Stomach | MYC | AMP | 76 | 994 | 0.07646 | 0.04066 | 14 | 315 | 0.04444 | 0.03957 |
| Esophagus_Stomach | MYCN | AMP | 2 | 994 | 0.00201 | 0.00325 | 0 | 315 | 0.00000 | 0.00484 |
| Esophagus_Stomach | MYD88 | MUT | 1 | 994 | 0.00101 | 0.00118 | 0 | 315 | 0.00000 | 0.00145 |
| Esophagus_Stomach | NOTCH1 | FUSION | 0 | 994 | 0.00000 | 0.00018 | 1 | 315 | 0.00317 | 0.00048 |
| Esophagus_Stomach | NRAS | MUT | 3 | 994 | 0.00302 | 0.02048 | 2 | 315 | 0.00635 | 0.02070 |
| Esophagus_Stomach | NRG1 | FUSION | 0 | 994 | 0.00000 | 0.00039 | 0 | 315 | 0.00000 | 0.00039 |
| Esophagus_Stomach | NTRK1 | FUSION | 0 | 994 | 0.00000 | 0.00026 | 0 | 315 | 0.00000 | 0.00097 |
| Esophagus_Stomach | NTRK3 | FUSION | 0 | 994 | 0.00000 | 0.00066 | 0 | 315 | 0.00000 | 0.00097 |
| Esophagus_Stomach | PDGFRA | AMP | 1 | 994 | 0.00101 | 0.00627 | 0 | 315 | 0.00000 | 0.00851 |
| Esophagus_Stomach | PDGFRA | MUT | 0 | 994 | 0.00000 | 0.00048 | 0 | 315 | 0.00000 | 0.00223 |
| Esophagus_Stomach | PIK3CA | AMP | 25 | 994 | 0.02515 | 0.02298 | 0 | 315 | 0.00000 | 0.00697 |
| Esophagus_Stomach | PIK3CA | MUT | 75 | 994 | 0.07545 | 0.12387 | 19 | 315 | 0.06032 | 0.11474 |
| Esophagus_Stomach | POLE | MUT | 0 | 994 | 0.00000 | 0.00079 | 0 | 315 | 0.00000 | 0.00252 |
| Esophagus_Stomach | PRKACA | FUSION | 0 | 994 | 0.00000 | 0.00039 | 0 | 315 | 0.00000 | 0.00087 |
| Esophagus_Stomach | PTEN | HOMDEL | 13 | 994 | 0.01308 | 0.03228 | 2 | 315 | 0.00635 | 0.02390 |
| Esophagus_Stomach | PTEN | MUT | 20 | 994 | 0.02012 | 0.06536 | 11 | 315 | 0.03492 | 0.05563 |
| Esophagus_Stomach | RAF1 | MUT | 0 | 994 | 0.00000 | 0.00154 | 1 | 315 | 0.00317 | 0.00261 |
| Esophagus_Stomach | RB1 | HOMDEL | 13 | 994 | 0.01308 | 0.02298 | 3 | 315 | 0.00952 | 0.01896 |
| Esophagus_Stomach | RB1 | MUT | 17 | 994 | 0.01710 | 0.04053 | 4 | 315 | 0.01270 | 0.04092 |
| Esophagus_Stomach | RET | FUSION | 0 | 994 | 0.00000 | 0.00219 | 1 | 315 | 0.00317 | 0.00281 |
| Esophagus_Stomach | RET | MUT | 0 | 994 | 0.00000 | 0.00175 | 1 | 315 | 0.00317 | 0.00213 |
| Esophagus_Stomach | RIT1 | MUT | 1 | 994 | 0.00101 | 0.00105 | 0 | 315 | 0.00000 | 0.00126 |
| Esophagus_Stomach | ROS1 | FUSION | 0 | 994 | 0.00000 | 0.00149 | 1 | 315 | 0.00317 | 0.00358 |
| Esophagus_Stomach | ROS1 | MUT | 0 | 994 | 0.00000 | 0.00009 | 0 | 315 | 0.00000 | 0.00019 |
| Esophagus_Stomach | SF3B1 | MUT | 0 | 994 | 0.00000 | 0.00807 | 0 | 315 | 0.00000 | 0.00803 |
| Esophagus_Stomach | SMO | MUT | 0 | 994 | 0.00000 | 0.00013 | 0 | 315 | 0.00000 | 0.00039 |
| Esophagus_Stomach | SPOP | MUT | 0 | 994 | 0.00000 | 0.00355 | 1 | 315 | 0.00317 | 0.00793 |
| Esophagus_Stomach | TERT | FUSION | 0 | 994 | 0.00000 | 0.00009 | 0 | 315 | 0.00000 | 0.00000 |
| Esophagus_Stomach | TERT | MUT | 4 | 994 | 0.00402 | 0.09795 | 2 | 315 | 0.00635 | 0.12307 |
| Esophagus_Stomach | TP53 | HOMDEL | 8 | 994 | 0.00805 | 0.01276 | 1 | 315 | 0.00317 | 0.00900 |
| Esophagus_Stomach | TP53 | MUT | 730 | 994 | 0.73441 | 0.52404 | 220 | 315 | 0.69841 | 0.41428 |
| Head and Neck | AKT1 | MUT | 6 | 746 | 0.00804 | 0.01026 | 2 | 282 | 0.00709 | 0.01045 |
| Head and Neck | ALK | AMP | 0 | 746 | 0.00000 | 0.00018 | 0 | 282 | 0.00000 | 0.00039 |
| Head and Neck | ALK | FUSION | 0 | 746 | 0.00000 | 0.00570 | 1 | 282 | 0.00355 | 0.00474 |
| Head and Neck | ALK | MUT | 0 | 746 | 0.00000 | 0.00061 | 0 | 282 | 0.00000 | 0.00184 |
| Head and Neck | AR | AMP | 0 | 746 | 0.00000 | 0.00601 | 1 | 282 | 0.00355 | 0.00842 |
| Head and Neck | AR | MUT | 0 | 746 | 0.00000 | 0.00136 | 0 | 282 | 0.00000 | 0.00213 |
| Head and Neck | ARAF | MUT | 0 | 746 | 0.00000 | 0.00039 | 0 | 282 | 0.00000 | 0.00068 |
| Head and Neck | BRAF | AMP | 1 | 746 | 0.00134 | 0.00250 | 0 | 282 | 0.00000 | 0.00164 |
| Head and Neck | BRAF | FUSION | 0 | 746 | 0.00000 | 0.00118 | 0 | 282 | 0.00000 | 0.00368 |
| Head and Neck | BRAF | MUT | 8 | 746 | 0.01072 | 0.04106 | 1 | 282 | 0.00355 | 0.04247 |
| Head and Neck | CCND1 | AMP | 116 | 746 | 0.15550 | 0.04299 | 22 | 282 | 0.07801 | 0.04189 |
| Head and Neck | CDK4 | AMP | 3 | 746 | 0.00402 | 0.01921 | 3 | 282 | 0.01064 | 0.02632 |

| | | | | | | | | | | |
|---|---|---|---|---|---|---|---|---|---|---|
| Head and Neck | CDK4 | MUT | 0 | 746 | 0.00000 | 0.00061 | 0 | 282 | 0.00000 | 0.00116 |
| Head and Neck | CDK6 | AMP | 5 | 746 | 0.00670 | 0.00697 | 3 | 282 | 0.01064 | 0.00590 |
| Head and Neck | CDKN2A | HOMDEL | 126 | 746 | 0.16890 | 0.12234 | 28 | 282 | 0.09929 | 0.07498 |
| Head and Neck | CDKN2A | MUT | 92 | 746 | 0.12332 | 0.04917 | 17 | 282 | 0.06028 | 0.03599 |
| Head and Neck | CTNNB1 | MUT | 4 | 746 | 0.00536 | 0.02601 | 0 | 282 | 0.00000 | 0.02254 |
| Head and Neck | EGFR | AMP | 28 | 746 | 0.03753 | 0.02996 | 13 | 282 | 0.04610 | 0.03154 |
| Head and Neck | EGFR | MUT | 3 | 746 | 0.00402 | 0.02649 | 0 | 282 | 0.00000 | 0.04112 |
| Head and Neck | ERBB2 | AMP | 14 | 746 | 0.01877 | 0.03202 | 12 | 282 | 0.04255 | 0.03938 |
| Head and Neck | ERBB2 | MUT | 2 | 746 | 0.00268 | 0.01689 | 4 | 282 | 0.01418 | 0.01945 |
| Head and Neck | ERBB3 | MUT | 5 | 746 | 0.00670 | 0.00561 | 2 | 282 | 0.00709 | 0.00987 |
| Head and Neck | ERBB4 | MUT | 0 | 746 | 0.00000 | 0.00022 | 0 | 282 | 0.00000 | 0.00155 |
| Head and Neck | ESR1 | MUT | 0 | 746 | 0.00000 | 0.00965 | 0 | 282 | 0.00000 | 0.01006 |
| Head and Neck | EZH2 | MUT | 0 | 746 | 0.00000 | 0.00140 | 1 | 282 | 0.00355 | 0.00281 |
| Head and Neck | FGFR1 | AMP | 9 | 746 | 0.01206 | 0.02816 | 5 | 282 | 0.01773 | 0.02554 |
| Head and Neck | FGFR1 | FUSION | 0 | 746 | 0.00000 | 0.00013 | 0 | 282 | 0.00000 | 0.00039 |
| Head and Neck | FGFR2 | AMP | 1 | 746 | 0.00134 | 0.00347 | 0 | 282 | 0.00000 | 0.00358 |
| Head and Neck | FGFR2 | FUSION | 1 | 746 | 0.00134 | 0.00083 | 0 | 282 | 0.00000 | 0.00300 |
| Head and Neck | FGFR2 | MUT | 3 | 746 | 0.00402 | 0.00531 | 2 | 282 | 0.00709 | 0.00435 |
| Head and Neck | FGFR3 | AMP | 1 | 746 | 0.00134 | 0.00263 | 1 | 282 | 0.00355 | 0.00387 |
| Head and Neck | FGFR3 | FUSION | 2 | 746 | 0.00268 | 0.00294 | 1 | 282 | 0.00355 | 0.00271 |
| Head and Neck | FGFR3 | MUT | 7 | 746 | 0.00938 | 0.00588 | 4 | 282 | 0.01418 | 0.01200 |
| Head and Neck | FGFR4 | AMP | 0 | 746 | 0.00000 | 0.00075 | 0 | 282 | 0.00000 | 0.00252 |
| Head and Neck | GNA11 | MUT | 1 | 746 | 0.00134 | 0.00105 | 0 | 282 | 0.00000 | 0.00184 |
| Head and Neck | GNAQ | MUT | 0 | 746 | 0.00000 | 0.00096 | 0 | 282 | 0.00000 | 0.00281 |
| Head and Neck | HRAS | MUT | 23 | 746 | 0.03083 | 0.00504 | 11 | 282 | 0.03901 | 0.00658 |
| Head and Neck | IDH1 | MUT | 3 | 746 | 0.00402 | 0.01776 | 1 | 282 | 0.00355 | 0.02351 |
| Head and Neck | IDH2 | MUT | 2 | 746 | 0.00268 | 0.00281 | 2 | 282 | 0.00709 | 0.00290 |
| Head and Neck | JAK1 | MUT | 0 | 746 | 0.00000 | 0.00026 | 1 | 282 | 0.00355 | 0.00755 |
| Head and Neck | JAK2 | MUT | 1 | 746 | 0.00134 | 0.00066 | 0 | 282 | 0.00000 | 0.00019 |
| Head and Neck | KIT | AMP | 3 | 746 | 0.00402 | 0.00675 | 7 | 282 | 0.02482 | 0.00832 |
| Head and Neck | KIT | MUT | 1 | 746 | 0.00134 | 0.00675 | 0 | 282 | 0.00000 | 0.01335 |
| Head and Neck | KRAS | AMP | 6 | 746 | 0.00804 | 0.01614 | 1 | 282 | 0.00355 | 0.01809 |
| Head and Neck | KRAS | MUT | 12 | 746 | 0.01609 | 0.17580 | 3 | 282 | 0.01064 | 0.15257 |
| Head and Neck | MAP2K1 | MUT | 1 | 746 | 0.00134 | 0.00412 | 0 | 282 | 0.00000 | 0.00629 |
| Head and Neck | MAP2K2 | MUT | 0 | 746 | 0.00000 | 0.00022 | 0 | 282 | 0.00000 | 0.00058 |
| Head and Neck | MAP2K4 | MUT | 0 | 746 | 0.00000 | 0.00092 | 1 | 282 | 0.00355 | 0.00687 |
| Head and Neck | MAPK1 | MUT | 4 | 746 | 0.00536 | 0.00079 | 4 | 282 | 0.01418 | 0.00116 |
| Head and Neck | MDM2 | AMP | 13 | 746 | 0.01743 | 0.02772 | 5 | 282 | 0.01773 | 0.03696 |
| Head and Neck | MET | AMP | 2 | 746 | 0.00268 | 0.00693 | 0 | 282 | 0.00000 | 0.00948 |
| Head and Neck | MET | MUT | 0 | 746 | 0.00000 | 0.00355 | 0 | 282 | 0.00000 | 0.00300 |
| Head and Neck | MTOR | MUT | 2 | 746 | 0.00268 | 0.00390 | 2 | 282 | 0.00709 | 0.00619 |
| Head and Neck | MYC | AMP | 16 | 746 | 0.02145 | 0.04066 | 4 | 282 | 0.01418 | 0.03957 |
| Head and Neck | MYCN | AMP | 1 | 746 | 0.00134 | 0.00325 | 1 | 282 | 0.00355 | 0.00484 |
| Head and Neck | MYD88 | MUT | 1 | 746 | 0.00134 | 0.00118 | 0 | 282 | 0.00000 | 0.00145 |
| Head and Neck | NOTCH1 | FUSION | 0 | 746 | 0.00000 | 0.00018 | 0 | 282 | 0.00000 | 0.00048 |
| Head and Neck | NRAS | MUT | 4 | 746 | 0.00536 | 0.02048 | 3 | 282 | 0.01064 | 0.02070 |
| Head and Neck | NRG1 | FUSION | 0 | 746 | 0.00000 | 0.00039 | 0 | 282 | 0.00000 | 0.00039 |
| Head and Neck | NTRK1 | FUSION | 0 | 746 | 0.00000 | 0.00026 | 0 | 282 | 0.00000 | 0.00097 |
| Head and Neck | NTRK3 | FUSION | 4 | 746 | 0.00536 | 0.00066 | 4 | 282 | 0.01418 | 0.00097 |
| Head and Neck | PDGFRA | AMP | 3 | 746 | 0.00402 | 0.00627 | 9 | 282 | 0.03191 | 0.00851 |
| Head and Neck | PDGFRA | MUT | 0 | 746 | 0.00000 | 0.00048 | 0 | 282 | 0.00000 | 0.00223 |
| Head and Neck | PIK3CA | AMP | 80 | 746 | 0.10724 | 0.02298 | 9 | 282 | 0.03191 | 0.00697 |
| Head and Neck | PIK3CA | MUT | 93 | 746 | 0.12466 | 0.12387 | 43 | 282 | 0.15248 | 0.11474 |
| Head and Neck | POLE | MUT | 0 | 746 | 0.00000 | 0.00079 | 0 | 282 | 0.00000 | 0.00252 |
| Head and Neck | PRKACA | FUSION | 0 | 746 | 0.00000 | 0.00039 | 0 | 282 | 0.00000 | 0.00087 |
| Head and Neck | PTEN | HOMDEL | 23 | 746 | 0.03083 | 0.03228 | 3 | 282 | 0.01064 | 0.02390 |
| Head and Neck | PTEN | MUT | 19 | 746 | 0.02547 | 0.06536 | 11 | 282 | 0.03901 | 0.05563 |
| Head and Neck | RAF1 | MUT | 0 | 746 | 0.00000 | 0.00154 | 1 | 282 | 0.00355 | 0.00261 |
| Head and Neck | RB1 | HOMDEL | 7 | 746 | 0.00938 | 0.02298 | 3 | 282 | 0.01064 | 0.01896 |
| Head and Neck | RB1 | MUT | 22 | 746 | 0.02949 | 0.04053 | 9 | 282 | 0.03191 | 0.04092 |
| Head and Neck | RET | FUSION | 2 | 746 | 0.00268 | 0.00219 | 0 | 282 | 0.00000 | 0.00281 |
| Head and Neck | RET | MUT | 0 | 746 | 0.00000 | 0.00175 | 1 | 282 | 0.00355 | 0.00213 |
| Head and Neck | RIT1 | MUT | 2 | 746 | 0.00268 | 0.00105 | 1 | 282 | 0.00355 | 0.00126 |
| Head and Neck | ROS1 | FUSION | 0 | 746 | 0.00000 | 0.00149 | 1 | 282 | 0.00355 | 0.00358 |
| Head and Neck | ROS1 | MUT | 0 | 746 | 0.00000 | 0.00009 | 0 | 282 | 0.00000 | 0.00019 |
| Head and Neck | SF3B1 | MUT | 4 | 746 | 0.00536 | 0.00807 | 1 | 282 | 0.00355 | 0.00803 |
| Head and Neck | SMO | MUT | 0 | 746 | 0.00000 | 0.00013 | 0 | 282 | 0.00000 | 0.00039 |
| Head and Neck | SPOP | MUT | 2 | 746 | 0.00268 | 0.00355 | 0 | 282 | 0.00000 | 0.00793 |
| Head and Neck | TERT | FUSION | 0 | 746 | 0.00000 | 0.00009 | 0 | 282 | 0.00000 | 0.00000 |
| Head and Neck | TERT | MUT | 122 | 746 | 0.16354 | 0.09795 | 50 | 282 | 0.17730 | 0.12307 |
| Head and Neck | TP53 | HOMDEL | 5 | 746 | 0.00670 | 0.01276 | 2 | 282 | 0.00709 | 0.00900 |
| Head and Neck | TP53 | MUT | 350 | 746 | 0.46917 | 0.52404 | 101 | 282 | 0.35816 | 0.41428 |
| Kidney | AKT1 | MUT | 2 | 510 | 0.00392 | 0.01026 | 1 | 322 | 0.00311 | 0.01045 |
| Kidney | ALK | AMP | 0 | 510 | 0.00000 | 0.00018 | 0 | 322 | 0.00000 | 0.00039 |
| Kidney | ALK | FUSION | 1 | 510 | 0.00196 | 0.00570 | 2 | 322 | 0.00621 | 0.00474 |
| Kidney | ALK | MUT | 0 | 510 | 0.00000 | 0.00061 | 0 | 322 | 0.00000 | 0.00184 |

| | | | | | | | | | | |
|---|---|---|---|---|---|---|---|---|---|---|
| Kidney | AR | AMP | 0 | 510 | 0.00000 | 0.00601 | 0 | 322 | 0.00000 | 0.00842 |
| Kidney | AR | MUT | 0 | 510 | 0.00000 | 0.00136 | 0 | 322 | 0.00000 | 0.00213 |
| Kidney | ARAF | MUT | 0 | 510 | 0.00000 | 0.00039 | 1 | 322 | 0.00311 | 0.00068 |
| Kidney | BRAF | AMP | 2 | 510 | 0.00392 | 0.00250 | 0 | 322 | 0.00000 | 0.00164 |
| Kidney | BRAF | FUSION | 0 | 510 | 0.00000 | 0.00118 | 0 | 322 | 0.00000 | 0.00368 |
| Kidney | BRAF | MUT | 0 | 510 | 0.00000 | 0.04106 | 0 | 322 | 0.00000 | 0.04247 |
| Kidney | CCND1 | AMP | 0 | 510 | 0.00000 | 0.04299 | 0 | 322 | 0.00000 | 0.04189 |
| Kidney | CDK4 | AMP | 0 | 510 | 0.00000 | 0.01921 | 0 | 322 | 0.00000 | 0.02632 |
| Kidney | CDK4 | MUT | 1 | 510 | 0.00196 | 0.00061 | 0 | 322 | 0.00000 | 0.00116 |
| Kidney | CDK6 | AMP | 0 | 510 | 0.00000 | 0.00697 | 0 | 322 | 0.00000 | 0.00590 |
| Kidney | CDKN2A | HOMDEL | 51 | 510 | 0.10000 | 0.12234 | 11 | 322 | 0.03416 | 0.07498 |
| Kidney | CDKN2A | MUT | 7 | 510 | 0.01373 | 0.04917 | 0 | 322 | 0.00000 | 0.03599 |
| Kidney | CTNNB1 | MUT | 0 | 510 | 0.00000 | 0.02601 | 0 | 322 | 0.00000 | 0.02254 |
| Kidney | EGFR | AMP | 1 | 510 | 0.00196 | 0.02996 | 0 | 322 | 0.00000 | 0.03154 |
| Kidney | EGFR | MUT | 0 | 510 | 0.00000 | 0.02649 | 0 | 322 | 0.00000 | 0.04112 |
| Kidney | ERBB2 | AMP | 0 | 510 | 0.00000 | 0.03202 | 0 | 322 | 0.00000 | 0.03938 |
| Kidney | ERBB2 | MUT | 1 | 510 | 0.00196 | 0.01689 | 0 | 322 | 0.00000 | 0.01945 |
| Kidney | ERBB3 | MUT | 0 | 510 | 0.00000 | 0.00561 | 1 | 322 | 0.00311 | 0.00987 |
| Kidney | ERBB4 | MUT | 0 | 510 | 0.00000 | 0.00022 | 0 | 322 | 0.00000 | 0.00155 |
| Kidney | ESR1 | MUT | 0 | 510 | 0.00000 | 0.00965 | 0 | 322 | 0.00000 | 0.01006 |
| Kidney | EZH2 | MUT | 0 | 510 | 0.00000 | 0.00140 | 0 | 322 | 0.00000 | 0.00281 |
| Kidney | FGFR1 | AMP | 3 | 510 | 0.00588 | 0.02816 | 1 | 322 | 0.00311 | 0.02554 |
| Kidney | FGFR1 | FUSION | 0 | 510 | 0.00000 | 0.00013 | 0 | 322 | 0.00000 | 0.00039 |
| Kidney | FGFR2 | AMP | 0 | 510 | 0.00000 | 0.00347 | 0 | 322 | 0.00000 | 0.00358 |
| Kidney | FGFR2 | FUSION | 0 | 510 | 0.00000 | 0.00083 | 0 | 322 | 0.00000 | 0.00300 |
| Kidney | FGFR2 | MUT | 0 | 510 | 0.00000 | 0.00531 | 0 | 322 | 0.00000 | 0.00435 |
| Kidney | FGFR3 | AMP | 0 | 510 | 0.00000 | 0.00263 | 0 | 322 | 0.00000 | 0.00387 |
| Kidney | FGFR3 | FUSION | 0 | 510 | 0.00000 | 0.00294 | 0 | 322 | 0.00000 | 0.00271 |
| Kidney | FGFR3 | MUT | 0 | 510 | 0.00000 | 0.00588 | 0 | 322 | 0.00000 | 0.01200 |
| Kidney | FGFR4 | AMP | 0 | 510 | 0.00000 | 0.00075 | 1 | 322 | 0.00311 | 0.00252 |
| Kidney | GNA11 | MUT | 0 | 510 | 0.00000 | 0.00105 | 0 | 322 | 0.00000 | 0.00184 |
| Kidney | GNAQ | MUT | 0 | 510 | 0.00000 | 0.00096 | 0 | 322 | 0.00000 | 0.00281 |
| Kidney | HRAS | MUT | 0 | 510 | 0.00000 | 0.00504 | 0 | 322 | 0.00000 | 0.00658 |
| Kidney | IDH1 | MUT | 0 | 510 | 0.00000 | 0.01776 | 0 | 322 | 0.00000 | 0.02351 |
| Kidney | IDH2 | MUT | 0 | 510 | 0.00000 | 0.00281 | 0 | 322 | 0.00000 | 0.00290 |
| Kidney | JAK1 | MUT | 0 | 510 | 0.00000 | 0.00026 | 1 | 322 | 0.00311 | 0.00755 |
| Kidney | JAK2 | MUT | 2 | 510 | 0.00392 | 0.00066 | 0 | 322 | 0.00000 | 0.00019 |
| Kidney | KIT | AMP | 6 | 510 | 0.01176 | 0.00675 | 0 | 322 | 0.00000 | 0.00832 |
| Kidney | KIT | MUT | 0 | 510 | 0.00000 | 0.00675 | 0 | 322 | 0.00000 | 0.01335 |
| Kidney | KRAS | AMP | 0 | 510 | 0.00000 | 0.01614 | 0 | 322 | 0.00000 | 0.01809 |
| Kidney | KRAS | MUT | 5 | 510 | 0.00980 | 0.17580 | 2 | 322 | 0.00621 | 0.15257 |
| Kidney | MAP2K1 | MUT | 0 | 510 | 0.00000 | 0.00412 | 1 | 322 | 0.00311 | 0.00629 |
| Kidney | MAP2K2 | MUT | 0 | 510 | 0.00000 | 0.00022 | 0 | 322 | 0.00000 | 0.00058 |
| Kidney | MAP2K4 | MUT | 0 | 510 | 0.00000 | 0.00092 | 0 | 322 | 0.00000 | 0.00687 |
| Kidney | MAPK1 | MUT | 0 | 510 | 0.00000 | 0.00079 | 0 | 322 | 0.00000 | 0.00116 |
| Kidney | MDM2 | AMP | 3 | 510 | 0.00588 | 0.02772 | 1 | 322 | 0.00311 | 0.03696 |
| Kidney | MET | AMP | 3 | 510 | 0.00588 | 0.00693 | 6 | 322 | 0.01863 | 0.00948 |
| Kidney | MET | MUT | 0 | 510 | 0.00000 | 0.00355 | 0 | 322 | 0.00000 | 0.00300 |
| Kidney | MTOR | MUT | 8 | 510 | 0.01569 | 0.00390 | 18 | 322 | 0.05590 | 0.00619 |
| Kidney | MYC | AMP | 2 | 510 | 0.00392 | 0.04066 | 4 | 322 | 0.01242 | 0.03957 |
| Kidney | MYCN | AMP | 0 | 510 | 0.00000 | 0.00325 | 0 | 322 | 0.00000 | 0.00484 |
| Kidney | MYD88 | MUT | 0 | 510 | 0.00000 | 0.00118 | 0 | 322 | 0.00000 | 0.00145 |
| Kidney | NOTCH1 | FUSION | 0 | 510 | 0.00000 | 0.00018 | 0 | 322 | 0.00000 | 0.00048 |
| Kidney | NRAS | MUT | 0 | 510 | 0.00000 | 0.02048 | 0 | 322 | 0.00000 | 0.02070 |
| Kidney | NRG1 | FUSION | 0 | 510 | 0.00000 | 0.00039 | 0 | 322 | 0.00000 | 0.00039 |
| Kidney | NTRK1 | FUSION | 0 | 510 | 0.00000 | 0.00026 | 0 | 322 | 0.00000 | 0.00097 |
| Kidney | NTRK3 | FUSION | 0 | 510 | 0.00000 | 0.00066 | 0 | 322 | 0.00000 | 0.00097 |
| Kidney | PDGFRA | AMP | 8 | 510 | 0.01569 | 0.00627 | 0 | 322 | 0.00000 | 0.00851 |
| Kidney | PDGFRA | MUT | 0 | 510 | 0.00000 | 0.00048 | 0 | 322 | 0.00000 | 0.00223 |
| Kidney | PIK3CA | AMP | 0 | 510 | 0.00000 | 0.02298 | 0 | 322 | 0.00000 | 0.00697 |
| Kidney | PIK3CA | MUT | 21 | 510 | 0.04118 | 0.12387 | 13 | 322 | 0.04037 | 0.11474 |
| Kidney | POLE | MUT | 0 | 510 | 0.00000 | 0.00079 | 0 | 322 | 0.00000 | 0.00252 |
| Kidney | PRKACA | FUSION | 0 | 510 | 0.00000 | 0.00039 | 0 | 322 | 0.00000 | 0.00087 |
| Kidney | PTEN | HOMDEL | 4 | 510 | 0.00784 | 0.03228 | 1 | 322 | 0.00311 | 0.02390 |
| Kidney | PTEN | MUT | 30 | 510 | 0.05882 | 0.06536 | 19 | 322 | 0.05901 | 0.05563 |
| Kidney | RAF1 | MUT | 0 | 510 | 0.00000 | 0.00154 | 0 | 322 | 0.00000 | 0.00261 |
| Kidney | RB1 | HOMDEL | 4 | 510 | 0.00784 | 0.02298 | 0 | 322 | 0.00000 | 0.01896 |
| Kidney | RB1 | MUT | 4 | 510 | 0.00784 | 0.04053 | 2 | 322 | 0.00621 | 0.04092 |
| Kidney | RET | FUSION | 0 | 510 | 0.00000 | 0.00219 | 0 | 322 | 0.00000 | 0.00281 |
| Kidney | RET | MUT | 0 | 510 | 0.00000 | 0.00175 | 0 | 322 | 0.00000 | 0.00213 |
| Kidney | RIT1 | MUT | 0 | 510 | 0.00000 | 0.00105 | 0 | 322 | 0.00000 | 0.00126 |
| Kidney | ROS1 | FUSION | 0 | 510 | 0.00000 | 0.00149 | 0 | 322 | 0.00000 | 0.00358 |
| Kidney | ROS1 | MUT | 0 | 510 | 0.00000 | 0.00009 | 0 | 322 | 0.00000 | 0.00019 |
| Kidney | SF3B1 | MUT | 2 | 510 | 0.00392 | 0.00807 | 2 | 322 | 0.00621 | 0.00803 |
| Kidney | SMO | MUT | 0 | 510 | 0.00000 | 0.00013 | 0 | 322 | 0.00000 | 0.00039 |
| Kidney | SPOP | MUT | 0 | 510 | 0.00000 | 0.00355 | 0 | 322 | 0.00000 | 0.00793 |

| | | | | | | | | | | |
|---|---|---|---|---|---|---|---|---|---|---|
| Kidney | TERT | FUSION | 0 | 510 | 0.00000 | 0.00009 | 0 | 322 | 0.00000 | 0.00000 |
| Kidney | TERT | MUT | 41 | 510 | 0.08039 | 0.09795 | 38 | 322 | 0.11801 | 0.12307 |
| Kidney | TP53 | HOMDEL | 1 | 510 | 0.00196 | 0.01276 | 0 | 322 | 0.00000 | 0.00900 |
| Kidney | TP53 | MUT | 72 | 510 | 0.14118 | 0.52404 | 47 | 322 | 0.14596 | 0.41428 |
| Lung - NSCLC | AKT1 | MUT | 8 | 2784 | 0.00287 | 0.01026 | 4 | 1539 | 0.00260 | 0.01045 |
| Lung - NSCLC | ALK | AMP | 1 | 2784 | 0.00036 | 0.00018 | 0 | 1539 | 0.00000 | 0.00039 |
| Lung - NSCLC | ALK | FUSION | 115 | 2784 | 0.04131 | 0.00570 | 43 | 1539 | 0.02794 | 0.00474 |
| Lung - NSCLC | ALK | MUT | 13 | 2784 | 0.00467 | 0.00061 | 2 | 1539 | 0.00130 | 0.00184 |
| Lung - NSCLC | AR | AMP | 2 | 2784 | 0.00072 | 0.00601 | 1 | 1539 | 0.00065 | 0.00842 |
| Lung - NSCLC | AR | MUT | 0 | 2784 | 0.00000 | 0.00136 | 0 | 1539 | 0.00000 | 0.00213 |
| Lung - NSCLC | ARAF | MUT | 3 | 2784 | 0.00108 | 0.00039 | 2 | 1539 | 0.00130 | 0.00068 |
| Lung - NSCLC | BRAF | AMP | 4 | 2784 | 0.00144 | 0.00250 | 4 | 1539 | 0.00260 | 0.00164 |
| Lung - NSCLC | BRAF | FUSION | 1 | 2784 | 0.00036 | 0.00118 | 6 | 1539 | 0.00390 | 0.00368 |
| Lung - NSCLC | BRAF | MUT | 91 | 2784 | 0.03269 | 0.04106 | 63 | 1539 | 0.04094 | 0.04247 |
| Lung - NSCLC | CCND1 | AMP | 103 | 2784 | 0.03700 | 0.04299 | 40 | 1539 | 0.02599 | 0.04189 |
| Lung - NSCLC | CDK4 | AMP | 79 | 2784 | 0.02838 | 0.01921 | 63 | 1539 | 0.04094 | 0.02632 |
| Lung - NSCLC | CDK4 | MUT | 5 | 2784 | 0.00180 | 0.00061 | 4 | 1539 | 0.00260 | 0.00116 |
| Lung - NSCLC | CDK6 | AMP | 15 | 2784 | 0.00539 | 0.00697 | 5 | 1539 | 0.00325 | 0.00590 |
| Lung - NSCLC | CDKN2A | HOMDEL | 476 | 2784 | 0.17098 | 0.12234 | 123 | 1539 | 0.07992 | 0.07498 |
| Lung - NSCLC | CDKN2A | MUT | 231 | 2784 | 0.08297 | 0.04917 | 92 | 1539 | 0.05978 | 0.03599 |
| Lung - NSCLC | CTNNB1 | MUT | 71 | 2784 | 0.02550 | 0.02601 | 46 | 1539 | 0.02989 | 0.02254 |
| Lung - NSCLC | EGFR | AMP | 134 | 2784 | 0.04813 | 0.02996 | 111 | 1539 | 0.07212 | 0.03154 |
| Lung - NSCLC | EGFR | MUT | 465 | 2784 | 0.16703 | 0.02649 | 336 | 1539 | 0.21832 | 0.04112 |
| Lung - NSCLC | ERBB2 | AMP | 35 | 2784 | 0.01257 | 0.03202 | 38 | 1539 | 0.02469 | 0.03938 |
| Lung - NSCLC | ERBB2 | MUT | 55 | 2784 | 0.01976 | 0.01689 | 41 | 1539 | 0.02664 | 0.01945 |
| Lung - NSCLC | ERBB3 | MUT | 5 | 2784 | 0.00180 | 0.00561 | 4 | 1539 | 0.00260 | 0.00987 |
| Lung - NSCLC | ERBB4 | MUT | 0 | 2784 | 0.00000 | 0.00022 | 0 | 1539 | 0.00000 | 0.00155 |
| Lung - NSCLC | ESR1 | MUT | 0 | 2784 | 0.00000 | 0.00965 | 0 | 1539 | 0.00000 | 0.01006 |
| Lung - NSCLC | EZH2 | MUT | 1 | 2784 | 0.00036 | 0.00140 | 1 | 1539 | 0.00065 | 0.00281 |
| Lung - NSCLC | FGFR1 | AMP | 74 | 2784 | 0.02658 | 0.02816 | 29 | 1539 | 0.01884 | 0.02554 |
| Lung - NSCLC | FGFR1 | FUSION | 1 | 2784 | 0.00036 | 0.00013 | 1 | 1539 | 0.00065 | 0.00039 |
| Lung - NSCLC | FGFR2 | AMP | 1 | 2784 | 0.00036 | 0.00347 | 1 | 1539 | 0.00065 | 0.00358 |
| Lung - NSCLC | FGFR2 | FUSION | 0 | 2784 | 0.00000 | 0.00083 | 1 | 1539 | 0.00065 | 0.00300 |
| Lung - NSCLC | FGFR2 | MUT | 3 | 2784 | 0.00108 | 0.00531 | 2 | 1539 | 0.00130 | 0.00435 |
| Lung - NSCLC | FGFR3 | AMP | 4 | 2784 | 0.00144 | 0.00263 | 5 | 1539 | 0.00325 | 0.00387 |
| Lung - NSCLC | FGFR3 | FUSION | 11 | 2784 | 0.00395 | 0.00294 | 5 | 1539 | 0.00325 | 0.00271 |
| Lung - NSCLC | FGFR3 | MUT | 12 | 2784 | 0.00431 | 0.00588 | 5 | 1539 | 0.00325 | 0.01200 |
| Lung - NSCLC | FGFR4 | AMP | 4 | 2784 | 0.00144 | 0.00075 | 7 | 1539 | 0.00455 | 0.00252 |
| Lung - NSCLC | GNA11 | MUT | 1 | 2784 | 0.00036 | 0.00105 | 0 | 1539 | 0.00000 | 0.00184 |
| Lung - NSCLC | GNAQ | MUT | 1 | 2784 | 0.00036 | 0.00096 | 0 | 1539 | 0.00000 | 0.00281 |
| Lung - NSCLC | HRAS | MUT | 7 | 2784 | 0.00251 | 0.00504 | 2 | 1539 | 0.00130 | 0.00658 |
| Lung - NSCLC | IDH1 | MUT | 12 | 2784 | 0.00431 | 0.01776 | 5 | 1539 | 0.00325 | 0.02351 |
| Lung - NSCLC | IDH2 | MUT | 3 | 2784 | 0.00108 | 0.00281 | 0 | 1539 | 0.00000 | 0.00290 |
| Lung - NSCLC | JAK1 | MUT | 0 | 2784 | 0.00000 | 0.00026 | 6 | 1539 | 0.00390 | 0.00755 |
| Lung - NSCLC | JAK2 | MUT | 1 | 2784 | 0.00036 | 0.00066 | 1 | 1539 | 0.00065 | 0.00019 |
| Lung - NSCLC | KIT | AMP | 19 | 2784 | 0.00682 | 0.00675 | 6 | 1539 | 0.00390 | 0.00832 |
| Lung - NSCLC | KIT | MUT | 6 | 2784 | 0.00216 | 0.00675 | 2 | 1539 | 0.00130 | 0.01335 |
| Lung - NSCLC | KRAS | AMP | 34 | 2784 | 0.01221 | 0.01614 | 25 | 1539 | 0.01624 | 0.01809 |
| Lung - NSCLC | KRAS | MUT | 702 | 2784 | 0.25216 | 0.17580 | 425 | 1539 | 0.27615 | 0.15257 |
| Lung - NSCLC | MAP2K1 | MUT | 10 | 2784 | 0.00359 | 0.00412 | 12 | 1539 | 0.00780 | 0.00629 |
| Lung - NSCLC | MAP2K2 | MUT | 1 | 2784 | 0.00036 | 0.00022 | 0 | 1539 | 0.00000 | 0.00058 |
| Lung - NSCLC | MAP2K4 | MUT | 1 | 2784 | 0.00036 | 0.00092 | 7 | 1539 | 0.00455 | 0.00687 |
| Lung - NSCLC | MAPK1 | MUT | 1 | 2784 | 0.00036 | 0.00079 | 1 | 1539 | 0.00065 | 0.00116 |
| Lung - NSCLC | MDM2 | AMP | 103 | 2784 | 0.03700 | 0.02772 | 85 | 1539 | 0.05523 | 0.03696 |
| Lung - NSCLC | MET | AMP | 50 | 2784 | 0.01796 | 0.00693 | 40 | 1539 | 0.02599 | 0.00948 |
| Lung - NSCLC | MET | MUT | 75 | 2784 | 0.02694 | 0.00355 | 30 | 1539 | 0.01949 | 0.00300 |
| Lung - NSCLC | MTOR | MUT | 8 | 2784 | 0.00287 | 0.00390 | 7 | 1539 | 0.00455 | 0.00619 |
| Lung - NSCLC | MYC | AMP | 117 | 2784 | 0.04203 | 0.04066 | 84 | 1539 | 0.05458 | 0.03957 |
| Lung - NSCLC | MYCN | AMP | 9 | 2784 | 0.00323 | 0.00325 | 1 | 1539 | 0.00065 | 0.00484 |
| Lung - NSCLC | MYD88 | MUT | 1 | 2784 | 0.00036 | 0.00118 | 2 | 1539 | 0.00130 | 0.00145 |
| Lung - NSCLC | NOTCH1 | FUSION | 0 | 2784 | 0.00000 | 0.00018 | 0 | 1539 | 0.00000 | 0.00048 |
| Lung - NSCLC | NRAS | MUT | 26 | 2784 | 0.00934 | 0.02048 | 16 | 1539 | 0.01040 | 0.02070 |
| Lung - NSCLC | NRG1 | FUSION | 7 | 2784 | 0.00251 | 0.00039 | 2 | 1539 | 0.00130 | 0.00039 |
| Lung - NSCLC | NTRK1 | FUSION | 0 | 2784 | 0.00000 | 0.00026 | 2 | 1539 | 0.00130 | 0.00097 |
| Lung - NSCLC | NTRK3 | FUSION | 2 | 2784 | 0.00072 | 0.00066 | 0 | 1539 | 0.00000 | 0.00097 |
| Lung - NSCLC | PDGFRA | AMP | 15 | 2784 | 0.00539 | 0.00627 | 2 | 1539 | 0.00130 | 0.00851 |
| Lung - NSCLC | PDGFRA | MUT | 0 | 2784 | 0.00000 | 0.00048 | 1 | 1539 | 0.00065 | 0.00223 |
| Lung - NSCLC | PIK3CA | AMP | 133 | 2784 | 0.04777 | 0.02298 | 25 | 1539 | 0.01624 | 0.00697 |
| Lung - NSCLC | PIK3CA | MUT | 150 | 2784 | 0.05388 | 0.12387 | 94 | 1539 | 0.06108 | 0.11474 |
| Lung - NSCLC | POLE | MUT | 0 | 2784 | 0.00000 | 0.00079 | 2 | 1539 | 0.00130 | 0.00252 |
| Lung - NSCLC | PRKACA | FUSION | 0 | 2784 | 0.00000 | 0.00039 | 0 | 1539 | 0.00000 | 0.00087 |
| Lung - NSCLC | PTEN | HOMDEL | 32 | 2784 | 0.01149 | 0.03228 | 13 | 1539 | 0.00845 | 0.02390 |
| Lung - NSCLC | PTEN | MUT | 94 | 2784 | 0.03376 | 0.06536 | 41 | 1539 | 0.02664 | 0.05563 |
| Lung - NSCLC | RAF1 | MUT | 4 | 2784 | 0.00144 | 0.00154 | 5 | 1539 | 0.00325 | 0.00261 |
| Lung - NSCLC | RB1 | HOMDEL | 56 | 2784 | 0.02011 | 0.02298 | 19 | 1539 | 0.01235 | 0.01896 |
| Lung - NSCLC | RB1 | MUT | 150 | 2784 | 0.05388 | 0.04053 | 72 | 1539 | 0.04678 | 0.04092 |

| | | | | | | | | | | |
|---|---|---|---|---|---|---|---|---|---|---|
| Lung - NSCLC | RET | FUSION | 32 | 2784 | 0.01149 | 0.00219 | 18 | 1539 | 0.01170 | 0.00281 |
| Lung - NSCLC | RET | MUT | 1 | 2784 | 0.00036 | 0.00175 | 3 | 1539 | 0.00195 | 0.00213 |
| Lung - NSCLC | RIT1 | MUT | 10 | 2784 | 0.00359 | 0.00105 | 1 | 1539 | 0.00065 | 0.00126 |
| Lung - NSCLC | ROS1 | FUSION | 26 | 2784 | 0.00934 | 0.00149 | 27 | 1539 | 0.01754 | 0.00358 |
| Lung - NSCLC | ROS1 | MUT | 1 | 2784 | 0.00036 | 0.00009 | 1 | 1539 | 0.00065 | 0.00019 |
| Lung - NSCLC | SF3B1 | MUT | 18 | 2784 | 0.00647 | 0.00807 | 7 | 1539 | 0.00455 | 0.00803 |
| Lung - NSCLC | SMO | MUT | 0 | 2784 | 0.00000 | 0.00013 | 0 | 1539 | 0.00000 | 0.00039 |
| Lung - NSCLC | SPOP | MUT | 2 | 2784 | 0.00072 | 0.00355 | 1 | 1539 | 0.00065 | 0.00793 |
| Lung - NSCLC | TERT | FUSION | 0 | 2784 | 0.00000 | 0.00009 | 0 | 1539 | 0.00000 | 0.00000 |
| Lung - NSCLC | TERT | MUT | 73 | 2784 | 0.02622 | 0.09795 | 30 | 1539 | 0.01949 | 0.12307 |
| Lung - NSCLC | TP53 | HOMDEL | 24 | 2784 | 0.00862 | 0.01276 | 5 | 1539 | 0.00325 | 0.00900 |
| Lung - NSCLC | TP53 | MUT | 1709 | 2784 | 0.61386 | 0.52404 | 848 | 1539 | 0.55101 | 0.41428 |
| Lung - SCLC | AKT1 | MUT | 3 | 433 | 0.00693 | 0.01026 | 0 | 81 | 0.00000 | 0.01045 |
| Lung - SCLC | ALK | AMP | 0 | 433 | 0.00000 | 0.00018 | 0 | 81 | 0.00000 | 0.00039 |
| Lung - SCLC | ALK | FUSION | 2 | 433 | 0.00462 | 0.00570 | 0 | 81 | 0.00000 | 0.00474 |
| Lung - SCLC | ALK | MUT | 0 | 433 | 0.00000 | 0.00061 | 1 | 81 | 0.01235 | 0.00184 |
| Lung - SCLC | AR | AMP | 2 | 433 | 0.00462 | 0.00601 | 0 | 81 | 0.00000 | 0.00842 |
| Lung - SCLC | AR | MUT | 0 | 433 | 0.00000 | 0.00136 | 0 | 81 | 0.00000 | 0.00213 |
| Lung - SCLC | ARAF | MUT | 0 | 433 | 0.00000 | 0.00039 | 0 | 81 | 0.00000 | 0.00068 |
| Lung - SCLC | BRAF | AMP | 2 | 433 | 0.00462 | 0.00250 | 0 | 81 | 0.00000 | 0.00164 |
| Lung - SCLC | BRAF | FUSION | 0 | 433 | 0.00000 | 0.00118 | 0 | 81 | 0.00000 | 0.00368 |
| Lung - SCLC | BRAF | MUT | 1 | 433 | 0.00231 | 0.04106 | 1 | 81 | 0.01235 | 0.04247 |
| Lung - SCLC | CCND1 | AMP | 5 | 433 | 0.01155 | 0.04299 | 2 | 81 | 0.02469 | 0.04189 |
| Lung - SCLC | CDK4 | AMP | 2 | 433 | 0.00462 | 0.01921 | 0 | 81 | 0.00000 | 0.02632 |
| Lung - SCLC | CDK4 | MUT | 0 | 433 | 0.00000 | 0.00061 | 1 | 81 | 0.01235 | 0.00116 |
| Lung - SCLC | CDK6 | AMP | 0 | 433 | 0.00000 | 0.00697 | 0 | 81 | 0.00000 | 0.00590 |
| Lung - SCLC | CDKN2A | HOMDEL | 26 | 433 | 0.06005 | 0.12234 | 3 | 81 | 0.03704 | 0.07498 |
| Lung - SCLC | CDKN2A | MUT | 9 | 433 | 0.02079 | 0.04917 | 0 | 81 | 0.00000 | 0.03599 |
| Lung - SCLC | CTNNB1 | MUT | 2 | 433 | 0.00462 | 0.02601 | 0 | 81 | 0.00000 | 0.02254 |
| Lung - SCLC | EGFR | AMP | 1 | 433 | 0.00231 | 0.02996 | 0 | 81 | 0.00000 | 0.03154 |
| Lung - SCLC | EGFR | MUT | 8 | 433 | 0.01848 | 0.02649 | 1 | 81 | 0.01235 | 0.04112 |
| Lung - SCLC | ERBB2 | AMP | 3 | 433 | 0.00693 | 0.03202 | 0 | 81 | 0.00000 | 0.03938 |
| Lung - SCLC | ERBB2 | MUT | 0 | 433 | 0.00000 | 0.01689 | 0 | 81 | 0.00000 | 0.01945 |
| Lung - SCLC | ERBB3 | MUT | 0 | 433 | 0.00000 | 0.00561 | 0 | 81 | 0.00000 | 0.00987 |
| Lung - SCLC | ERBB4 | MUT | 0 | 433 | 0.00000 | 0.00022 | 0 | 81 | 0.00000 | 0.00155 |
| Lung - SCLC | ESR1 | MUT | 0 | 433 | 0.00000 | 0.00965 | 0 | 81 | 0.00000 | 0.01006 |
| Lung - SCLC | EZH2 | MUT | 0 | 433 | 0.00000 | 0.00140 | 0 | 81 | 0.00000 | 0.00281 |
| Lung - SCLC | FGFR1 | AMP | 12 | 433 | 0.02771 | 0.02816 | 2 | 81 | 0.02469 | 0.02554 |
| Lung - SCLC | FGFR1 | FUSION | 0 | 433 | 0.00000 | 0.00013 | 0 | 81 | 0.00000 | 0.00039 |
| Lung - SCLC | FGFR2 | AMP | 0 | 433 | 0.00000 | 0.00347 | 0 | 81 | 0.00000 | 0.00358 |
| Lung - SCLC | FGFR2 | FUSION | 0 | 433 | 0.00000 | 0.00083 | 0 | 81 | 0.00000 | 0.00300 |
| Lung - SCLC | FGFR2 | MUT | 2 | 433 | 0.00462 | 0.00531 | 0 | 81 | 0.00000 | 0.00435 |
| Lung - SCLC | FGFR3 | AMP | 0 | 433 | 0.00000 | 0.00263 | 1 | 81 | 0.01235 | 0.00387 |
| Lung - SCLC | FGFR3 | FUSION | 0 | 433 | 0.00000 | 0.00294 | 0 | 81 | 0.00000 | 0.00271 |
| Lung - SCLC | FGFR3 | MUT | 1 | 433 | 0.00231 | 0.00588 | 0 | 81 | 0.00000 | 0.01200 |
| Lung - SCLC | FGFR4 | AMP | 0 | 433 | 0.00000 | 0.00075 | 0 | 81 | 0.00000 | 0.00252 |
| Lung - SCLC | GNA11 | MUT | 0 | 433 | 0.00000 | 0.00105 | 0 | 81 | 0.00000 | 0.00184 |
| Lung - SCLC | GNAQ | MUT | 0 | 433 | 0.00000 | 0.00096 | 0 | 81 | 0.00000 | 0.00281 |
| Lung - SCLC | HRAS | MUT | 2 | 433 | 0.00462 | 0.00504 | 0 | 81 | 0.00000 | 0.00658 |
| Lung - SCLC | IDH1 | MUT | 0 | 433 | 0.00000 | 0.01776 | 0 | 81 | 0.00000 | 0.02351 |
| Lung - SCLC | IDH2 | MUT | 0 | 433 | 0.00000 | 0.00281 | 0 | 81 | 0.00000 | 0.00290 |
| Lung - SCLC | JAK1 | MUT | 0 | 433 | 0.00000 | 0.00026 | 0 | 81 | 0.00000 | 0.00755 |
| Lung - SCLC | JAK2 | MUT | 0 | 433 | 0.00000 | 0.00066 | 0 | 81 | 0.00000 | 0.00019 |
| Lung - SCLC | KIT | AMP | 7 | 433 | 0.01617 | 0.00675 | 2 | 81 | 0.02469 | 0.00832 |
| Lung - SCLC | KIT | MUT | 1 | 433 | 0.00231 | 0.00675 | 0 | 81 | 0.00000 | 0.01335 |
| Lung - SCLC | KRAS | AMP | 6 | 433 | 0.01386 | 0.01614 | 0 | 81 | 0.00000 | 0.01809 |
| Lung - SCLC | KRAS | MUT | 28 | 433 | 0.06467 | 0.17580 | 0 | 81 | 0.00000 | 0.15257 |
| Lung - SCLC | MAP2K1 | MUT | 0 | 433 | 0.00000 | 0.00412 | 0 | 81 | 0.00000 | 0.00629 |
| Lung - SCLC | MAP2K2 | MUT | 0 | 433 | 0.00000 | 0.00022 | 0 | 81 | 0.00000 | 0.00058 |
| Lung - SCLC | MAP2K4 | MUT | 0 | 433 | 0.00000 | 0.00092 | 1 | 81 | 0.01235 | 0.00687 |
| Lung - SCLC | MAPK1 | MUT | 0 | 433 | 0.00000 | 0.00079 | 0 | 81 | 0.00000 | 0.00116 |
| Lung - SCLC | MDM2 | AMP | 5 | 433 | 0.01155 | 0.02772 | 0 | 81 | 0.00000 | 0.03696 |
| Lung - SCLC | MET | AMP | 1 | 433 | 0.00231 | 0.00693 | 0 | 81 | 0.00000 | 0.00948 |
| Lung - SCLC | MET | MUT | 1 | 433 | 0.00231 | 0.00355 | 0 | 81 | 0.00000 | 0.00300 |
| Lung - SCLC | MTOR | MUT | 0 | 433 | 0.00000 | 0.00390 | 1 | 81 | 0.01235 | 0.00619 |
| Lung - SCLC | MYC | AMP | 24 | 433 | 0.05543 | 0.04066 | 1 | 81 | 0.01235 | 0.03957 |
| Lung - SCLC | MYCN | AMP | 14 | 433 | 0.03233 | 0.00325 | 0 | 81 | 0.00000 | 0.00484 |
| Lung - SCLC | MYD88 | MUT | 1 | 433 | 0.00231 | 0.00118 | 0 | 81 | 0.00000 | 0.00145 |
| Lung - SCLC | NOTCH1 | FUSION | 0 | 433 | 0.00000 | 0.00018 | 0 | 81 | 0.00000 | 0.00048 |
| Lung - SCLC | NRAS | MUT | 1 | 433 | 0.00231 | 0.02048 | 0 | 81 | 0.00000 | 0.02070 |
| Lung - SCLC | NRG1 | FUSION | 0 | 433 | 0.00000 | 0.00039 | 0 | 81 | 0.00000 | 0.00039 |
| Lung - SCLC | NTRK1 | FUSION | 0 | 433 | 0.00000 | 0.00026 | 0 | 81 | 0.00000 | 0.00097 |
| Lung - SCLC | NTRK3 | FUSION | 0 | 433 | 0.00000 | 0.00066 | 0 | 81 | 0.00000 | 0.00097 |
| Lung - SCLC | PDGFRA | AMP | 6 | 433 | 0.01386 | 0.00627 | 2 | 81 | 0.02469 | 0.00851 |
| Lung - SCLC | PDGFRA | MUT | 0 | 433 | 0.00000 | 0.00048 | 0 | 81 | 0.00000 | 0.00223 |
| Lung - SCLC | PIK3CA | AMP | 17 | 433 | 0.03926 | 0.02298 | 3 | 81 | 0.03704 | 0.00697 |

| | | | | | | | | | | |
|---|---|---|---|---|---|---|---|---|---|---|
| Lung - SCLC | PIK3CA | MUT | 19 | 433 | 0.04388 | 0.12387 | 2 | 81 | 0.02469 | 0.11474 |
| Lung - SCLC | POLE | MUT | 0 | 433 | 0.00000 | 0.00079 | 1 | 81 | 0.01235 | 0.00252 |
| Lung - SCLC | PRKACA | FUSION | 0 | 433 | 0.00000 | 0.00039 | 0 | 81 | 0.00000 | 0.00087 |
| Lung - SCLC | PTEN | HOMDEL | 12 | 433 | 0.02771 | 0.03228 | 5 | 81 | 0.06173 | 0.02390 |
| Lung - SCLC | PTEN | MUT | 26 | 433 | 0.06005 | 0.06536 | 5 | 81 | 0.06173 | 0.05563 |
| Lung - SCLC | RAF1 | MUT | 0 | 433 | 0.00000 | 0.00154 | 0 | 81 | 0.00000 | 0.00261 |
| Lung - SCLC | RB1 | HOMDEL | 48 | 433 | 0.11085 | 0.02298 | 14 | 81 | 0.17284 | 0.01896 |
| Lung - SCLC | RB1 | MUT | 205 | 433 | 0.47344 | 0.04053 | 48 | 81 | 0.59259 | 0.04092 |
| Lung - SCLC | RET | FUSION | 1 | 433 | 0.00231 | 0.00219 | 0 | 81 | 0.00000 | 0.00281 |
| Lung - SCLC | RET | MUT | 0 | 433 | 0.00000 | 0.00175 | 0 | 81 | 0.00000 | 0.00213 |
| Lung - SCLC | RIT1 | MUT | 0 | 433 | 0.00000 | 0.00105 | 1 | 81 | 0.01235 | 0.00126 |
| Lung - SCLC | ROS1 | FUSION | 0 | 433 | 0.00000 | 0.00149 | 0 | 81 | 0.00000 | 0.00358 |
| Lung - SCLC | ROS1 | MUT | 0 | 433 | 0.00000 | 0.00009 | 0 | 81 | 0.00000 | 0.00019 |
| Lung - SCLC | SF3B1 | MUT | 0 | 433 | 0.00000 | 0.00807 | 0 | 81 | 0.00000 | 0.00803 |
| Lung - SCLC | SMO | MUT | 0 | 433 | 0.00000 | 0.00013 | 0 | 81 | 0.00000 | 0.00039 |
| Lung - SCLC | SPOP | MUT | 0 | 433 | 0.00000 | 0.00355 | 0 | 81 | 0.00000 | 0.00793 |
| Lung - SCLC | TERT | FUSION | 0 | 433 | 0.00000 | 0.00009 | 0 | 81 | 0.00000 | 0.00000 |
| Lung - SCLC | TERT | MUT | 5 | 433 | 0.01155 | 0.09795 | 1 | 81 | 0.01235 | 0.12307 |
| Lung - SCLC | TP53 | HOMDEL | 8 | 433 | 0.01848 | 0.01276 | 4 | 81 | 0.04938 | 0.00900 |
| Lung - SCLC | TP53 | MUT | 341 | 433 | 0.78753 | 0.52404 | 70 | 81 | 0.86420 | 0.41428 |
| Lymphoma | AKT1 | MUT | 0 | 182 | 0.00000 | 0.01026 | 1 | 171 | 0.00585 | 0.01045 |
| Lymphoma | ALK | AMP | 0 | 182 | 0.00000 | 0.00018 | 0 | 171 | 0.00000 | 0.00039 |
| Lymphoma | ALK | FUSION | 0 | 182 | 0.00000 | 0.00570 | 0 | 171 | 0.00000 | 0.00474 |
| Lymphoma | ALK | MUT | 0 | 182 | 0.00000 | 0.00061 | 0 | 171 | 0.00000 | 0.00184 |
| Lymphoma | AR | AMP | 0 | 182 | 0.00000 | 0.00601 | 0 | 171 | 0.00000 | 0.00842 |
| Lymphoma | AR | MUT | 0 | 182 | 0.00000 | 0.00136 | 0 | 171 | 0.00000 | 0.00213 |
| Lymphoma | ARAF | MUT | 0 | 182 | 0.00000 | 0.00039 | 0 | 171 | 0.00000 | 0.00068 |
| Lymphoma | BRAF | AMP | 0 | 182 | 0.00000 | 0.00250 | 0 | 171 | 0.00000 | 0.00164 |
| Lymphoma | BRAF | FUSION | 0 | 182 | 0.00000 | 0.00118 | 0 | 171 | 0.00000 | 0.00368 |
| Lymphoma | BRAF | MUT | 8 | 182 | 0.04396 | 0.04106 | 7 | 171 | 0.04094 | 0.04247 |
| Lymphoma | CCND1 | AMP | 0 | 182 | 0.00000 | 0.04299 | 1 | 171 | 0.00585 | 0.04189 |
| Lymphoma | CDK4 | AMP | 3 | 182 | 0.01648 | 0.01921 | 0 | 171 | 0.00000 | 0.02632 |
| Lymphoma | CDK4 | MUT | 0 | 182 | 0.00000 | 0.00061 | 0 | 171 | 0.00000 | 0.00116 |
| Lymphoma | CDK6 | AMP | 2 | 182 | 0.01099 | 0.00697 | 2 | 171 | 0.01170 | 0.00590 |
| Lymphoma | CDKN2A | HOMDEL | 25 | 182 | 0.13736 | 0.12234 | 20 | 171 | 0.11696 | 0.07498 |
| Lymphoma | CDKN2A | MUT | 2 | 182 | 0.01099 | 0.04917 | 3 | 171 | 0.01754 | 0.03599 |
| Lymphoma | CTNNB1 | MUT | 0 | 182 | 0.00000 | 0.02601 | 0 | 171 | 0.00000 | 0.02254 |
| Lymphoma | EGFR | AMP | 0 | 182 | 0.00000 | 0.02996 | 0 | 171 | 0.00000 | 0.03154 |
| Lymphoma | EGFR | MUT | 0 | 182 | 0.00000 | 0.02649 | 1 | 171 | 0.00585 | 0.04112 |
| Lymphoma | ERBB2 | AMP | 0 | 182 | 0.00000 | 0.03202 | 0 | 171 | 0.00000 | 0.03938 |
| Lymphoma | ERBB2 | MUT | 0 | 182 | 0.00000 | 0.01689 | 0 | 171 | 0.00000 | 0.01945 |
| Lymphoma | ERBB3 | MUT | 0 | 182 | 0.00000 | 0.00561 | 0 | 171 | 0.00000 | 0.00987 |
| Lymphoma | ERBB4 | MUT | 0 | 182 | 0.00000 | 0.00022 | 0 | 171 | 0.00000 | 0.00155 |
| Lymphoma | ESR1 | MUT | 0 | 182 | 0.00000 | 0.00965 | 0 | 171 | 0.00000 | 0.01006 |
| Lymphoma | EZH2 | MUT | 19 | 182 | 0.10440 | 0.00140 | 9 | 171 | 0.05263 | 0.00281 |
| Lymphoma | FGFR1 | AMP | 0 | 182 | 0.00000 | 0.02816 | 0 | 171 | 0.00000 | 0.02554 |
| Lymphoma | FGFR1 | FUSION | 0 | 182 | 0.00000 | 0.00013 | 0 | 171 | 0.00000 | 0.00039 |
| Lymphoma | FGFR2 | AMP | 0 | 182 | 0.00000 | 0.00347 | 0 | 171 | 0.00000 | 0.00358 |
| Lymphoma | FGFR2 | FUSION | 0 | 182 | 0.00000 | 0.00083 | 0 | 171 | 0.00000 | 0.00300 |
| Lymphoma | FGFR2 | MUT | 0 | 182 | 0.00000 | 0.00531 | 0 | 171 | 0.00000 | 0.00435 |
| Lymphoma | FGFR3 | AMP | 0 | 182 | 0.00000 | 0.00263 | 0 | 171 | 0.00000 | 0.00387 |
| Lymphoma | FGFR3 | FUSION | 0 | 182 | 0.00000 | 0.00294 | 0 | 171 | 0.00000 | 0.00271 |
| Lymphoma | FGFR3 | MUT | 0 | 182 | 0.00000 | 0.00588 | 0 | 171 | 0.00000 | 0.01200 |
| Lymphoma | FGFR4 | AMP | 0 | 182 | 0.00000 | 0.00075 | 0 | 171 | 0.00000 | 0.00252 |
| Lymphoma | GNA11 | MUT | 0 | 182 | 0.00000 | 0.00105 | 0 | 171 | 0.00000 | 0.00184 |
| Lymphoma | GNAQ | MUT | 0 | 182 | 0.00000 | 0.00096 | 0 | 171 | 0.00000 | 0.00281 |
| Lymphoma | HRAS | MUT | 0 | 182 | 0.00000 | 0.00504 | 0 | 171 | 0.00000 | 0.00658 |
| Lymphoma | IDH1 | MUT | 1 | 182 | 0.00549 | 0.01776 | 0 | 171 | 0.00000 | 0.02351 |
| Lymphoma | IDH2 | MUT | 1 | 182 | 0.00549 | 0.00281 | 7 | 171 | 0.04094 | 0.00290 |
| Lymphoma | JAK1 | MUT | 0 | 182 | 0.00000 | 0.00026 | 0 | 171 | 0.00000 | 0.00755 |
| Lymphoma | JAK2 | MUT | 0 | 182 | 0.00000 | 0.00066 | 0 | 171 | 0.00000 | 0.00019 |
| Lymphoma | KIT | AMP | 0 | 182 | 0.00000 | 0.00675 | 0 | 171 | 0.00000 | 0.00832 |
| Lymphoma | KIT | MUT | 0 | 182 | 0.00000 | 0.00675 | 0 | 171 | 0.00000 | 0.01335 |
| Lymphoma | KRAS | AMP | 0 | 182 | 0.00000 | 0.01614 | 0 | 171 | 0.00000 | 0.01809 |
| Lymphoma | KRAS | MUT | 6 | 182 | 0.03297 | 0.17580 | 3 | 171 | 0.01754 | 0.15257 |
| Lymphoma | MAP2K1 | MUT | 4 | 182 | 0.02198 | 0.00412 | 3 | 171 | 0.01754 | 0.00629 |
| Lymphoma | MAP2K2 | MUT | 0 | 182 | 0.00000 | 0.00022 | 0 | 171 | 0.00000 | 0.00058 |
| Lymphoma | MAP2K4 | MUT | 0 | 182 | 0.00000 | 0.00092 | 0 | 171 | 0.00000 | 0.00687 |
| Lymphoma | MAPK1 | MUT | 0 | 182 | 0.00000 | 0.00079 | 1 | 171 | 0.00585 | 0.00116 |
| Lymphoma | MDM2 | AMP | 1 | 182 | 0.00549 | 0.02772 | 1 | 171 | 0.00585 | 0.03696 |
| Lymphoma | MET | AMP | 0 | 182 | 0.00000 | 0.00693 | 1 | 171 | 0.00585 | 0.00948 |
| Lymphoma | MET | MUT | 0 | 182 | 0.00000 | 0.00355 | 0 | 171 | 0.00000 | 0.00300 |
| Lymphoma | MTOR | MUT | 1 | 182 | 0.00549 | 0.00390 | 2 | 171 | 0.01170 | 0.00619 |
| Lymphoma | MYC | AMP | 0 | 182 | 0.00000 | 0.04066 | 3 | 171 | 0.01754 | 0.03957 |
| Lymphoma | MYCN | AMP | 0 | 182 | 0.00000 | 0.00325 | 0 | 171 | 0.00000 | 0.00484 |
| Lymphoma | MYD88 | MUT | 18 | 182 | 0.09890 | 0.00118 | 10 | 171 | 0.05848 | 0.00145 |

| | | | | | | | | | | |
|---|---|---|---|---|---|---|---|---|---|---|
| Lymphoma | NOTCH1 | FUSION | 0 | 182 | 0.00000 | 0.00018 | 0 | 171 | 0.00000 | 0.00048 |
| Lymphoma | NRAS | MUT | 4 | 182 | 0.02198 | 0.02048 | 1 | 171 | 0.00585 | 0.02070 |
| Lymphoma | NRG1 | FUSION | 0 | 182 | 0.00000 | 0.00039 | 0 | 171 | 0.00000 | 0.00039 |
| Lymphoma | NTRK1 | FUSION | 0 | 182 | 0.00000 | 0.00026 | 0 | 171 | 0.00000 | 0.00097 |
| Lymphoma | NTRK3 | FUSION | 0 | 182 | 0.00000 | 0.00066 | 0 | 171 | 0.00000 | 0.00097 |
| Lymphoma | PDGFRA | AMP | 0 | 182 | 0.00000 | 0.00627 | 0 | 171 | 0.00000 | 0.00851 |
| Lymphoma | PDGFRA | MUT | 0 | 182 | 0.00000 | 0.00048 | 0 | 171 | 0.00000 | 0.00223 |
| Lymphoma | PIK3CA | AMP | 1 | 182 | 0.00549 | 0.02298 | 0 | 171 | 0.00000 | 0.00697 |
| Lymphoma | PIK3CA | MUT | 1 | 182 | 0.00549 | 0.12387 | 1 | 171 | 0.00585 | 0.11474 |
| Lymphoma | POLE | MUT | 0 | 182 | 0.00000 | 0.00079 | 0 | 171 | 0.00000 | 0.00252 |
| Lymphoma | PRKACA | FUSION | 0 | 182 | 0.00000 | 0.00039 | 0 | 171 | 0.00000 | 0.00087 |
| Lymphoma | PTEN | HOMDEL | 3 | 182 | 0.01648 | 0.03228 | 2 | 171 | 0.01170 | 0.02390 |
| Lymphoma | PTEN | MUT | 4 | 182 | 0.02198 | 0.06536 | 4 | 171 | 0.02339 | 0.05563 |
| Lymphoma | RAF1 | MUT | 1 | 182 | 0.00549 | 0.00154 | 0 | 171 | 0.00000 | 0.00261 |
| Lymphoma | RB1 | HOMDEL | 3 | 182 | 0.01648 | 0.02298 | 2 | 171 | 0.01170 | 0.01896 |
| Lymphoma | RB1 | MUT | 2 | 182 | 0.01099 | 0.04053 | 1 | 171 | 0.00585 | 0.04092 |
| Lymphoma | RET | FUSION | 0 | 182 | 0.00000 | 0.00219 | 0 | 171 | 0.00000 | 0.00281 |
| Lymphoma | RET | MUT | 0 | 182 | 0.00000 | 0.00175 | 0 | 171 | 0.00000 | 0.00213 |
| Lymphoma | RIT1 | MUT | 0 | 182 | 0.00000 | 0.00105 | 0 | 171 | 0.00000 | 0.00126 |
| Lymphoma | ROS1 | FUSION | 0 | 182 | 0.00000 | 0.00149 | 0 | 171 | 0.00000 | 0.00358 |
| Lymphoma | ROS1 | MUT | 0 | 182 | 0.00000 | 0.00009 | 0 | 171 | 0.00000 | 0.00019 |
| Lymphoma | SF3B1 | MUT | 3 | 182 | 0.01648 | 0.00807 | 1 | 171 | 0.00585 | 0.00803 |
| Lymphoma | SMO | MUT | 0 | 182 | 0.00000 | 0.00013 | 0 | 171 | 0.00000 | 0.00039 |
| Lymphoma | SPOP | MUT | 0 | 182 | 0.00000 | 0.00355 | 0 | 171 | 0.00000 | 0.00793 |
| Lymphoma | TERT | FUSION | 0 | 182 | 0.00000 | 0.00009 | 0 | 171 | 0.00000 | 0.00000 |
| Lymphoma | TERT | MUT | 2 | 182 | 0.01099 | 0.09795 | 1 | 171 | 0.00585 | 0.12307 |
| Lymphoma | TP53 | HOMDEL | 2 | 182 | 0.01099 | 0.01276 | 3 | 171 | 0.01754 | 0.00900 |
| Lymphoma | TP53 | MUT | 33 | 182 | 0.18132 | 0.52404 | 35 | 171 | 0.20468 | 0.41428 |
| Melanoma | AKT1 | MUT | 0 | 697 | 0.00000 | 0.01026 | 0 | 349 | 0.00000 | 0.01045 |
| Melanoma | ALK | AMP | 0 | 697 | 0.00000 | 0.00018 | 0 | 349 | 0.00000 | 0.00039 |
| Melanoma | ALK | FUSION | 0 | 697 | 0.00000 | 0.00570 | 0 | 349 | 0.00000 | 0.00474 |
| Melanoma | ALK | MUT | 0 | 697 | 0.00000 | 0.00061 | 1 | 349 | 0.00287 | 0.00184 |
| Melanoma | AR | AMP | 0 | 697 | 0.00000 | 0.00601 | 0 | 349 | 0.00000 | 0.00842 |
| Melanoma | AR | MUT | 0 | 697 | 0.00000 | 0.00136 | 0 | 349 | 0.00000 | 0.00213 |
| Melanoma | ARAF | MUT | 3 | 697 | 0.00430 | 0.00039 | 0 | 349 | 0.00000 | 0.00068 |
| Melanoma | BRAF | AMP | 13 | 697 | 0.01865 | 0.00250 | 4 | 349 | 0.01146 | 0.00164 |
| Melanoma | BRAF | FUSION | 3 | 697 | 0.00430 | 0.00118 | 6 | 349 | 0.01719 | 0.00368 |
| Melanoma | BRAF | MUT | 263 | 697 | 0.37733 | 0.04106 | 102 | 349 | 0.29226 | 0.04247 |
| Melanoma | CCND1 | AMP | 16 | 697 | 0.02296 | 0.04299 | 17 | 349 | 0.04871 | 0.04189 |
| Melanoma | CDK4 | AMP | 24 | 697 | 0.03443 | 0.01921 | 11 | 349 | 0.03152 | 0.02632 |
| Melanoma | CDK4 | MUT | 5 | 697 | 0.00717 | 0.00061 | 5 | 349 | 0.01433 | 0.00116 |
| Melanoma | CDK6 | AMP | 2 | 697 | 0.00287 | 0.00697 | 3 | 349 | 0.00860 | 0.00590 |
| Melanoma | CDKN2A | HOMDEL | 165 | 697 | 0.23673 | 0.12234 | 61 | 349 | 0.17479 | 0.07498 |
| Melanoma | CDKN2A | MUT | 97 | 697 | 0.13917 | 0.04917 | 46 | 349 | 0.13181 | 0.03599 |
| Melanoma | CTNNB1 | MUT | 33 | 697 | 0.04735 | 0.02601 | 16 | 349 | 0.04585 | 0.02254 |
| Melanoma | EGFR | AMP | 0 | 697 | 0.00000 | 0.02996 | 2 | 349 | 0.00573 | 0.03154 |
| Melanoma | EGFR | MUT | 0 | 697 | 0.00000 | 0.02649 | 2 | 349 | 0.00573 | 0.04112 |
| Melanoma | ERBB2 | AMP | 1 | 697 | 0.00143 | 0.03202 | 4 | 349 | 0.01146 | 0.03938 |
| Melanoma | ERBB2 | MUT | 2 | 697 | 0.00287 | 0.01689 | 2 | 349 | 0.00573 | 0.01945 |
| Melanoma | ERBB3 | MUT | 0 | 697 | 0.00000 | 0.00561 | 0 | 349 | 0.00000 | 0.00987 |
| Melanoma | ERBB4 | MUT | 2 | 697 | 0.00287 | 0.00022 | 4 | 349 | 0.01146 | 0.00155 |
| Melanoma | ESR1 | MUT | 0 | 697 | 0.00000 | 0.00965 | 0 | 349 | 0.00000 | 0.01006 |
| Melanoma | EZH2 | MUT | 8 | 697 | 0.01148 | 0.00140 | 5 | 349 | 0.01433 | 0.00281 |
| Melanoma | FGFR1 | AMP | 1 | 697 | 0.00143 | 0.02816 | 1 | 349 | 0.00287 | 0.02554 |
| Melanoma | FGFR1 | FUSION | 0 | 697 | 0.00000 | 0.00013 | 0 | 349 | 0.00000 | 0.00039 |
| Melanoma | FGFR2 | AMP | 0 | 697 | 0.00000 | 0.00347 | 1 | 349 | 0.00287 | 0.00358 |
| Melanoma | FGFR2 | FUSION | 0 | 697 | 0.00000 | 0.00083 | 0 | 349 | 0.00000 | 0.00300 |
| Melanoma | FGFR2 | MUT | 0 | 697 | 0.00000 | 0.00531 | 3 | 349 | 0.00860 | 0.00435 |
| Melanoma | FGFR3 | AMP | 1 | 697 | 0.00143 | 0.00263 | 0 | 349 | 0.00000 | 0.00387 |
| Melanoma | FGFR3 | FUSION | 0 | 697 | 0.00000 | 0.00294 | 0 | 349 | 0.00000 | 0.00271 |
| Melanoma | FGFR3 | MUT | 0 | 697 | 0.00000 | 0.00588 | 2 | 349 | 0.00573 | 0.01200 |
| Melanoma | FGFR4 | AMP | 0 | 697 | 0.00000 | 0.00075 | 0 | 349 | 0.00000 | 0.00252 |
| Melanoma | GNA11 | MUT | 21 | 697 | 0.03013 | 0.00105 | 19 | 349 | 0.05444 | 0.00184 |
| Melanoma | GNAQ | MUT | 21 | 697 | 0.03013 | 0.00096 | 28 | 349 | 0.08023 | 0.00281 |
| Melanoma | HRAS | MUT | 6 | 697 | 0.00861 | 0.00504 | 9 | 349 | 0.02579 | 0.00658 |
| Melanoma | IDH1 | MUT | 14 | 697 | 0.02009 | 0.01776 | 8 | 349 | 0.02292 | 0.02351 |
| Melanoma | IDH2 | MUT | 0 | 697 | 0.00000 | 0.00281 | 0 | 349 | 0.00000 | 0.00290 |
| Melanoma | JAK1 | MUT | 1 | 697 | 0.00143 | 0.00026 | 2 | 349 | 0.00573 | 0.00755 |
| Melanoma | JAK2 | MUT | 1 | 697 | 0.00143 | 0.00066 | 0 | 349 | 0.00000 | 0.00019 |
| Melanoma | KIT | AMP | 12 | 697 | 0.01722 | 0.00675 | 11 | 349 | 0.03152 | 0.00832 |
| Melanoma | KIT | MUT | 28 | 697 | 0.04017 | 0.00675 | 18 | 349 | 0.05158 | 0.01335 |
| Melanoma | KRAS | AMP | 5 | 697 | 0.00717 | 0.01614 | 4 | 349 | 0.01146 | 0.01809 |
| Melanoma | KRAS | MUT | 13 | 697 | 0.01865 | 0.17580 | 5 | 349 | 0.01433 | 0.15257 |
| Melanoma | MAP2K1 | MUT | 15 | 697 | 0.02152 | 0.00412 | 17 | 349 | 0.04871 | 0.00629 |
| Melanoma | MAP2K2 | MUT | 3 | 697 | 0.00430 | 0.00022 | 2 | 349 | 0.00573 | 0.00058 |
| Melanoma | MAP2K4 | MUT | 0 | 697 | 0.00000 | 0.00092 | 0 | 349 | 0.00000 | 0.00687 |

| | | | | | | | | | | |
|---|---|---|---|---|---|---|---|---|---|---|
| Melanoma | MAPK1 | MUT | 0 | 697 | 0.00000 | 0.00079 | 0 | 349 | 0.00000 | 0.00116 |
| Melanoma | MDM2 | AMP | 20 | 697 | 0.02869 | 0.02772 | 9 | 349 | 0.02579 | 0.03696 |
| Melanoma | MET | AMP | 3 | 697 | 0.00430 | 0.00693 | 5 | 349 | 0.01433 | 0.00948 |
| Melanoma | MET | MUT | 0 | 697 | 0.00000 | 0.00355 | 0 | 349 | 0.00000 | 0.00300 |
| Melanoma | MTOR | MUT | 2 | 697 | 0.00287 | 0.00390 | 1 | 349 | 0.00287 | 0.00619 |
| Melanoma | MYC | AMP | 9 | 697 | 0.01291 | 0.04066 | 21 | 349 | 0.06017 | 0.03957 |
| Melanoma | MYCN | AMP | 1 | 697 | 0.00143 | 0.00325 | 0 | 349 | 0.00000 | 0.00484 |
| Melanoma | MYD88 | MUT | 0 | 697 | 0.00000 | 0.00118 | 0 | 349 | 0.00000 | 0.00145 |
| Melanoma | NOTCH1 | FUSION | 0 | 697 | 0.00000 | 0.00018 | 0 | 349 | 0.00000 | 0.00048 |
| Melanoma | NRAS | MUT | 163 | 697 | 0.23386 | 0.02048 | 74 | 349 | 0.21203 | 0.02070 |
| Melanoma | NRG1 | FUSION | 0 | 697 | 0.00000 | 0.00039 | 0 | 349 | 0.00000 | 0.00039 |
| Melanoma | NTRK1 | FUSION | 0 | 697 | 0.00000 | 0.00026 | 2 | 349 | 0.00573 | 0.00097 |
| Melanoma | NTRK3 | FUSION | 0 | 697 | 0.00000 | 0.00066 | 0 | 349 | 0.00000 | 0.00097 |
| Melanoma | PDGFRA | AMP | 4 | 697 | 0.00574 | 0.00627 | 7 | 349 | 0.02006 | 0.00851 |
| Melanoma | PDGFRA | MUT | 0 | 697 | 0.00000 | 0.00048 | 0 | 349 | 0.00000 | 0.00223 |
| Melanoma | PIK3CA | AMP | 0 | 697 | 0.00000 | 0.02298 | 1 | 349 | 0.00287 | 0.00697 |
| Melanoma | PIK3CA | MUT | 14 | 697 | 0.02009 | 0.12387 | 7 | 349 | 0.02006 | 0.11474 |
| Melanoma | POLE | MUT | 0 | 697 | 0.00000 | 0.00079 | 0 | 349 | 0.00000 | 0.00252 |
| Melanoma | PRKACA | FUSION | 0 | 697 | 0.00000 | 0.00039 | 0 | 349 | 0.00000 | 0.00087 |
| Melanoma | PTEN | HOMDEL | 36 | 697 | 0.05165 | 0.03228 | 11 | 349 | 0.03152 | 0.02390 |
| Melanoma | PTEN | MUT | 54 | 697 | 0.07747 | 0.06536 | 29 | 349 | 0.08309 | 0.05563 |
| Melanoma | RAF1 | MUT | 3 | 697 | 0.00430 | 0.00154 | 6 | 349 | 0.01719 | 0.00261 |
| Melanoma | RB1 | HOMDEL | 2 | 697 | 0.00287 | 0.02298 | 1 | 349 | 0.00287 | 0.01896 |
| Melanoma | RB1 | MUT | 12 | 697 | 0.01722 | 0.04053 | 13 | 349 | 0.03725 | 0.04092 |
| Melanoma | RET | FUSION | 0 | 697 | 0.00000 | 0.00219 | 0 | 349 | 0.00000 | 0.00281 |
| Melanoma | RET | MUT | 1 | 697 | 0.00143 | 0.00175 | 0 | 349 | 0.00000 | 0.00213 |
| Melanoma | RIT1 | MUT | 0 | 697 | 0.00000 | 0.00105 | 1 | 349 | 0.00287 | 0.00126 |
| Melanoma | ROS1 | FUSION | 1 | 697 | 0.00143 | 0.00149 | 0 | 349 | 0.00000 | 0.00358 |
| Melanoma | ROS1 | MUT | 0 | 697 | 0.00000 | 0.00009 | 0 | 349 | 0.00000 | 0.00019 |
| Melanoma | SF3B1 | MUT | 26 | 697 | 0.03730 | 0.00807 | 29 | 349 | 0.08309 | 0.00803 |
| Melanoma | SMO | MUT | 0 | 697 | 0.00000 | 0.00013 | 0 | 349 | 0.00000 | 0.00039 |
| Melanoma | SPOP | MUT | 0 | 697 | 0.00000 | 0.00355 | 0 | 349 | 0.00000 | 0.00793 |
| Melanoma | TERT | FUSION | 1 | 697 | 0.00143 | 0.00009 | 0 | 349 | 0.00000 | 0.00000 |
| Melanoma | TERT | MUT | 367 | 697 | 0.52654 | 0.09795 | 171 | 349 | 0.48997 | 0.12307 |
| Melanoma | TP53 | HOMDEL | 1 | 697 | 0.00143 | 0.01276 | 1 | 349 | 0.00287 | 0.00900 |
| Melanoma | TP53 | MUT | 140 | 697 | 0.20086 | 0.52404 | 68 | 349 | 0.19484 | 0.41428 |
| Non-Melanoma Skin | AKT1 | MUT | 2 | 132 | 0.01515 | 0.01026 | 0 | 141 | 0.00000 | 0.01045 |
| Non-Melanoma Skin | ALK | AMP | 0 | 132 | 0.00000 | 0.00018 | 0 | 141 | 0.00000 | 0.00039 |
| Non-Melanoma Skin | ALK | FUSION | 0 | 132 | 0.00000 | 0.00570 | 0 | 141 | 0.00000 | 0.00474 |
| Non-Melanoma Skin | ALK | MUT | 0 | 132 | 0.00000 | 0.00061 | 1 | 141 | 0.00709 | 0.00184 |
| Non-Melanoma Skin | AR | AMP | 0 | 132 | 0.00000 | 0.00601 | 0 | 141 | 0.00000 | 0.00842 |
| Non-Melanoma Skin | AR | MUT | 0 | 132 | 0.00000 | 0.00136 | 0 | 141 | 0.00000 | 0.00213 |
| Non-Melanoma Skin | ARAF | MUT | 0 | 132 | 0.00000 | 0.00039 | 0 | 141 | 0.00000 | 0.00068 |
| Non-Melanoma Skin | BRAF | AMP | 0 | 132 | 0.00000 | 0.00250 | 0 | 141 | 0.00000 | 0.00164 |
| Non-Melanoma Skin | BRAF | FUSION | 0 | 132 | 0.00000 | 0.00118 | 0 | 141 | 0.00000 | 0.00368 |
| Non-Melanoma Skin | BRAF | MUT | 3 | 132 | 0.02273 | 0.04106 | 1 | 141 | 0.00709 | 0.04247 |
| Non-Melanoma Skin | CCND1 | AMP | 2 | 132 | 0.01515 | 0.04299 | 3 | 141 | 0.02128 | 0.04189 |
| Non-Melanoma Skin | CDK4 | AMP | 0 | 132 | 0.00000 | 0.01921 | 1 | 141 | 0.00709 | 0.02632 |
| Non-Melanoma Skin | CDK4 | MUT | 0 | 132 | 0.00000 | 0.00061 | 0 | 141 | 0.00000 | 0.00116 |
| Non-Melanoma Skin | CDK6 | AMP | 0 | 132 | 0.00000 | 0.00697 | 0 | 141 | 0.00000 | 0.00590 |
| Non-Melanoma Skin | CDKN2A | HOMDEL | 11 | 132 | 0.08333 | 0.12234 | 3 | 141 | 0.02128 | 0.07498 |
| Non-Melanoma Skin | CDKN2A | MUT | 37 | 132 | 0.28030 | 0.04917 | 28 | 141 | 0.19858 | 0.03599 |
| Non-Melanoma Skin | CTNNB1 | MUT | 1 | 132 | 0.00758 | 0.02601 | 1 | 141 | 0.00709 | 0.02254 |
| Non-Melanoma Skin | EGFR | AMP | 8 | 132 | 0.06061 | 0.02996 | 2 | 141 | 0.01418 | 0.03154 |
| Non-Melanoma Skin | EGFR | MUT | 0 | 132 | 0.00000 | 0.02649 | 2 | 141 | 0.01418 | 0.04112 |
| Non-Melanoma Skin | ERBB2 | AMP | 2 | 132 | 0.01515 | 0.03202 | 1 | 141 | 0.00709 | 0.03938 |
| Non-Melanoma Skin | ERBB2 | MUT | 2 | 132 | 0.01515 | 0.01689 | 2 | 141 | 0.01418 | 0.01945 |
| Non-Melanoma Skin | ERBB3 | MUT | 0 | 132 | 0.00000 | 0.00561 | 0 | 141 | 0.00000 | 0.00987 |
| Non-Melanoma Skin | ERBB4 | MUT | 2 | 132 | 0.01515 | 0.00022 | 2 | 141 | 0.01418 | 0.00155 |
| Non-Melanoma Skin | ESR1 | MUT | 0 | 132 | 0.00000 | 0.00965 | 1 | 141 | 0.00709 | 0.01006 |
| Non-Melanoma Skin | EZH2 | MUT | 0 | 132 | 0.00000 | 0.00140 | 0 | 141 | 0.00000 | 0.00281 |
| Non-Melanoma Skin | FGFR1 | AMP | 1 | 132 | 0.00758 | 0.02816 | 0 | 141 | 0.00000 | 0.02554 |
| Non-Melanoma Skin | FGFR1 | FUSION | 0 | 132 | 0.00000 | 0.00013 | 0 | 141 | 0.00000 | 0.00039 |
| Non-Melanoma Skin | FGFR2 | AMP | 0 | 132 | 0.00000 | 0.00347 | 0 | 141 | 0.00000 | 0.00358 |
| Non-Melanoma Skin | FGFR2 | FUSION | 0 | 132 | 0.00000 | 0.00083 | 0 | 141 | 0.00000 | 0.00300 |
| Non-Melanoma Skin | FGFR2 | MUT | 0 | 132 | 0.00000 | 0.00531 | 1 | 141 | 0.00709 | 0.00435 |
| Non-Melanoma Skin | FGFR3 | AMP | 0 | 132 | 0.00000 | 0.00263 | 0 | 141 | 0.00000 | 0.00387 |
| Non-Melanoma Skin | FGFR3 | FUSION | 0 | 132 | 0.00000 | 0.00294 | 0 | 141 | 0.00000 | 0.00271 |
| Non-Melanoma Skin | FGFR3 | MUT | 1 | 132 | 0.00758 | 0.00588 | 3 | 141 | 0.02128 | 0.01200 |
| Non-Melanoma Skin | FGFR4 | AMP | 0 | 132 | 0.00000 | 0.00075 | 0 | 141 | 0.00000 | 0.00252 |
| Non-Melanoma Skin | GNA11 | MUT | 1 | 132 | 0.00758 | 0.00105 | 0 | 141 | 0.00000 | 0.00184 |
| Non-Melanoma Skin | GNAQ | MUT | 0 | 132 | 0.00000 | 0.00096 | 0 | 141 | 0.00000 | 0.00281 |
| Non-Melanoma Skin | HRAS | MUT | 6 | 132 | 0.04545 | 0.00504 | 3 | 141 | 0.02128 | 0.00658 |
| Non-Melanoma Skin | IDH1 | MUT | 1 | 132 | 0.00758 | 0.01776 | 0 | 141 | 0.00000 | 0.02351 |
| Non-Melanoma Skin | IDH2 | MUT | 0 | 132 | 0.00000 | 0.00281 | 0 | 141 | 0.00000 | 0.00290 |
| Non-Melanoma Skin | JAK1 | MUT | 0 | 132 | 0.00000 | 0.00026 | 3 | 141 | 0.02128 | 0.00755 |

| | | | | | | | | | | |
|---|---|---|---|---|---|---|---|---|---|---|
| Non-Melanoma Skin | JAK2 | MUT | 0 | 132 | 0.00000 | 0.00066 | 0 | 141 | 0.00000 | 0.00019 |
| Non-Melanoma Skin | KIT | AMP | 0 | 132 | 0.00000 | 0.00675 | 0 | 141 | 0.00000 | 0.00832 |
| Non-Melanoma Skin | KIT | MUT | 1 | 132 | 0.00758 | 0.00675 | 0 | 141 | 0.00000 | 0.01335 |
| Non-Melanoma Skin | KRAS | AMP | 0 | 132 | 0.00000 | 0.01614 | 0 | 141 | 0.00000 | 0.01809 |
| Non-Melanoma Skin | KRAS | MUT | 5 | 132 | 0.03788 | 0.17580 | 0 | 141 | 0.00000 | 0.15257 |
| Non-Melanoma Skin | MAP2K1 | MUT | 1 | 132 | 0.00758 | 0.00412 | 0 | 141 | 0.00000 | 0.00629 |
| Non-Melanoma Skin | MAP2K2 | MUT | 0 | 132 | 0.00000 | 0.00022 | 0 | 141 | 0.00000 | 0.00058 |
| Non-Melanoma Skin | MAP2K4 | MUT | 1 | 132 | 0.00758 | 0.00092 | 0 | 141 | 0.00000 | 0.00687 |
| Non-Melanoma Skin | MAPK1 | MUT | 1 | 132 | 0.00758 | 0.00079 | 1 | 141 | 0.00709 | 0.00116 |
| Non-Melanoma Skin | MDM2 | AMP | 0 | 132 | 0.00000 | 0.02772 | 0 | 141 | 0.00000 | 0.03696 |
| Non-Melanoma Skin | MET | AMP | 0 | 132 | 0.00000 | 0.00693 | 0 | 141 | 0.00000 | 0.00948 |
| Non-Melanoma Skin | MET | MUT | 0 | 132 | 0.00000 | 0.00355 | 0 | 141 | 0.00000 | 0.00300 |
| Non-Melanoma Skin | MTOR | MUT | 3 | 132 | 0.02273 | 0.00390 | 0 | 141 | 0.00000 | 0.00619 |
| Non-Melanoma Skin | MYC | AMP | 2 | 132 | 0.01515 | 0.04066 | 4 | 141 | 0.02837 | 0.03957 |
| Non-Melanoma Skin | MYCN | AMP | 0 | 132 | 0.00000 | 0.00325 | 2 | 141 | 0.01418 | 0.00484 |
| Non-Melanoma Skin | MYD88 | MUT | 0 | 132 | 0.00000 | 0.00118 | 0 | 141 | 0.00000 | 0.00145 |
| Non-Melanoma Skin | NOTCH1 | FUSION | 0 | 132 | 0.00000 | 0.00018 | 0 | 141 | 0.00000 | 0.00048 |
| Non-Melanoma Skin | NRAS | MUT | 1 | 132 | 0.00758 | 0.02048 | 1 | 141 | 0.00709 | 0.02070 |
| Non-Melanoma Skin | NRG1 | FUSION | 0 | 132 | 0.00000 | 0.00039 | 0 | 141 | 0.00000 | 0.00039 |
| Non-Melanoma Skin | NTRK1 | FUSION | 0 | 132 | 0.00000 | 0.00026 | 0 | 141 | 0.00000 | 0.00097 |
| Non-Melanoma Skin | NTRK3 | FUSION | 0 | 132 | 0.00000 | 0.00066 | 0 | 141 | 0.00000 | 0.00097 |
| Non-Melanoma Skin | PDGFRA | AMP | 0 | 132 | 0.00000 | 0.00627 | 0 | 141 | 0.00000 | 0.00851 |
| Non-Melanoma Skin | PDGFRA | MUT | 0 | 132 | 0.00000 | 0.00048 | 0 | 141 | 0.00000 | 0.00223 |
| Non-Melanoma Skin | PIK3CA | AMP | 0 | 132 | 0.00000 | 0.02298 | 0 | 141 | 0.00000 | 0.00697 |
| Non-Melanoma Skin | PIK3CA | MUT | 12 | 132 | 0.09091 | 0.12387 | 14 | 141 | 0.09929 | 0.11474 |
| Non-Melanoma Skin | POLE | MUT | 0 | 132 | 0.00000 | 0.00079 | 0 | 141 | 0.00000 | 0.00252 |
| Non-Melanoma Skin | PRKACA | FUSION | 0 | 132 | 0.00000 | 0.00039 | 0 | 141 | 0.00000 | 0.00087 |
| Non-Melanoma Skin | PTEN | HOMDEL | 3 | 132 | 0.02273 | 0.03228 | 1 | 141 | 0.00709 | 0.02390 |
| Non-Melanoma Skin | PTEN | MUT | 6 | 132 | 0.04545 | 0.06536 | 5 | 141 | 0.03546 | 0.05563 |
| Non-Melanoma Skin | RAF1 | MUT | 1 | 132 | 0.00758 | 0.00154 | 0 | 141 | 0.00000 | 0.00261 |
| Non-Melanoma Skin | RB1 | HOMDEL | 3 | 132 | 0.02273 | 0.02298 | 5 | 141 | 0.03546 | 0.01896 |
| Non-Melanoma Skin | RB1 | MUT | 16 | 132 | 0.12121 | 0.04053 | 19 | 141 | 0.13475 | 0.04092 |
| Non-Melanoma Skin | RET | FUSION | 0 | 132 | 0.00000 | 0.00219 | 0 | 141 | 0.00000 | 0.00281 |
| Non-Melanoma Skin | RET | MUT | 0 | 132 | 0.00000 | 0.00175 | 0 | 141 | 0.00000 | 0.00213 |
| Non-Melanoma Skin | RIT1 | MUT | 0 | 132 | 0.00000 | 0.00105 | 0 | 141 | 0.00000 | 0.00126 |
| Non-Melanoma Skin | ROS1 | FUSION | 0 | 132 | 0.00000 | 0.00149 | 0 | 141 | 0.00000 | 0.00358 |
| Non-Melanoma Skin | ROS1 | MUT | 0 | 132 | 0.00000 | 0.00009 | 0 | 141 | 0.00000 | 0.00019 |
| Non-Melanoma Skin | SF3B1 | MUT | 0 | 132 | 0.00000 | 0.00807 | 0 | 141 | 0.00000 | 0.00803 |
| Non-Melanoma Skin | SMO | MUT | 0 | 132 | 0.00000 | 0.00013 | 0 | 141 | 0.00000 | 0.00039 |
| Non-Melanoma Skin | SPOP | MUT | 0 | 132 | 0.00000 | 0.00355 | 1 | 141 | 0.00709 | 0.00793 |
| Non-Melanoma Skin | TERT | FUSION | 0 | 132 | 0.00000 | 0.00009 | 0 | 141 | 0.00000 | 0.00000 |
| Non-Melanoma Skin | TERT | MUT | 47 | 132 | 0.35606 | 0.09795 | 44 | 141 | 0.31206 | 0.12307 |
| Non-Melanoma Skin | TP53 | HOMDEL | 1 | 132 | 0.00758 | 0.01276 | 1 | 141 | 0.00709 | 0.00900 |
| Non-Melanoma Skin | TP53 | MUT | 90 | 132 | 0.68182 | 0.52404 | 73 | 141 | 0.51773 | 0.41428 |
| Ovary | AKT1 | MUT | 7 | 1484 | 0.00472 | 0.01026 | 2 | 215 | 0.00930 | 0.01045 |
| Ovary | ALK | AMP | 1 | 1484 | 0.00067 | 0.00018 | 0 | 215 | 0.00000 | 0.00039 |
| Ovary | ALK | FUSION | 1 | 1484 | 0.00067 | 0.00570 | 0 | 215 | 0.00000 | 0.00474 |
| Ovary | ALK | MUT | 0 | 1484 | 0.00000 | 0.00061 | 0 | 215 | 0.00000 | 0.00184 |
| Ovary | AR | AMP | 0 | 1484 | 0.00000 | 0.00601 | 0 | 215 | 0.00000 | 0.00842 |
| Ovary | AR | MUT | 0 | 1484 | 0.00000 | 0.00136 | 0 | 215 | 0.00000 | 0.00213 |
| Ovary | ARAF | MUT | 0 | 1484 | 0.00000 | 0.00039 | 0 | 215 | 0.00000 | 0.00068 |
| Ovary | BRAF | AMP | 11 | 1484 | 0.00741 | 0.00250 | 0 | 215 | 0.00000 | 0.00164 |
| Ovary | BRAF | FUSION | 1 | 1484 | 0.00067 | 0.00118 | 0 | 215 | 0.00000 | 0.00368 |
| Ovary | BRAF | MUT | 21 | 1484 | 0.01415 | 0.04106 | 3 | 215 | 0.01395 | 0.04247 |
| Ovary | CCND1 | AMP | 13 | 1484 | 0.00876 | 0.04299 | 1 | 215 | 0.00465 | 0.04189 |
| Ovary | CDK4 | AMP | 4 | 1484 | 0.00270 | 0.01921 | 2 | 215 | 0.00930 | 0.02632 |
| Ovary | CDK4 | MUT | 0 | 1484 | 0.00000 | 0.00061 | 0 | 215 | 0.00000 | 0.00116 |
| Ovary | CDK6 | AMP | 0 | 1484 | 0.00000 | 0.00697 | 0 | 215 | 0.00000 | 0.00590 |
| Ovary | CDKN2A | HOMDEL | 50 | 1484 | 0.03369 | 0.12234 | 7 | 215 | 0.03256 | 0.07498 |
| Ovary | CDKN2A | MUT | 13 | 1484 | 0.00876 | 0.04917 | 4 | 215 | 0.01860 | 0.03599 |
| Ovary | CTNNB1 | MUT | 37 | 1484 | 0.02493 | 0.02601 | 4 | 215 | 0.01860 | 0.02254 |
| Ovary | EGFR | AMP | 2 | 1484 | 0.00135 | 0.02996 | 0 | 215 | 0.00000 | 0.03154 |
| Ovary | EGFR | MUT | 0 | 1484 | 0.00000 | 0.02649 | 0 | 215 | 0.00000 | 0.04112 |
| Ovary | ERBB2 | AMP | 27 | 1484 | 0.01819 | 0.03202 | 9 | 215 | 0.04186 | 0.03938 |
| Ovary | ERBB2 | MUT | 9 | 1484 | 0.00606 | 0.01689 | 3 | 215 | 0.01395 | 0.01945 |
| Ovary | ERBB3 | MUT | 2 | 1484 | 0.00135 | 0.00561 | 0 | 215 | 0.00000 | 0.00987 |
| Ovary | ERBB4 | MUT | 0 | 1484 | 0.00000 | 0.00022 | 0 | 215 | 0.00000 | 0.00155 |
| Ovary | ESR1 | MUT | 3 | 1484 | 0.00202 | 0.00965 | 1 | 215 | 0.00465 | 0.01006 |
| Ovary | EZH2 | MUT | 0 | 1484 | 0.00000 | 0.00140 | 0 | 215 | 0.00000 | 0.00281 |
| Ovary | FGFR1 | AMP | 37 | 1484 | 0.02493 | 0.02816 | 3 | 215 | 0.01395 | 0.02554 |
| Ovary | FGFR1 | FUSION | 0 | 1484 | 0.00000 | 0.00013 | 0 | 215 | 0.00000 | 0.00039 |
| Ovary | FGFR2 | AMP | 7 | 1484 | 0.00472 | 0.00347 | 2 | 215 | 0.00930 | 0.00358 |
| Ovary | FGFR2 | FUSION | 0 | 1484 | 0.00000 | 0.00083 | 0 | 215 | 0.00000 | 0.00300 |
| Ovary | FGFR2 | MUT | 11 | 1484 | 0.00741 | 0.00531 | 1 | 215 | 0.00465 | 0.00435 |
| Ovary | FGFR3 | AMP | 6 | 1484 | 0.00404 | 0.00263 | 2 | 215 | 0.00930 | 0.00387 |
| Ovary | FGFR3 | FUSION | 0 | 1484 | 0.00000 | 0.00294 | 0 | 215 | 0.00000 | 0.00271 |

| | | | | | | | | | | |
|---|---|---|---|---|---|---|---|---|---|---|
| Ovary | FGFR3 | MUT | 0 | 1484 | 0.00000 | 0.00588 | 0 | 215 | 0.00000 | 0.01200 |
| Ovary | FGFR4 | AMP | 2 | 1484 | 0.00135 | 0.00075 | 1 | 215 | 0.00465 | 0.00252 |
| Ovary | GNA11 | MUT | 0 | 1484 | 0.00000 | 0.00105 | 0 | 215 | 0.00000 | 0.00184 |
| Ovary | GNAQ | MUT | 0 | 1484 | 0.00000 | 0.00096 | 0 | 215 | 0.00000 | 0.00281 |
| Ovary | HRAS | MUT | 2 | 1484 | 0.00135 | 0.00504 | 0 | 215 | 0.00000 | 0.00658 |
| Ovary | IDH1 | MUT | 0 | 1484 | 0.00000 | 0.01776 | 0 | 215 | 0.00000 | 0.02351 |
| Ovary | IDH2 | MUT | 2 | 1484 | 0.00135 | 0.00281 | 0 | 215 | 0.00000 | 0.00290 |
| Ovary | JAK1 | MUT | 0 | 1484 | 0.00000 | 0.00026 | 0 | 215 | 0.00000 | 0.00755 |
| Ovary | JAK2 | MUT | 0 | 1484 | 0.00000 | 0.00066 | 0 | 215 | 0.00000 | 0.00019 |
| Ovary | KIT | AMP | 5 | 1484 | 0.00337 | 0.00675 | 0 | 215 | 0.00000 | 0.00832 |
| Ovary | KIT | MUT | 1 | 1484 | 0.00067 | 0.00675 | 0 | 215 | 0.00000 | 0.01335 |
| Ovary | KRAS | AMP | 53 | 1484 | 0.03571 | 0.01614 | 12 | 215 | 0.05581 | 0.01809 |
| Ovary | KRAS | MUT | 127 | 1484 | 0.08558 | 0.17580 | 19 | 215 | 0.08837 | 0.15257 |
| Ovary | MAP2K1 | MUT | 2 | 1484 | 0.00135 | 0.00412 | 0 | 215 | 0.00000 | 0.00629 |
| Ovary | MAP2K2 | MUT | 0 | 1484 | 0.00000 | 0.00022 | 0 | 215 | 0.00000 | 0.00058 |
| Ovary | MAP2K4 | MUT | 0 | 1484 | 0.00000 | 0.00092 | 1 | 215 | 0.00465 | 0.00687 |
| Ovary | MAPK1 | MUT | 0 | 1484 | 0.00000 | 0.00079 | 0 | 215 | 0.00000 | 0.00116 |
| Ovary | MDM2 | AMP | 25 | 1484 | 0.01685 | 0.02772 | 2 | 215 | 0.00930 | 0.03696 |
| Ovary | MET | AMP | 3 | 1484 | 0.00202 | 0.00693 | 2 | 215 | 0.00930 | 0.00948 |
| Ovary | MET | MUT | 0 | 1484 | 0.00000 | 0.00355 | 0 | 215 | 0.00000 | 0.00300 |
| Ovary | MTOR | MUT | 0 | 1484 | 0.00000 | 0.00390 | 0 | 215 | 0.00000 | 0.00619 |
| Ovary | MYC | AMP | 82 | 1484 | 0.05526 | 0.04066 | 14 | 215 | 0.06512 | 0.03957 |
| Ovary | MYCN | AMP | 5 | 1484 | 0.00337 | 0.00325 | 0 | 215 | 0.00000 | 0.00484 |
| Ovary | MYD88 | MUT | 1 | 1484 | 0.00067 | 0.00118 | 0 | 215 | 0.00000 | 0.00145 |
| Ovary | NOTCH1 | FUSION | 0 | 1484 | 0.00000 | 0.00018 | 1 | 215 | 0.00465 | 0.00048 |
| Ovary | NRAS | MUT | 15 | 1484 | 0.01011 | 0.02048 | 4 | 215 | 0.01860 | 0.02070 |
| Ovary | NRG1 | FUSION | 0 | 1484 | 0.00000 | 0.00039 | 0 | 215 | 0.00000 | 0.00039 |
| Ovary | NTRK1 | FUSION | 0 | 1484 | 0.00000 | 0.00026 | 0 | 215 | 0.00000 | 0.00097 |
| Ovary | NTRK3 | FUSION | 0 | 1484 | 0.00000 | 0.00066 | 0 | 215 | 0.00000 | 0.00097 |
| Ovary | PDGFRA | AMP | 3 | 1484 | 0.00202 | 0.00627 | 0 | 215 | 0.00000 | 0.00851 |
| Ovary | PDGFRA | MUT | 0 | 1484 | 0.00000 | 0.00048 | 0 | 215 | 0.00000 | 0.00223 |
| Ovary | PIK3CA | AMP | 79 | 1484 | 0.05323 | 0.02298 | 6 | 215 | 0.02791 | 0.00697 |
| Ovary | PIK3CA | MUT | 120 | 1484 | 0.08086 | 0.12387 | 22 | 215 | 0.10233 | 0.11474 |
| Ovary | POLE | MUT | 1 | 1484 | 0.00067 | 0.00079 | 0 | 215 | 0.00000 | 0.00252 |
| Ovary | PRKACA | FUSION | 0 | 1484 | 0.00000 | 0.00039 | 0 | 215 | 0.00000 | 0.00087 |
| Ovary | PTEN | HOMDEL | 21 | 1484 | 0.01415 | 0.03228 | 5 | 215 | 0.02326 | 0.02390 |
| Ovary | PTEN | MUT | 52 | 1484 | 0.03504 | 0.06536 | 10 | 215 | 0.04651 | 0.05563 |
| Ovary | RAF1 | MUT | 1 | 1484 | 0.00067 | 0.00154 | 0 | 215 | 0.00000 | 0.00261 |
| Ovary | RB1 | HOMDEL | 36 | 1484 | 0.02426 | 0.02298 | 6 | 215 | 0.02791 | 0.01896 |
| Ovary | RB1 | MUT | 23 | 1484 | 0.01550 | 0.04053 | 6 | 215 | 0.02791 | 0.04092 |
| Ovary | RET | FUSION | 0 | 1484 | 0.00000 | 0.00219 | 0 | 215 | 0.00000 | 0.00281 |
| Ovary | RET | MUT | 3 | 1484 | 0.00202 | 0.00175 | 0 | 215 | 0.00000 | 0.00213 |
| Ovary | RIT1 | MUT | 2 | 1484 | 0.00135 | 0.00105 | 0 | 215 | 0.00000 | 0.00126 |
| Ovary | ROS1 | FUSION | 2 | 1484 | 0.00135 | 0.00149 | 1 | 215 | 0.00465 | 0.00358 |
| Ovary | ROS1 | MUT | 0 | 1484 | 0.00000 | 0.00009 | 1 | 215 | 0.00465 | 0.00019 |
| Ovary | SF3B1 | MUT | 0 | 1484 | 0.00000 | 0.00807 | 0 | 215 | 0.00000 | 0.00803 |
| Ovary | SMO | MUT | 0 | 1484 | 0.00000 | 0.00013 | 0 | 215 | 0.00000 | 0.00039 |
| Ovary | SPOP | MUT | 0 | 1484 | 0.00000 | 0.00355 | 2 | 215 | 0.00930 | 0.00793 |
| Ovary | TERT | FUSION | 0 | 1484 | 0.00000 | 0.00009 | 0 | 215 | 0.00000 | 0.00000 |
| Ovary | TERT | MUT | 27 | 1484 | 0.01819 | 0.09795 | 7 | 215 | 0.03256 | 0.12307 |
| Ovary | TP53 | HOMDEL | 8 | 1484 | 0.00539 | 0.01276 | 0 | 215 | 0.00000 | 0.00900 |
| Ovary | TP53 | MUT | 1147 | 1484 | 0.77291 | 0.52404 | 153 | 215 | 0.71163 | 0.41428 |
| Pancreas | AKT1 | MUT | 2 | 1080 | 0.00185 | 0.01026 | 1 | 419 | 0.00239 | 0.01045 |
| Pancreas | ALK | AMP | 0 | 1080 | 0.00000 | 0.00018 | 0 | 419 | 0.00000 | 0.00039 |
| Pancreas | ALK | FUSION | 0 | 1080 | 0.00000 | 0.00570 | 1 | 419 | 0.00239 | 0.00474 |
| Pancreas | ALK | MUT | 0 | 1080 | 0.00000 | 0.00061 | 0 | 419 | 0.00000 | 0.00184 |
| Pancreas | AR | AMP | 0 | 1080 | 0.00000 | 0.00601 | 0 | 419 | 0.00000 | 0.00842 |
| Pancreas | AR | MUT | 0 | 1080 | 0.00000 | 0.00136 | 0 | 419 | 0.00000 | 0.00213 |
| Pancreas | ARAF | MUT | 0 | 1080 | 0.00000 | 0.00039 | 0 | 419 | 0.00000 | 0.00068 |
| Pancreas | BRAF | AMP | 0 | 1080 | 0.00000 | 0.00250 | 0 | 419 | 0.00000 | 0.00164 |
| Pancreas | BRAF | FUSION | 6 | 1080 | 0.00556 | 0.00118 | 7 | 419 | 0.01671 | 0.00368 |
| Pancreas | BRAF | MUT | 12 | 1080 | 0.01111 | 0.04106 | 5 | 419 | 0.01193 | 0.04247 |
| Pancreas | CCND1 | AMP | 15 | 1080 | 0.01389 | 0.04299 | 4 | 419 | 0.00955 | 0.04189 |
| Pancreas | CDK4 | AMP | 16 | 1080 | 0.01481 | 0.01921 | 3 | 419 | 0.00716 | 0.02632 |
| Pancreas | CDK4 | MUT | 1 | 1080 | 0.00093 | 0.00061 | 0 | 419 | 0.00000 | 0.00116 |
| Pancreas | CDK6 | AMP | 21 | 1080 | 0.01944 | 0.00697 | 4 | 419 | 0.00955 | 0.00590 |
| Pancreas | CDKN2A | HOMDEL | 297 | 1080 | 0.27500 | 0.12234 | 33 | 419 | 0.07876 | 0.07498 |
| Pancreas | CDKN2A | MUT | 193 | 1080 | 0.17870 | 0.04917 | 54 | 419 | 0.12888 | 0.03599 |
| Pancreas | CTNNB1 | MUT | 17 | 1080 | 0.01574 | 0.02601 | 11 | 419 | 0.02625 | 0.02254 |
| Pancreas | EGFR | AMP | 2 | 1080 | 0.00185 | 0.02996 | 0 | 419 | 0.00000 | 0.03154 |
| Pancreas | EGFR | MUT | 0 | 1080 | 0.00000 | 0.02649 | 0 | 419 | 0.00000 | 0.04112 |
| Pancreas | ERBB2 | AMP | 19 | 1080 | 0.01759 | 0.03202 | 6 | 419 | 0.01432 | 0.03938 |
| Pancreas | ERBB2 | MUT | 5 | 1080 | 0.00463 | 0.01689 | 3 | 419 | 0.00716 | 0.01945 |
| Pancreas | ERBB3 | MUT | 4 | 1080 | 0.00370 | 0.00561 | 1 | 419 | 0.00239 | 0.00987 |
| Pancreas | ERBB4 | MUT | 0 | 1080 | 0.00000 | 0.00022 | 0 | 419 | 0.00000 | 0.00155 |
| Pancreas | ESR1 | MUT | 0 | 1080 | 0.00000 | 0.00965 | 0 | 419 | 0.00000 | 0.01006 |

| | | | | | | | | | | |
|---|---|---|---|---|---|---|---|---|---|---|
| Pancreas | EZH2 | MUT | 0 | 1080 | 0.00000 | 0.00140 | 0 | 419 | 0.00000 | 0.00281 |
| Pancreas | FGFR1 | AMP | 16 | 1080 | 0.01481 | 0.02816 | 8 | 419 | 0.01909 | 0.02554 |
| Pancreas | FGFR1 | FUSION | 0 | 1080 | 0.00000 | 0.00013 | 0 | 419 | 0.00000 | 0.00039 |
| Pancreas | FGFR2 | AMP | 1 | 1080 | 0.00093 | 0.00347 | 0 | 419 | 0.00000 | 0.00358 |
| Pancreas | FGFR2 | FUSION | 1 | 1080 | 0.00093 | 0.00083 | 2 | 419 | 0.00477 | 0.00300 |
| Pancreas | FGFR2 | MUT | 0 | 1080 | 0.00000 | 0.00531 | 0 | 419 | 0.00000 | 0.00435 |
| Pancreas | FGFR3 | AMP | 5 | 1080 | 0.00463 | 0.00263 | 1 | 419 | 0.00239 | 0.00387 |
| Pancreas | FGFR3 | FUSION | 0 | 1080 | 0.00000 | 0.00294 | 0 | 419 | 0.00000 | 0.00271 |
| Pancreas | FGFR3 | MUT | 0 | 1080 | 0.00000 | 0.00588 | 0 | 419 | 0.00000 | 0.01200 |
| Pancreas | FGFR4 | AMP | 1 | 1080 | 0.00093 | 0.00075 | 2 | 419 | 0.00477 | 0.00252 |
| Pancreas | GNA11 | MUT | 0 | 1080 | 0.00000 | 0.00105 | 0 | 419 | 0.00000 | 0.00184 |
| Pancreas | GNAQ | MUT | 0 | 1080 | 0.00000 | 0.00096 | 0 | 419 | 0.00000 | 0.00281 |
| Pancreas | HRAS | MUT | 1 | 1080 | 0.00093 | 0.00504 | 0 | 419 | 0.00000 | 0.00658 |
| Pancreas | IDH1 | MUT | 2 | 1080 | 0.00185 | 0.01776 | 2 | 419 | 0.00477 | 0.02351 |
| Pancreas | IDH2 | MUT | 3 | 1080 | 0.00278 | 0.00281 | 1 | 419 | 0.00239 | 0.00290 |
| Pancreas | JAK1 | MUT | 0 | 1080 | 0.00000 | 0.00026 | 0 | 419 | 0.00000 | 0.00755 |
| Pancreas | JAK2 | MUT | 0 | 1080 | 0.00000 | 0.00066 | 0 | 419 | 0.00000 | 0.00019 |
| Pancreas | KIT | AMP | 1 | 1080 | 0.00093 | 0.00675 | 0 | 419 | 0.00000 | 0.00832 |
| Pancreas | KIT | MUT | 0 | 1080 | 0.00000 | 0.00675 | 0 | 419 | 0.00000 | 0.01335 |
| Pancreas | KRAS | AMP | 28 | 1080 | 0.02593 | 0.01614 | 8 | 419 | 0.01909 | 0.01809 |
| Pancreas | KRAS | MUT | 923 | 1080 | 0.85463 | 0.17580 | 363 | 419 | 0.86635 | 0.15257 |
| Pancreas | MAP2K1 | MUT | 5 | 1080 | 0.00463 | 0.00412 | 0 | 419 | 0.00000 | 0.00629 |
| Pancreas | MAP2K2 | MUT | 0 | 1080 | 0.00000 | 0.00022 | 2 | 419 | 0.00477 | 0.00058 |
| Pancreas | MAP2K4 | MUT | 0 | 1080 | 0.00000 | 0.00092 | 2 | 419 | 0.00477 | 0.00687 |
| Pancreas | MAPK1 | MUT | 0 | 1080 | 0.00000 | 0.00079 | 0 | 419 | 0.00000 | 0.00116 |
| Pancreas | MDM2 | AMP | 22 | 1080 | 0.02037 | 0.02772 | 5 | 419 | 0.01193 | 0.03696 |
| Pancreas | MET | AMP | 1 | 1080 | 0.00093 | 0.00693 | 0 | 419 | 0.00000 | 0.00948 |
| Pancreas | MET | MUT | 0 | 1080 | 0.00000 | 0.00355 | 0 | 419 | 0.00000 | 0.00300 |
| Pancreas | MTOR | MUT | 1 | 1080 | 0.00093 | 0.00390 | 0 | 419 | 0.00000 | 0.00619 |
| Pancreas | MYC | AMP | 46 | 1080 | 0.04259 | 0.04066 | 12 | 419 | 0.02864 | 0.03957 |
| Pancreas | MYCN | AMP | 0 | 1080 | 0.00000 | 0.00325 | 0 | 419 | 0.00000 | 0.00484 |
| Pancreas | MYD88 | MUT | 0 | 1080 | 0.00000 | 0.00118 | 0 | 419 | 0.00000 | 0.00145 |
| Pancreas | NOTCH1 | FUSION | 0 | 1080 | 0.00000 | 0.00018 | 1 | 419 | 0.00239 | 0.00048 |
| Pancreas | NRAS | MUT | 4 | 1080 | 0.00370 | 0.02048 | 1 | 419 | 0.00239 | 0.02070 |
| Pancreas | NRG1 | FUSION | 0 | 1080 | 0.00000 | 0.00039 | 0 | 419 | 0.00000 | 0.00039 |
| Pancreas | NTRK1 | FUSION | 0 | 1080 | 0.00000 | 0.00026 | 0 | 419 | 0.00000 | 0.00097 |
| Pancreas | NTRK3 | FUSION | 1 | 1080 | 0.00093 | 0.00066 | 2 | 419 | 0.00477 | 0.00097 |
| Pancreas | PDGFRA | AMP | 0 | 1080 | 0.00000 | 0.00627 | 0 | 419 | 0.00000 | 0.00851 |
| Pancreas | PDGFRA | MUT | 0 | 1080 | 0.00000 | 0.00048 | 0 | 419 | 0.00000 | 0.00223 |
| Pancreas | PIK3CA | AMP | 7 | 1080 | 0.00648 | 0.02298 | 0 | 419 | 0.00000 | 0.00697 |
| Pancreas | PIK3CA | MUT | 21 | 1080 | 0.01944 | 0.12387 | 12 | 419 | 0.02864 | 0.11474 |
| Pancreas | POLE | MUT | 0 | 1080 | 0.00000 | 0.00079 | 0 | 419 | 0.00000 | 0.00252 |
| Pancreas | PRKACA | FUSION | 1 | 1080 | 0.00093 | 0.00039 | 0 | 419 | 0.00000 | 0.00087 |
| Pancreas | PTEN | HOMDEL | 8 | 1080 | 0.00741 | 0.03228 | 0 | 419 | 0.00000 | 0.02390 |
| Pancreas | PTEN | MUT | 7 | 1080 | 0.00648 | 0.06536 | 4 | 419 | 0.00955 | 0.05563 |
| Pancreas | RAF1 | MUT | 0 | 1080 | 0.00000 | 0.00154 | 0 | 419 | 0.00000 | 0.00261 |
| Pancreas | RB1 | HOMDEL | 5 | 1080 | 0.00463 | 0.02298 | 2 | 419 | 0.00477 | 0.01896 |
| Pancreas | RB1 | MUT | 19 | 1080 | 0.01759 | 0.04053 | 12 | 419 | 0.02864 | 0.04092 |
| Pancreas | RET | FUSION | 1 | 1080 | 0.00093 | 0.00219 | 0 | 419 | 0.00000 | 0.00281 |
| Pancreas | RET | MUT | 0 | 1080 | 0.00000 | 0.00175 | 0 | 419 | 0.00000 | 0.00213 |
| Pancreas | RIT1 | MUT | 0 | 1080 | 0.00000 | 0.00105 | 0 | 419 | 0.00000 | 0.00126 |
| Pancreas | ROS1 | FUSION | 0 | 1080 | 0.00000 | 0.00149 | 1 | 419 | 0.00239 | 0.00358 |
| Pancreas | ROS1 | MUT | 0 | 1080 | 0.00000 | 0.00009 | 0 | 419 | 0.00000 | 0.00019 |
| Pancreas | SF3B1 | MUT | 18 | 1080 | 0.01667 | 0.00807 | 4 | 419 | 0.00955 | 0.00803 |
| Pancreas | SMO | MUT | 0 | 1080 | 0.00000 | 0.00013 | 0 | 419 | 0.00000 | 0.00039 |
| Pancreas | SPOP | MUT | 0 | 1080 | 0.00000 | 0.00355 | 1 | 419 | 0.00239 | 0.00793 |
| Pancreas | TERT | FUSION | 0 | 1080 | 0.00000 | 0.00009 | 0 | 419 | 0.00000 | 0.00000 |
| Pancreas | TERT | MUT | 7 | 1080 | 0.00648 | 0.09795 | 3 | 419 | 0.00716 | 0.12307 |
| Pancreas | TP53 | HOMDEL | 6 | 1080 | 0.00556 | 0.01276 | 0 | 419 | 0.00000 | 0.00900 |
| Pancreas | TP53 | MUT | 771 | 1080 | 0.71389 | 0.52404 | 276 | 419 | 0.65871 | 0.41428 |
| Prostate | AKT1 | MUT | 22 | 1283 | 0.01715 | 0.01026 | 6 | 621 | 0.00966 | 0.01045 |
| Prostate | ALK | AMP | 0 | 1283 | 0.00000 | 0.00018 | 0 | 621 | 0.00000 | 0.00039 |
| Prostate | ALK | FUSION | 1 | 1283 | 0.00078 | 0.00570 | 0 | 621 | 0.00000 | 0.00474 |
| Prostate | ALK | MUT | 0 | 1283 | 0.00000 | 0.00061 | 1 | 621 | 0.00161 | 0.00184 |
| Prostate | AR | AMP | 117 | 1283 | 0.09119 | 0.00601 | 79 | 621 | 0.12721 | 0.00842 |
| Prostate | AR | MUT | 26 | 1283 | 0.02027 | 0.00136 | 21 | 621 | 0.03382 | 0.00213 |
| Prostate | ARAF | MUT | 0 | 1283 | 0.00000 | 0.00039 | 0 | 621 | 0.00000 | 0.00068 |
| Prostate | BRAF | AMP | 8 | 1283 | 0.00624 | 0.00250 | 0 | 621 | 0.00000 | 0.00164 |
| Prostate | BRAF | FUSION | 7 | 1283 | 0.00546 | 0.00118 | 5 | 621 | 0.00805 | 0.00368 |
| Prostate | BRAF | MUT | 12 | 1283 | 0.00935 | 0.04106 | 8 | 621 | 0.01288 | 0.04247 |
| Prostate | CCND1 | AMP | 25 | 1283 | 0.01949 | 0.04299 | 13 | 621 | 0.02093 | 0.04189 |
| Prostate | CDK4 | AMP | 6 | 1283 | 0.00468 | 0.01921 | 5 | 621 | 0.00805 | 0.02632 |
| Prostate | CDK4 | MUT | 0 | 1283 | 0.00000 | 0.00061 | 0 | 621 | 0.00000 | 0.00116 |
| Prostate | CDK6 | AMP | 2 | 1283 | 0.00156 | 0.00697 | 2 | 621 | 0.00322 | 0.00590 |
| Prostate | CDKN2A | HOMDEL | 33 | 1283 | 0.02572 | 0.12234 | 8 | 621 | 0.01288 | 0.07498 |
| Prostate | CDKN2A | MUT | 8 | 1283 | 0.00624 | 0.04917 | 0 | 621 | 0.00000 | 0.03599 |

| | | | | | | | | | | |
|---|---|---|---|---|---|---|---|---|---|---|
| Prostate | CTNNB1 | MUT | 65 | 1283 | 0.05066 | 0.02601 | 22 | 621 | 0.03543 | 0.02254 |
| Prostate | EGFR | AMP | 0 | 1283 | 0.00000 | 0.02996 | 3 | 621 | 0.00483 | 0.03154 |
| Prostate | EGFR | MUT | 0 | 1283 | 0.00000 | 0.02649 | 1 | 621 | 0.00161 | 0.04112 |
| Prostate | ERBB2 | AMP | 1 | 1283 | 0.00078 | 0.03202 | 0 | 621 | 0.00000 | 0.03938 |
| Prostate | ERBB2 | MUT | 1 | 1283 | 0.00078 | 0.01689 | 2 | 621 | 0.00322 | 0.01945 |
| Prostate | ERBB3 | MUT | 1 | 1283 | 0.00078 | 0.00561 | 1 | 621 | 0.00161 | 0.00987 |
| Prostate | ERBB4 | MUT | 0 | 1283 | 0.00000 | 0.00022 | 0 | 621 | 0.00000 | 0.00155 |
| Prostate | ERG | FUSION | 509 | 1283 | 0.39673 | 0.02246 | 307 | 621 | 0.49436 | 0.03028 |
| Prostate | ESR1 | MUT | 0 | 1283 | 0.00000 | 0.00965 | 0 | 621 | 0.00000 | 0.01006 |
| Prostate | ETV1 | FUSION | 5 | 1283 | 0.00390 | 0.00022 | 8 | 621 | 0.01288 | 0.00077 |
| Prostate | ETV4 | FUSION | 5 | 1283 | 0.00390 | 0.00022 | 1 | 621 | 0.00161 | 0.00010 |
| Prostate | ETV5 | FUSION | 3 | 1283 | 0.00234 | 0.00013 | 1 | 621 | 0.00161 | 0.00010 |
| Prostate | EZH2 | MUT | 1 | 1283 | 0.00078 | 0.00140 | 0 | 621 | 0.00000 | 0.00281 |
| Prostate | FGFR1 | AMP | 13 | 1283 | 0.01013 | 0.02816 | 8 | 621 | 0.01288 | 0.02554 |
| Prostate | FGFR1 | FUSION | 0 | 1283 | 0.00000 | 0.00013 | 1 | 621 | 0.00161 | 0.00039 |
| Prostate | FGFR2 | AMP | 1 | 1283 | 0.00078 | 0.00347 | 0 | 621 | 0.00000 | 0.00358 |
| Prostate | FGFR2 | FUSION | 1 | 1283 | 0.00078 | 0.00083 | 0 | 621 | 0.00000 | 0.00300 |
| Prostate | FGFR2 | MUT | 0 | 1283 | 0.00000 | 0.00531 | 0 | 621 | 0.00000 | 0.00435 |
| Prostate | FGFR3 | AMP | 1 | 1283 | 0.00078 | 0.00263 | 2 | 621 | 0.00322 | 0.00387 |
| Prostate | FGFR3 | FUSION | 3 | 1283 | 0.00234 | 0.00294 | 1 | 621 | 0.00161 | 0.00271 |
| Prostate | FGFR3 | MUT | 0 | 1283 | 0.00000 | 0.00588 | 0 | 621 | 0.00000 | 0.01200 |
| Prostate | FGFR4 | AMP | 1 | 1283 | 0.00078 | 0.00075 | 0 | 621 | 0.00000 | 0.00252 |
| Prostate | GNA11 | MUT | 0 | 1283 | 0.00000 | 0.00105 | 0 | 621 | 0.00000 | 0.00184 |
| Prostate | GNAQ | MUT | 0 | 1283 | 0.00000 | 0.00096 | 0 | 621 | 0.00000 | 0.00281 |
| Prostate | HRAS | MUT | 12 | 1283 | 0.00935 | 0.00504 | 5 | 621 | 0.00805 | 0.00658 |
| Prostate | IDH1 | MUT | 3 | 1283 | 0.00234 | 0.01776 | 3 | 621 | 0.00483 | 0.02351 |
| Prostate | IDH2 | MUT | 1 | 1283 | 0.00078 | 0.00281 | 0 | 621 | 0.00000 | 0.00290 |
| Prostate | JAK1 | MUT | 0 | 1283 | 0.00000 | 0.00026 | 9 | 621 | 0.01449 | 0.00755 |
| Prostate | JAK2 | MUT | 1 | 1283 | 0.00078 | 0.00066 | 0 | 621 | 0.00000 | 0.00019 |
| Prostate | KIT | AMP | 2 | 1283 | 0.00156 | 0.00675 | 0 | 621 | 0.00000 | 0.00832 |
| Prostate | KIT | MUT | 0 | 1283 | 0.00000 | 0.00675 | 0 | 621 | 0.00000 | 0.01335 |
| Prostate | KRAS | AMP | 2 | 1283 | 0.00156 | 0.01614 | 2 | 621 | 0.00322 | 0.01809 |
| Prostate | KRAS | MUT | 4 | 1283 | 0.00312 | 0.17580 | 5 | 621 | 0.00805 | 0.15257 |
| Prostate | MAP2K1 | MUT | 0 | 1283 | 0.00000 | 0.00412 | 1 | 621 | 0.00161 | 0.00629 |
| Prostate | MAP2K2 | MUT | 0 | 1283 | 0.00000 | 0.00022 | 0 | 621 | 0.00000 | 0.00058 |
| Prostate | MAP2K4 | MUT | 0 | 1283 | 0.00000 | 0.00092 | 1 | 621 | 0.00161 | 0.00687 |
| Prostate | MAPK1 | MUT | 2 | 1283 | 0.00156 | 0.00079 | 0 | 621 | 0.00000 | 0.00116 |
| Prostate | MDM2 | AMP | 2 | 1283 | 0.00156 | 0.02772 | 5 | 621 | 0.00805 | 0.03696 |
| Prostate | MET | AMP | 0 | 1283 | 0.00000 | 0.00693 | 1 | 621 | 0.00161 | 0.00948 |
| Prostate | MET | MUT | 0 | 1283 | 0.00000 | 0.00355 | 0 | 621 | 0.00000 | 0.00300 |
| Prostate | MTOR | MUT | 3 | 1283 | 0.00234 | 0.00390 | 1 | 621 | 0.00161 | 0.00619 |
| Prostate | MYC | AMP | 42 | 1283 | 0.03274 | 0.04066 | 30 | 621 | 0.04831 | 0.03957 |
| Prostate | MYCN | AMP | 1 | 1283 | 0.00078 | 0.00325 | 0 | 621 | 0.00000 | 0.00484 |
| Prostate | MYD88 | MUT | 1 | 1283 | 0.00078 | 0.00118 | 0 | 621 | 0.00000 | 0.00145 |
| Prostate | NOTCH1 | FUSION | 0 | 1283 | 0.00000 | 0.00018 | 0 | 621 | 0.00000 | 0.00048 |
| Prostate | NRAS | MUT | 1 | 1283 | 0.00078 | 0.02048 | 0 | 621 | 0.00000 | 0.02070 |
| Prostate | NRG1 | FUSION | 0 | 1283 | 0.00000 | 0.00039 | 0 | 621 | 0.00000 | 0.00039 |
| Prostate | NTRK1 | FUSION | 0 | 1283 | 0.00000 | 0.00026 | 0 | 621 | 0.00000 | 0.00097 |
| Prostate | NTRK3 | FUSION | 0 | 1283 | 0.00000 | 0.00066 | 0 | 621 | 0.00000 | 0.00097 |
| Prostate | PDGFRA | AMP | 2 | 1283 | 0.00156 | 0.00627 | 0 | 621 | 0.00000 | 0.00851 |
| Prostate | PDGFRA | MUT | 0 | 1283 | 0.00000 | 0.00048 | 0 | 621 | 0.00000 | 0.00223 |
| Prostate | PIK3CA | AMP | 12 | 1283 | 0.00935 | 0.02298 | 0 | 621 | 0.00000 | 0.00697 |
| Prostate | PIK3CA | MUT | 50 | 1283 | 0.03897 | 0.12387 | 19 | 621 | 0.03060 | 0.11474 |
| Prostate | POLE | MUT | 1 | 1283 | 0.00078 | 0.00079 | 1 | 621 | 0.00161 | 0.00252 |
| Prostate | PRKACA | FUSION | 0 | 1283 | 0.00000 | 0.00039 | 0 | 621 | 0.00000 | 0.00087 |
| Prostate | PTEN | HOMDEL | 239 | 1283 | 0.18628 | 0.03228 | 72 | 621 | 0.11594 | 0.02390 |
| Prostate | PTEN | MUT | 91 | 1283 | 0.07093 | 0.06536 | 38 | 621 | 0.06119 | 0.05563 |
| Prostate | RAF1 | MUT | 0 | 1283 | 0.00000 | 0.00154 | 0 | 621 | 0.00000 | 0.00261 |
| Prostate | RB1 | HOMDEL | 67 | 1283 | 0.05222 | 0.02298 | 28 | 621 | 0.04509 | 0.01896 |
| Prostate | RB1 | MUT | 35 | 1283 | 0.02728 | 0.04053 | 19 | 621 | 0.03060 | 0.04092 |
| Prostate | RET | FUSION | 0 | 1283 | 0.00000 | 0.00219 | 1 | 621 | 0.00161 | 0.00281 |
| Prostate | RET | MUT | 1 | 1283 | 0.00078 | 0.00175 | 0 | 621 | 0.00000 | 0.00213 |
| Prostate | RIT1 | MUT | 0 | 1283 | 0.00000 | 0.00105 | 0 | 621 | 0.00000 | 0.00126 |
| Prostate | ROS1 | FUSION | 0 | 1283 | 0.00000 | 0.00149 | 0 | 621 | 0.00000 | 0.00358 |
| Prostate | ROS1 | MUT | 0 | 1283 | 0.00000 | 0.00009 | 0 | 621 | 0.00000 | 0.00019 |
| Prostate | SF3B1 | MUT | 12 | 1283 | 0.00935 | 0.00807 | 0 | 621 | 0.00000 | 0.00803 |
| Prostate | SMO | MUT | 0 | 1283 | 0.00000 | 0.00013 | 0 | 621 | 0.00000 | 0.00039 |
| Prostate | SPOP | MUT | 75 | 1283 | 0.05846 | 0.00355 | 62 | 621 | 0.09984 | 0.00793 |
| Prostate | TERT | FUSION | 0 | 1283 | 0.00000 | 0.00009 | 0 | 621 | 0.00000 | 0.00000 |
| Prostate | TERT | MUT | 1 | 1283 | 0.00078 | 0.09795 | 2 | 621 | 0.00322 | 0.12307 |
| Prostate | TP53 | HOMDEL | 47 | 1283 | 0.03663 | 0.01276 | 11 | 621 | 0.01771 | 0.00900 |
| Prostate | TP53 | MUT | 446 | 1283 | 0.34762 | 0.52404 | 184 | 621 | 0.29630 | 0.41428 |
| Sarcoma | AKT1 | MUT | 0 | 764 | 0.00000 | 0.01026 | 2 | 673 | 0.00297 | 0.01045 |
| Sarcoma | ALK | AMP | 1 | 764 | 0.00131 | 0.00018 | 1 | 673 | 0.00149 | 0.00039 |
| Sarcoma | ALK | FUSION | 0 | 764 | 0.00000 | 0.00570 | 1 | 673 | 0.00149 | 0.00474 |
| Sarcoma | ALK | MUT | 1 | 764 | 0.00131 | 0.00061 | 1 | 673 | 0.00149 | 0.00184 |

| | | | | | | | | | | |
|---|---|---|---|---|---|---|---|---|---|---|
| Sarcoma | AR | AMP | 0 | 764 | 0.00000 | 0.00601 | 2 | 673 | 0.00297 | 0.00842 |
| Sarcoma | AR | MUT | 0 | 764 | 0.00000 | 0.00136 | 0 | 673 | 0.00000 | 0.00213 |
| Sarcoma | ARAF | MUT | 0 | 764 | 0.00000 | 0.00039 | 1 | 673 | 0.00149 | 0.00068 |
| Sarcoma | BRAF | AMP | 1 | 764 | 0.00131 | 0.00250 | 0 | 673 | 0.00000 | 0.00164 |
| Sarcoma | BRAF | FUSION | 1 | 764 | 0.00131 | 0.00118 | 2 | 673 | 0.00297 | 0.00368 |
| Sarcoma | BRAF | MUT | 3 | 764 | 0.00393 | 0.04106 | 2 | 673 | 0.00297 | 0.04247 |
| Sarcoma | CCND1 | AMP | 1 | 764 | 0.00131 | 0.04299 | 6 | 673 | 0.00892 | 0.04189 |
| Sarcoma | CDK4 | AMP | 82 | 764 | 0.10733 | 0.01921 | 83 | 673 | 0.12333 | 0.02632 |
| Sarcoma | CDK4 | MUT | 0 | 764 | 0.00000 | 0.00061 | 0 | 673 | 0.00000 | 0.00116 |
| Sarcoma | CDK6 | AMP | 0 | 764 | 0.00000 | 0.00697 | 3 | 673 | 0.00446 | 0.00590 |
| Sarcoma | CDKN2A | HOMDEL | 115 | 764 | 0.15052 | 0.12234 | 65 | 673 | 0.09658 | 0.07498 |
| Sarcoma | CDKN2A | MUT | 11 | 764 | 0.01440 | 0.04917 | 10 | 673 | 0.01486 | 0.03599 |
| Sarcoma | CTNNB1 | MUT | 12 | 764 | 0.01571 | 0.02601 | 4 | 673 | 0.00594 | 0.02254 |
| Sarcoma | EGFR | AMP | 1 | 764 | 0.00131 | 0.02996 | 6 | 673 | 0.00892 | 0.03154 |
| Sarcoma | EGFR | MUT | 0 | 764 | 0.00000 | 0.02649 | 0 | 673 | 0.00000 | 0.04112 |
| Sarcoma | ERBB2 | AMP | 0 | 764 | 0.00000 | 0.03202 | 0 | 673 | 0.00000 | 0.03938 |
| Sarcoma | ERBB2 | MUT | 0 | 764 | 0.00000 | 0.01689 | 0 | 673 | 0.00000 | 0.01945 |
| Sarcoma | ERBB3 | MUT | 0 | 764 | 0.00000 | 0.00561 | 0 | 673 | 0.00000 | 0.00987 |
| Sarcoma | ERBB4 | MUT | 0 | 764 | 0.00000 | 0.00022 | 2 | 673 | 0.00297 | 0.00155 |
| Sarcoma | ESR1 | MUT | 1 | 764 | 0.00131 | 0.00965 | 0 | 673 | 0.00000 | 0.01006 |
| Sarcoma | EZH2 | MUT | 1 | 764 | 0.00131 | 0.00140 | 2 | 673 | 0.00297 | 0.00281 |
| Sarcoma | FGFR1 | AMP | 5 | 764 | 0.00654 | 0.02816 | 10 | 673 | 0.01486 | 0.02554 |
| Sarcoma | FGFR1 | FUSION | 0 | 764 | 0.00000 | 0.00013 | 0 | 673 | 0.00000 | 0.00039 |
| Sarcoma | FGFR2 | AMP | 0 | 764 | 0.00000 | 0.00347 | 0 | 673 | 0.00000 | 0.00358 |
| Sarcoma | FGFR2 | FUSION | 0 | 764 | 0.00000 | 0.00083 | 1 | 673 | 0.00149 | 0.00300 |
| Sarcoma | FGFR2 | MUT | 0 | 764 | 0.00000 | 0.00531 | 0 | 673 | 0.00000 | 0.00435 |
| Sarcoma | FGFR3 | AMP | 0 | 764 | 0.00000 | 0.00263 | 1 | 673 | 0.00149 | 0.00387 |
| Sarcoma | FGFR3 | FUSION | 0 | 764 | 0.00000 | 0.00294 | 0 | 673 | 0.00000 | 0.00271 |
| Sarcoma | FGFR3 | MUT | 0 | 764 | 0.00000 | 0.00588 | 0 | 673 | 0.00000 | 0.01200 |
| Sarcoma | FGFR4 | AMP | 1 | 764 | 0.00131 | 0.00075 | 4 | 673 | 0.00594 | 0.00252 |
| Sarcoma | GNA11 | MUT | 0 | 764 | 0.00000 | 0.00105 | 0 | 673 | 0.00000 | 0.00184 |
| Sarcoma | GNAQ | MUT | 0 | 764 | 0.00000 | 0.00096 | 0 | 673 | 0.00000 | 0.00281 |
| Sarcoma | HRAS | MUT | 6 | 764 | 0.00785 | 0.00504 | 5 | 673 | 0.00743 | 0.00658 |
| Sarcoma | IDH1 | MUT | 8 | 764 | 0.01047 | 0.01776 | 7 | 673 | 0.01040 | 0.02351 |
| Sarcoma | IDH2 | MUT | 6 | 764 | 0.00785 | 0.00281 | 1 | 673 | 0.00149 | 0.00290 |
| Sarcoma | JAK1 | MUT | 0 | 764 | 0.00000 | 0.00026 | 2 | 673 | 0.00297 | 0.00755 |
| Sarcoma | JAK2 | MUT | 0 | 764 | 0.00000 | 0.00066 | 0 | 673 | 0.00000 | 0.00019 |
| Sarcoma | KIT | AMP | 10 | 764 | 0.01309 | 0.00675 | 13 | 673 | 0.01932 | 0.00832 |
| Sarcoma | KIT | MUT | 51 | 764 | 0.06675 | 0.00675 | 1 | 673 | 0.00149 | 0.01335 |
| Sarcoma | KRAS | AMP | 4 | 764 | 0.00524 | 0.01614 | 6 | 673 | 0.00892 | 0.01809 |
| Sarcoma | KRAS | MUT | 7 | 764 | 0.00916 | 0.17580 | 7 | 673 | 0.01040 | 0.15257 |
| Sarcoma | MAP2K1 | MUT | 1 | 764 | 0.00131 | 0.00412 | 1 | 673 | 0.00149 | 0.00629 |
| Sarcoma | MAP2K2 | MUT | 0 | 764 | 0.00000 | 0.00022 | 0 | 673 | 0.00000 | 0.00058 |
| Sarcoma | MAP2K4 | MUT | 0 | 764 | 0.00000 | 0.00092 | 0 | 673 | 0.00000 | 0.00687 |
| Sarcoma | MAPK1 | MUT | 0 | 764 | 0.00000 | 0.00079 | 0 | 673 | 0.00000 | 0.00116 |
| Sarcoma | MDM2 | AMP | 97 | 764 | 0.12696 | 0.02772 | 96 | 673 | 0.14264 | 0.03696 |
| Sarcoma | MET | AMP | 1 | 764 | 0.00131 | 0.00693 | 6 | 673 | 0.00892 | 0.00948 |
| Sarcoma | MET | MUT | 0 | 764 | 0.00000 | 0.00355 | 0 | 673 | 0.00000 | 0.00300 |
| Sarcoma | MTOR | MUT | 4 | 764 | 0.00524 | 0.00390 | 0 | 673 | 0.00000 | 0.00619 |
| Sarcoma | MYC | AMP | 21 | 764 | 0.02749 | 0.04066 | 14 | 673 | 0.02080 | 0.03957 |
| Sarcoma | MYCN | AMP | 0 | 764 | 0.00000 | 0.00325 | 1 | 673 | 0.00149 | 0.00484 |
| Sarcoma | MYD88 | MUT | 0 | 764 | 0.00000 | 0.00118 | 0 | 673 | 0.00000 | 0.00145 |
| Sarcoma | NOTCH1 | FUSION | 0 | 764 | 0.00000 | 0.00018 | 0 | 673 | 0.00000 | 0.00048 |
| Sarcoma | NRAS | MUT | 11 | 764 | 0.01440 | 0.02048 | 7 | 673 | 0.01040 | 0.02070 |
| Sarcoma | NRG1 | FUSION | 0 | 764 | 0.00000 | 0.00039 | 0 | 673 | 0.00000 | 0.00039 |
| Sarcoma | NTRK1 | FUSION | 0 | 764 | 0.00000 | 0.00026 | 1 | 673 | 0.00149 | 0.00097 |
| Sarcoma | NTRK3 | FUSION | 0 | 764 | 0.00000 | 0.00066 | 0 | 673 | 0.00000 | 0.00097 |
| Sarcoma | PDGFRA | AMP | 10 | 764 | 0.01309 | 0.00627 | 17 | 673 | 0.02526 | 0.00851 |
| Sarcoma | PDGFRA | MUT | 1 | 764 | 0.00131 | 0.00048 | 0 | 673 | 0.00000 | 0.00223 |
| Sarcoma | PIK3CA | AMP | 3 | 764 | 0.00393 | 0.02298 | 0 | 673 | 0.00000 | 0.00697 |
| Sarcoma | PIK3CA | MUT | 23 | 764 | 0.03010 | 0.12387 | 22 | 673 | 0.03269 | 0.11474 |
| Sarcoma | POLE | MUT | 0 | 764 | 0.00000 | 0.00079 | 0 | 673 | 0.00000 | 0.00252 |
| Sarcoma | PRKACA | FUSION | 0 | 764 | 0.00000 | 0.00039 | 0 | 673 | 0.00000 | 0.00087 |
| Sarcoma | PTEN | HOMDEL | 33 | 764 | 0.04319 | 0.03228 | 25 | 673 | 0.03715 | 0.02390 |
| Sarcoma | PTEN | MUT | 21 | 764 | 0.02749 | 0.06536 | 14 | 673 | 0.02080 | 0.05563 |
| Sarcoma | RAF1 | MUT | 0 | 764 | 0.00000 | 0.00154 | 0 | 673 | 0.00000 | 0.00261 |
| Sarcoma | RB1 | HOMDEL | 105 | 764 | 0.13743 | 0.02298 | 55 | 673 | 0.08172 | 0.01896 |
| Sarcoma | RB1 | MUT | 50 | 764 | 0.06545 | 0.04053 | 32 | 673 | 0.04755 | 0.04092 |
| Sarcoma | RET | FUSION | 0 | 764 | 0.00000 | 0.00219 | 0 | 673 | 0.00000 | 0.00281 |
| Sarcoma | RET | MUT | 0 | 764 | 0.00000 | 0.00175 | 0 | 673 | 0.00000 | 0.00213 |
| Sarcoma | RIT1 | MUT | 0 | 764 | 0.00000 | 0.00105 | 0 | 673 | 0.00000 | 0.00126 |
| Sarcoma | ROS1 | FUSION | 1 | 764 | 0.00131 | 0.00149 | 2 | 673 | 0.00297 | 0.00358 |
| Sarcoma | ROS1 | MUT | 0 | 764 | 0.00000 | 0.00009 | 0 | 673 | 0.00000 | 0.00019 |
| Sarcoma | SF3B1 | MUT | 2 | 764 | 0.00262 | 0.00807 | 1 | 673 | 0.00149 | 0.00803 |
| Sarcoma | SMO | MUT | 0 | 764 | 0.00000 | 0.00013 | 0 | 673 | 0.00000 | 0.00039 |
| Sarcoma | SPOP | MUT | 0 | 764 | 0.00000 | 0.00355 | 0 | 673 | 0.00000 | 0.00793 |

| | | | | | | | | | | |
|---|---|---|---|---|---|---|---|---|---|---|
| Sarcoma | TERT | FUSION | 1 | 764 | 0.00131 | 0.00009 | 0 | 673 | 0.00000 | 0.00000 |
| Sarcoma | TERT | MUT | 36 | 764 | 0.04712 | 0.09795 | 67 | 673 | 0.09955 | 0.12307 |
| Sarcoma | TP53 | HOMDEL | 57 | 764 | 0.07461 | 0.01276 | 37 | 673 | 0.05498 | 0.00900 |
| Sarcoma | TP53 | MUT | 238 | 764 | 0.31152 | 0.52404 | 146 | 673 | 0.21694 | 0.41428 |
| Thyroid | AKT1 | MUT | 2 | 249 | 0.00803 | 0.01026 | 1 | 221 | 0.00452 | 0.01045 |
| Thyroid | ALK | AMP | 0 | 249 | 0.00000 | 0.00018 | 1 | 221 | 0.00452 | 0.00039 |
| Thyroid | ALK | FUSION | 0 | 249 | 0.00000 | 0.00570 | 0 | 221 | 0.00000 | 0.00474 |
| Thyroid | ALK | MUT | 0 | 249 | 0.00000 | 0.00061 | 0 | 221 | 0.00000 | 0.00184 |
| Thyroid | AR | AMP | 0 | 249 | 0.00000 | 0.00601 | 0 | 221 | 0.00000 | 0.00842 |
| Thyroid | AR | MUT | 0 | 249 | 0.00000 | 0.00136 | 0 | 221 | 0.00000 | 0.00213 |
| Thyroid | ARAF | MUT | 0 | 249 | 0.00000 | 0.00039 | 0 | 221 | 0.00000 | 0.00068 |
| Thyroid | BRAF | AMP | 1 | 249 | 0.00402 | 0.00250 | 0 | 221 | 0.00000 | 0.00164 |
| Thyroid | BRAF | FUSION | 1 | 249 | 0.00402 | 0.00118 | 3 | 221 | 0.01357 | 0.00368 |
| Thyroid | BRAF | MUT | 101 | 249 | 0.40562 | 0.04106 | 83 | 221 | 0.37557 | 0.04247 |
| Thyroid | CCND1 | AMP | 0 | 249 | 0.00000 | 0.04299 | 0 | 221 | 0.00000 | 0.04189 |
| Thyroid | CDK4 | AMP | 1 | 249 | 0.00402 | 0.01921 | 1 | 221 | 0.00452 | 0.02632 |
| Thyroid | CDK4 | MUT | 0 | 249 | 0.00000 | 0.00061 | 0 | 221 | 0.00000 | 0.00116 |
| Thyroid | CDK6 | AMP | 0 | 249 | 0.00000 | 0.00697 | 0 | 221 | 0.00000 | 0.00590 |
| Thyroid | CDKN2A | HOMDEL | 11 | 249 | 0.04418 | 0.12234 | 11 | 221 | 0.04977 | 0.07498 |
| Thyroid | CDKN2A | MUT | 7 | 249 | 0.02811 | 0.04917 | 3 | 221 | 0.01357 | 0.03599 |
| Thyroid | CTNNB1 | MUT | 0 | 249 | 0.00000 | 0.02601 | 1 | 221 | 0.00452 | 0.02254 |
| Thyroid | EGFR | AMP | 0 | 249 | 0.00000 | 0.02996 | 0 | 221 | 0.00000 | 0.03154 |
| Thyroid | EGFR | MUT | 0 | 249 | 0.00000 | 0.02649 | 0 | 221 | 0.00000 | 0.04112 |
| Thyroid | ERBB2 | AMP | 0 | 249 | 0.00000 | 0.03202 | 0 | 221 | 0.00000 | 0.03938 |
| Thyroid | ERBB2 | MUT | 0 | 249 | 0.00000 | 0.01689 | 0 | 221 | 0.00000 | 0.01945 |
| Thyroid | ERBB3 | MUT | 0 | 249 | 0.00000 | 0.00561 | 0 | 221 | 0.00000 | 0.00987 |
| Thyroid | ERBB4 | MUT | 0 | 249 | 0.00000 | 0.00022 | 0 | 221 | 0.00000 | 0.00155 |
| Thyroid | ESR1 | MUT | 0 | 249 | 0.00000 | 0.00965 | 0 | 221 | 0.00000 | 0.01006 |
| Thyroid | EZH2 | MUT | 0 | 249 | 0.00000 | 0.00140 | 1 | 221 | 0.00452 | 0.00281 |
| Thyroid | FGFR1 | AMP | 0 | 249 | 0.00000 | 0.02816 | 1 | 221 | 0.00452 | 0.02554 |
| Thyroid | FGFR1 | FUSION | 0 | 249 | 0.00000 | 0.00013 | 0 | 221 | 0.00000 | 0.00039 |
| Thyroid | FGFR2 | AMP | 0 | 249 | 0.00000 | 0.00347 | 0 | 221 | 0.00000 | 0.00358 |
| Thyroid | FGFR2 | FUSION | 0 | 249 | 0.00000 | 0.00083 | 0 | 221 | 0.00000 | 0.00300 |
| Thyroid | FGFR2 | MUT | 0 | 249 | 0.00000 | 0.00531 | 0 | 221 | 0.00000 | 0.00435 |
| Thyroid | FGFR3 | AMP | 1 | 249 | 0.00402 | 0.00263 | 0 | 221 | 0.00000 | 0.00387 |
| Thyroid | FGFR3 | FUSION | 0 | 249 | 0.00000 | 0.00294 | 0 | 221 | 0.00000 | 0.00271 |
| Thyroid | FGFR3 | MUT | 0 | 249 | 0.00000 | 0.00588 | 0 | 221 | 0.00000 | 0.01200 |
| Thyroid | FGFR4 | AMP | 0 | 249 | 0.00000 | 0.00075 | 0 | 221 | 0.00000 | 0.00252 |
| Thyroid | GNA11 | MUT | 0 | 249 | 0.00000 | 0.00105 | 0 | 221 | 0.00000 | 0.00184 |
| Thyroid | GNAQ | MUT | 0 | 249 | 0.00000 | 0.00096 | 0 | 221 | 0.00000 | 0.00281 |
| Thyroid | HRAS | MUT | 7 | 249 | 0.02811 | 0.00504 | 8 | 221 | 0.03620 | 0.00658 |
| Thyroid | IDH1 | MUT | 0 | 249 | 0.00000 | 0.01776 | 1 | 221 | 0.00452 | 0.02351 |
| Thyroid | IDH2 | MUT | 0 | 249 | 0.00000 | 0.00281 | 0 | 221 | 0.00000 | 0.00290 |
| Thyroid | JAK1 | MUT | 0 | 249 | 0.00000 | 0.00026 | 0 | 221 | 0.00000 | 0.00755 |
| Thyroid | JAK2 | MUT | 0 | 249 | 0.00000 | 0.00066 | 0 | 221 | 0.00000 | 0.00019 |
| Thyroid | KIT | AMP | 3 | 249 | 0.01205 | 0.00675 | 1 | 221 | 0.00452 | 0.00832 |
| Thyroid | KIT | MUT | 0 | 249 | 0.00000 | 0.00675 | 0 | 221 | 0.00000 | 0.01335 |
| Thyroid | KRAS | AMP | 0 | 249 | 0.00000 | 0.01614 | 0 | 221 | 0.00000 | 0.01809 |
| Thyroid | KRAS | MUT | 4 | 249 | 0.01606 | 0.17580 | 7 | 221 | 0.03167 | 0.15257 |
| Thyroid | MAP2K1 | MUT | 1 | 249 | 0.00402 | 0.00412 | 0 | 221 | 0.00000 | 0.00629 |
| Thyroid | MAP2K2 | MUT | 0 | 249 | 0.00000 | 0.00022 | 0 | 221 | 0.00000 | 0.00058 |
| Thyroid | MAP2K4 | MUT | 0 | 249 | 0.00000 | 0.00092 | 3 | 221 | 0.01357 | 0.00687 |
| Thyroid | MAPK1 | MUT | 0 | 249 | 0.00000 | 0.00079 | 0 | 221 | 0.00000 | 0.00116 |
| Thyroid | MDM2 | AMP | 0 | 249 | 0.00000 | 0.02772 | 0 | 221 | 0.00000 | 0.03696 |
| Thyroid | MET | AMP | 1 | 249 | 0.00402 | 0.00693 | 0 | 221 | 0.00000 | 0.00948 |
| Thyroid | MET | MUT | 0 | 249 | 0.00000 | 0.00355 | 0 | 221 | 0.00000 | 0.00300 |
| Thyroid | MTOR | MUT | 0 | 249 | 0.00000 | 0.00390 | 1 | 221 | 0.00452 | 0.00619 |
| Thyroid | MYC | AMP | 0 | 249 | 0.00000 | 0.04066 | 0 | 221 | 0.00000 | 0.03957 |
| Thyroid | MYCN | AMP | 0 | 249 | 0.00000 | 0.00325 | 0 | 221 | 0.00000 | 0.00484 |
| Thyroid | MYD88 | MUT | 0 | 249 | 0.00000 | 0.00118 | 0 | 221 | 0.00000 | 0.00145 |
| Thyroid | NOTCH1 | FUSION | 0 | 249 | 0.00000 | 0.00018 | 0 | 221 | 0.00000 | 0.00048 |
| Thyroid | NRAS | MUT | 28 | 249 | 0.11245 | 0.02048 | 36 | 221 | 0.16290 | 0.02070 |
| Thyroid | NRG1 | FUSION | 0 | 249 | 0.00000 | 0.00039 | 0 | 221 | 0.00000 | 0.00039 |
| Thyroid | NTRK1 | FUSION | 0 | 249 | 0.00000 | 0.00026 | 0 | 221 | 0.00000 | 0.00097 |
| Thyroid | NTRK3 | FUSION | 3 | 249 | 0.01205 | 0.00066 | 0 | 221 | 0.00000 | 0.00097 |
| Thyroid | PDGFRA | AMP | 3 | 249 | 0.01205 | 0.00627 | 1 | 221 | 0.00452 | 0.00851 |
| Thyroid | PDGFRA | MUT | 0 | 249 | 0.00000 | 0.00048 | 0 | 221 | 0.00000 | 0.00223 |
| Thyroid | PIK3CA | AMP | 1 | 249 | 0.00402 | 0.02298 | 0 | 221 | 0.00000 | 0.00697 |
| Thyroid | PIK3CA | MUT | 17 | 249 | 0.06827 | 0.12387 | 13 | 221 | 0.05882 | 0.11474 |
| Thyroid | POLE | MUT | 0 | 249 | 0.00000 | 0.00079 | 0 | 221 | 0.00000 | 0.00252 |
| Thyroid | PRKACA | FUSION | 0 | 249 | 0.00000 | 0.00039 | 0 | 221 | 0.00000 | 0.00087 |
| Thyroid | PTEN | HOMDEL | 0 | 249 | 0.00000 | 0.03228 | 1 | 221 | 0.00452 | 0.02390 |
| Thyroid | PTEN | MUT | 10 | 249 | 0.04016 | 0.06536 | 19 | 221 | 0.08597 | 0.05563 |
| Thyroid | RAF1 | MUT | 0 | 249 | 0.00000 | 0.00154 | 0 | 221 | 0.00000 | 0.00261 |
| Thyroid | RB1 | HOMDEL | 1 | 249 | 0.00402 | 0.02298 | 0 | 221 | 0.00000 | 0.01896 |
| Thyroid | RB1 | MUT | 4 | 249 | 0.01606 | 0.04053 | 7 | 221 | 0.03167 | 0.04092 |

| | | | | | | | | | | |
|---|---|---|---|---|---|---|---|---|---|---|
| Thyroid | RET | FUSION | 12 | 249 | 0.04819 | 0.00219 | 8 | 221 | 0.03620 | 0.00281 |
| Thyroid | RET | MUT | 26 | 249 | 0.10442 | 0.00175 | 12 | 221 | 0.05430 | 0.00213 |
| Thyroid | RIT1 | MUT | 0 | 249 | 0.00000 | 0.00105 | 0 | 221 | 0.00000 | 0.00126 |
| Thyroid | ROS1 | FUSION | 0 | 249 | 0.00000 | 0.00149 | 0 | 221 | 0.00000 | 0.00358 |
| Thyroid | ROS1 | MUT | 0 | 249 | 0.00000 | 0.00009 | 0 | 221 | 0.00000 | 0.00019 |
| Thyroid | SF3B1 | MUT | 1 | 249 | 0.00402 | 0.00807 | 0 | 221 | 0.00000 | 0.00803 |
| Thyroid | SMO | MUT | 0 | 249 | 0.00000 | 0.00013 | 0 | 221 | 0.00000 | 0.00039 |
| Thyroid | SPOP | MUT | 0 | 249 | 0.00000 | 0.00355 | 0 | 221 | 0.00000 | 0.00793 |
| Thyroid | TERT | FUSION | 0 | 249 | 0.00000 | 0.00009 | 0 | 221 | 0.00000 | 0.00000 |
| Thyroid | TERT | MUT | 125 | 249 | 0.50201 | 0.09795 | 135 | 221 | 0.61086 | 0.12307 |
| Thyroid | TP53 | HOMDEL | 0 | 249 | 0.00000 | 0.01276 | 5 | 221 | 0.02262 | 0.00900 |
| Thyroid | TP53 | MUT | 33 | 249 | 0.13253 | 0.52404 | 37 | 221 | 0.16742 | 0.41428 |
| Unknown Primary | AKT1 | MUT | 5 | 1042 | 0.00480 | 0.01026 | 4 | 172 | 0.02326 | 0.01045 |
| Unknown Primary | ALK | AMP | 0 | 1042 | 0.00000 | 0.00018 | 0 | 172 | 0.00000 | 0.00039 |
| Unknown Primary | ALK | FUSION | 3 | 1042 | 0.00288 | 0.00570 | 0 | 172 | 0.00000 | 0.00474 |
| Unknown Primary | ALK | MUT | 0 | 1042 | 0.00000 | 0.00061 | 0 | 172 | 0.00000 | 0.00184 |
| Unknown Primary | AR | AMP | 5 | 1042 | 0.00480 | 0.00601 | 0 | 172 | 0.00000 | 0.00842 |
| Unknown Primary | AR | MUT | 0 | 1042 | 0.00000 | 0.00136 | 0 | 172 | 0.00000 | 0.00213 |
| Unknown Primary | ARAF | MUT | 1 | 1042 | 0.00096 | 0.00039 | 0 | 172 | 0.00000 | 0.00068 |
| Unknown Primary | BRAF | AMP | 1 | 1042 | 0.00096 | 0.00250 | 1 | 172 | 0.00581 | 0.00164 |
| Unknown Primary | BRAF | FUSION | 0 | 1042 | 0.00000 | 0.00118 | 1 | 172 | 0.00581 | 0.00368 |
| Unknown Primary | BRAF | MUT | 41 | 1042 | 0.03935 | 0.04106 | 10 | 172 | 0.05814 | 0.04247 |
| Unknown Primary | CCND1 | AMP | 35 | 1042 | 0.03359 | 0.04299 | 4 | 172 | 0.02326 | 0.04189 |
| Unknown Primary | CDK4 | AMP | 7 | 1042 | 0.00672 | 0.01921 | 3 | 172 | 0.01744 | 0.02632 |
| Unknown Primary | CDK4 | MUT | 0 | 1042 | 0.00000 | 0.00061 | 0 | 172 | 0.00000 | 0.00116 |
| Unknown Primary | CDK6 | AMP | 12 | 1042 | 0.01152 | 0.00697 | 0 | 172 | 0.00000 | 0.00590 |
| Unknown Primary | CDKN2A | HOMDEL | 212 | 1042 | 0.20345 | 0.12234 | 16 | 172 | 0.09302 | 0.07498 |
| Unknown Primary | CDKN2A | MUT | 101 | 1042 | 0.09693 | 0.04917 | 11 | 172 | 0.06395 | 0.03599 |
| Unknown Primary | CTNNB1 | MUT | 36 | 1042 | 0.03455 | 0.02601 | 7 | 172 | 0.04070 | 0.02254 |
| Unknown Primary | EGFR | AMP | 11 | 1042 | 0.01056 | 0.02996 | 5 | 172 | 0.02907 | 0.03154 |
| Unknown Primary | EGFR | MUT | 5 | 1042 | 0.00480 | 0.02649 | 1 | 172 | 0.00581 | 0.04112 |
| Unknown Primary | ERBB2 | AMP | 26 | 1042 | 0.02495 | 0.03202 | 8 | 172 | 0.04651 | 0.03938 |
| Unknown Primary | ERBB2 | MUT | 18 | 1042 | 0.01727 | 0.01689 | 5 | 172 | 0.02907 | 0.01945 |
| Unknown Primary | ERBB3 | MUT | 5 | 1042 | 0.00480 | 0.00561 | 2 | 172 | 0.01163 | 0.00987 |
| Unknown Primary | ERBB4 | MUT | 0 | 1042 | 0.00000 | 0.00022 | 0 | 172 | 0.00000 | 0.00155 |
| Unknown Primary | ESR1 | MUT | 1 | 1042 | 0.00096 | 0.00965 | 1 | 172 | 0.00581 | 0.01006 |
| Unknown Primary | EZH2 | MUT | 2 | 1042 | 0.00192 | 0.00140 | 0 | 172 | 0.00000 | 0.00281 |
| Unknown Primary | FGFR1 | AMP | 22 | 1042 | 0.02111 | 0.02816 | 0 | 172 | 0.00000 | 0.02554 |
| Unknown Primary | FGFR1 | FUSION | 0 | 1042 | 0.00000 | 0.00013 | 0 | 172 | 0.00000 | 0.00039 |
| Unknown Primary | FGFR2 | AMP | 4 | 1042 | 0.00384 | 0.00347 | 0 | 172 | 0.00000 | 0.00358 |
| Unknown Primary | FGFR2 | FUSION | 4 | 1042 | 0.00384 | 0.00083 | 2 | 172 | 0.01163 | 0.00300 |
| Unknown Primary | FGFR2 | MUT | 3 | 1042 | 0.00288 | 0.00531 | 1 | 172 | 0.00581 | 0.00435 |
| Unknown Primary | FGFR3 | AMP | 4 | 1042 | 0.00384 | 0.00263 | 0 | 172 | 0.00000 | 0.00387 |
| Unknown Primary | FGFR3 | FUSION | 2 | 1042 | 0.00192 | 0.00294 | 0 | 172 | 0.00000 | 0.00271 |
| Unknown Primary | FGFR3 | MUT | 5 | 1042 | 0.00480 | 0.00588 | 0 | 172 | 0.00000 | 0.01200 |
| Unknown Primary | FGFR4 | AMP | 0 | 1042 | 0.00000 | 0.00075 | 0 | 172 | 0.00000 | 0.00252 |
| Unknown Primary | GNA11 | MUT | 0 | 1042 | 0.00000 | 0.00105 | 0 | 172 | 0.00000 | 0.00184 |
| Unknown Primary | GNAQ | MUT | 0 | 1042 | 0.00000 | 0.00096 | 0 | 172 | 0.00000 | 0.00281 |
| Unknown Primary | HRAS | MUT | 9 | 1042 | 0.00864 | 0.00504 | 0 | 172 | 0.00000 | 0.00658 |
| Unknown Primary | IDH1 | MUT | 22 | 1042 | 0.02111 | 0.01776 | 0 | 172 | 0.00000 | 0.02351 |
| Unknown Primary | IDH2 | MUT | 5 | 1042 | 0.00480 | 0.00281 | 1 | 172 | 0.00581 | 0.00290 |
| Unknown Primary | JAK1 | MUT | 1 | 1042 | 0.00096 | 0.00026 | 0 | 172 | 0.00000 | 0.00755 |
| Unknown Primary | JAK2 | MUT | 1 | 1042 | 0.00096 | 0.00066 | 0 | 172 | 0.00000 | 0.00019 |
| Unknown Primary | KIT | AMP | 4 | 1042 | 0.00384 | 0.00675 | 0 | 172 | 0.00000 | 0.00832 |
| Unknown Primary | KIT | MUT | 4 | 1042 | 0.00384 | 0.00675 | 0 | 172 | 0.00000 | 0.01335 |
| Unknown Primary | KRAS | AMP | 25 | 1042 | 0.02399 | 0.01614 | 2 | 172 | 0.01163 | 0.01809 |
| Unknown Primary | KRAS | MUT | 214 | 1042 | 0.20537 | 0.17580 | 37 | 172 | 0.21512 | 0.15257 |
| Unknown Primary | MAP2K1 | MUT | 12 | 1042 | 0.01152 | 0.00412 | 1 | 172 | 0.00581 | 0.00629 |
| Unknown Primary | MAP2K2 | MUT | 0 | 1042 | 0.00000 | 0.00022 | 1 | 172 | 0.00581 | 0.00058 |
| Unknown Primary | MAP2K4 | MUT | 2 | 1042 | 0.00192 | 0.00092 | 1 | 172 | 0.00581 | 0.00687 |
| Unknown Primary | MAPK1 | MUT | 1 | 1042 | 0.00096 | 0.00079 | 0 | 172 | 0.00000 | 0.00116 |
| Unknown Primary | MDM2 | AMP | 20 | 1042 | 0.01919 | 0.02772 | 2 | 172 | 0.01163 | 0.03696 |
| Unknown Primary | MET | AMP | 17 | 1042 | 0.01631 | 0.00693 | 1 | 172 | 0.00581 | 0.00948 |
| Unknown Primary | MET | MUT | 2 | 1042 | 0.00192 | 0.00355 | 0 | 172 | 0.00000 | 0.00300 |
| Unknown Primary | MTOR | MUT | 0 | 1042 | 0.00000 | 0.00390 | 0 | 172 | 0.00000 | 0.00619 |
| Unknown Primary | MYC | AMP | 45 | 1042 | 0.04319 | 0.04066 | 6 | 172 | 0.03488 | 0.03957 |
| Unknown Primary | MYCN | AMP | 5 | 1042 | 0.00480 | 0.00325 | 0 | 172 | 0.00000 | 0.00484 |
| Unknown Primary | MYD88 | MUT | 1 | 1042 | 0.00096 | 0.00118 | 0 | 172 | 0.00000 | 0.00145 |
| Unknown Primary | NOTCH1 | FUSION | 0 | 1042 | 0.00000 | 0.00018 | 0 | 172 | 0.00000 | 0.00048 |
| Unknown Primary | NRAS | MUT | 25 | 1042 | 0.02399 | 0.02048 | 5 | 172 | 0.02907 | 0.02070 |
| Unknown Primary | NRG1 | FUSION | 1 | 1042 | 0.00096 | 0.00039 | 1 | 172 | 0.00581 | 0.00039 |
| Unknown Primary | NTRK1 | FUSION | 0 | 1042 | 0.00000 | 0.00026 | 1 | 172 | 0.00581 | 0.00097 |
| Unknown Primary | NTRK3 | FUSION | 1 | 1042 | 0.00096 | 0.00066 | 0 | 172 | 0.00000 | 0.00097 |
| Unknown Primary | PDGFRA | AMP | 4 | 1042 | 0.00384 | 0.00627 | 0 | 172 | 0.00000 | 0.00851 |
| Unknown Primary | PDGFRA | MUT | 0 | 1042 | 0.00000 | 0.00048 | 0 | 172 | 0.00000 | 0.00223 |
| Unknown Primary | PIK3CA | AMP | 19 | 1042 | 0.01823 | 0.02298 | 0 | 172 | 0.00000 | 0.00697 |

| | | | | | | | | | | |
|---|---|---|---|---|---|---|---|---|---|---|
| Unknown Primary | PIK3CA | MUT | 92 | 1042 | 0.08829 | 0.12387 | 16 | 172 | 0.09302 | 0.11474 |
| Unknown Primary | POLE | MUT | 0 | 1042 | 0.00000 | 0.00079 | 1 | 172 | 0.00581 | 0.00252 |
| Unknown Primary | PRKACA | FUSION | 1 | 1042 | 0.00096 | 0.00039 | 0 | 172 | 0.00000 | 0.00087 |
| Unknown Primary | PTEN | HOMDEL | 25 | 1042 | 0.02399 | 0.03228 | 4 | 172 | 0.02326 | 0.02390 |
| Unknown Primary | PTEN | MUT | 58 | 1042 | 0.05566 | 0.06536 | 5 | 172 | 0.02907 | 0.05563 |
| Unknown Primary | RAF1 | MUT | 1 | 1042 | 0.00096 | 0.00154 | 0 | 172 | 0.00000 | 0.00261 |
| Unknown Primary | RB1 | HOMDEL | 29 | 1042 | 0.02783 | 0.02298 | 2 | 172 | 0.01163 | 0.01896 |
| Unknown Primary | RB1 | MUT | 66 | 1042 | 0.06334 | 0.04053 | 11 | 172 | 0.06395 | 0.04092 |
| Unknown Primary | RET | FUSION | 0 | 1042 | 0.00000 | 0.00219 | 0 | 172 | 0.00000 | 0.00281 |
| Unknown Primary | RET | MUT | 0 | 1042 | 0.00000 | 0.00175 | 2 | 172 | 0.01163 | 0.00213 |
| Unknown Primary | RIT1 | MUT | 2 | 1042 | 0.00192 | 0.00105 | 0 | 172 | 0.00000 | 0.00126 |
| Unknown Primary | ROS1 | FUSION | 0 | 1042 | 0.00000 | 0.00149 | 1 | 172 | 0.00581 | 0.00358 |
| Unknown Primary | ROS1 | MUT | 0 | 1042 | 0.00000 | 0.00009 | 0 | 172 | 0.00000 | 0.00019 |
| Unknown Primary | SF3B1 | MUT | 7 | 1042 | 0.00672 | 0.00807 | 1 | 172 | 0.00581 | 0.00803 |
| Unknown Primary | SMO | MUT | 0 | 1042 | 0.00000 | 0.00013 | 0 | 172 | 0.00000 | 0.00039 |
| Unknown Primary | SPOP | MUT | 0 | 1042 | 0.00000 | 0.00355 | 3 | 172 | 0.01744 | 0.00793 |
| Unknown Primary | TERT | FUSION | 0 | 1042 | 0.00000 | 0.00009 | 0 | 172 | 0.00000 | 0.00000 |
| Unknown Primary | TERT | MUT | 98 | 1042 | 0.09405 | 0.09795 | 12 | 172 | 0.06977 | 0.12307 |
| Unknown Primary | TP53 | HOMDEL | 9 | 1042 | 0.00864 | 0.01276 | 0 | 172 | 0.00000 | 0.00900 |
| Unknown Primary | TP53 | MUT | 585 | 1042 | 0.56142 | 0.52404 | 80 | 172 | 0.46512 | 0.41428 |

Prioritized biomarker frequencies in this PCR-CGP cohort are compared to those in the Memorial Sloan Kettering (MSK)-IMPACT pan-solid tumor, hybrid capture based CGP cohort of 10,917 clinical tumor samples. For each tumor type with ≥ 50 samples in each cohort, biomarker frequencies were determined per genes and variant class: MUT (mutations [SNVs and indels], HOMDEL (deep deletions), AMP (amplification), and FUSION (gene fusions). Per cohort, the number of samples per tumor type with the indicated variant (Variant *n*), the total number of samples in that tumor type (Sample *n*), the variant frequency in the cancer type (Cancer Type Freq), and the overall variant frequency (Overall Freq) are shown. To facilitate comparison, several PCR-CGP tumor types were combined in this analysis ('Biliary_Liver': Biliary and Liver, and 'Esophagus_Stomach': Esophagus and Stomach). Samples with multiple reported variants in a single gene + biomarker category were only counted once in corresponding frequency calculations. See **Methods** for additional details.

**Supplementary Table 8. PCR-CGP vs. MSK-IMPACT Biomarker Frequency Correlations**

| Cancer Type | Sample Number | | Pearson *r* |
|---|---|---|---|
| | PCR-CGP | MSK-IMPACT | |
| Biliary_Liver | 558 | 343 | 0.932 |
| Bladder | 614 | 402 | 0.961 |
| Brain | 1,203 | 513 | 0.987 |
| Breast | 2,659 | 1,228 | 0.990 |
| Colon and Rectum | 3,020 | 1,053 | 0.999 |
| Endometrium | 919 | 210 | 0.982 |
| Esophagus_Stomach | 994 | 315 | 0.972 |
| Head and Neck | 746 | 282 | 0.957 |
| Kidney | 510 | 322 | 0.897 |
| Lung - NSCLC | 2,784 | 1,539 | 0.980 |
| Lung - SCLC | 433 | 81 | 0.990 |
| Lymphoma | 182 | 171 | 0.941 |
| Melanoma | 697 | 349 | 0.982 |
| Non-Melanoma Skin | 132 | 141 | 0.986 |
| Ovary | 1,484 | 215 | 0.996 |
| Pancreas | 1,080 | 419 | 0.983 |
| Prostate | 1,283 | 621 | 0.969 |
| Sarcoma | 764 | 673 | 0.939 |
| Thyroid | 249 | 221 | 0.983 |
| Unknown Primary | 1,042 | 172 | 0.975 |
| **Overall** | **21,353** | **9,270** | **0.990** |

Prioritized biomarker frequencies in this PCR-CGP cohort are compared to those in the Memorial Sloan Kettering (MSK)-IMPACT pan-solid tumor, hybrid capture based CGP cohort. For each tumor type with ≥ 50 samples in each cohort, biomarker rates were determined per genes and variant class as shown in **Supplementary Table 7**. For each indicated tumor type, the total sample number per cohort, along with the Pearson correlation (*r*) of all included biomarker rates are shown. The overall sample number of included samples for both cohorts and the overall correlation of all biomarkers is also shown.

**Supplementary Table 9. Informative PCR-CGP for Prostate Cancer**

| Specimen | | Prostate Cancer | | PCR-CGP Results | | | |
|---|---|---|---|---|---|---|---|
| | | | | Informative | | Informative (Positive) | |
| Requirement Status | Attribute | n | % | n | % | n | % |
| **Pass** | Large (>25mm$^2$) | 275 | 20.5% | 266 | 96.7% | 37 | 13.5% |
| | Small (2-25mm$^2$) | 617 | 45.9% | 588 | 95.3% | 95 | 15.4% |
| | **Subtotal** | **892** | **66.4%** | **854** | **95.7%** | **132** | **14.8%** |
| **Exception** | TC < 20% | 88 | 6.5% | 6 | 6.8% | 6 | 6.8% |
| | TSA < 2mm$^2$ | 61 | 4.5% | 8 | 13.1% | 8 | 13.1% |
| | Age > 5 years | 145 | 10.8% | 100 | 69.0% | 14 | 9.7% |
| | DNA/RNA < 1ng/uL | 158 | 11.8% | 115 | 72.8% | 28 | 17.7% |
| | **Subtotal** | **452** | **33.6%** | **229** | **50.7%** | **56** | **12.4%** |
| | ***Total*** | ***1,344*** | ***100.0%*** | ***1,083*** | ***80.6%*** | ***853*** | ***87.8%*** |

This table summarizes overall sample characteristics and infromative test reports in 1,344 consecutive prostate cancer samples tested by the current StrataNGS PCR-CGP test version. Sample requirement status indicates whether a given sample met all minimum PCR-CGP test requirements ('Pass': tumor surface area (TSA) >= 2mm2, tumor content (TC) >= 20%, sample age <= 5 years, and DNA & RNA concentration >= 1ng/uL), or failed to meet at least one of these requirements ('Exception'). Here, PCR-CGP results were only considered Informative if the sample had at least one positive actionable biomarker result (microsatellite instable high (MSI-H) or deleterious mutation/copy number deep deletion in *MSH2/6, BRCA1/2* or *ATM*, OR the sample passed all sequencing quality control QC metrics and had >=20% TC. *'Informative (Positive)'* results indicate that the sample contained at least one positive actionable biomarker result.

**Supplementary Table 10: Informative PCR-CGP for NSCLC Adenocarcinoma**

| Specimen | | NSCLC | | PCR-CGP Results | | | |
|---|---|---|---|---|---|---|---|
| Requirement Status | Attribute | n | % | Informative | | Informative (Positive) | |
| | | | | n | % | n | % |
| **Pass** | Large (>25mm$^2$) | 194 | 17.0% | 190 | 97.9% | 153 | 78.9% |
| | Small (2-25mm$^2$) | 484 | 42.3% | 476 | 98.3% | 383 | 79.1% |
| | **Subtotal** | **678** | **59.3%** | **666** | **98.2%** | **536** | **79.1%** |
| **Exception** | TC < 20% | 249 | 21.8% | 144 | 57.8% | 144 | 57.8% |
| | TSA < 2mm$^2$ | 138 | 12.1% | 125 | 90.6% | 109 | 79.0% |
| | Age > 5 years | 10 | 0.9% | 8 | 80.0% | 8 | 80.0% |
| | DNA/RNA < 1ng/uL | 69 | 6.0% | 61 | 88.4% | 56 | 81.2% |
| | **Subtotal** | **466** | **40.7%** | **338** | **72.5%** | **317** | **68.0%** |
| | ***Total*** | ***1,144*** | ***100.0%*** | ***1,004*** | ***87.8%*** | ***853*** | ***74.6%*** |

This table summarizes overall sample characteristics and infromative test reports in 1,144 consecutive non-small cell lung cancer (NSCLC) tissue specimens tested by the current StrataNGS PCR-CGP test version when NSCLC subtype (adenocarcinoma) was prospectively determined at the time of histopathologic review. Sample requirement status indicates whether a given sample met all minimum PCR-CGP test requirements ('Pass': tumor surface area (TSA) >= 2mm$^2$, tumor content (TC) >= 20%, sample age <= 5 years, and DNA & RNA concentration >= 1ng/uL), or failed to meet at least one of these requirements ('Exception'). Here, PCR-CGP results were only considered Informative if the sample had at least one positive therapy informative/actionable biomarker result (see **Supplementary Table 2**) OR the sample passed all sequencing quality control (QC) metrics and had >=20% TC. '*Informative (Positive)'* results indicate that the sample contained at least one positive therapy informative/actionable biomarker result.
